# A Conformable CMOS Ultrasound System for Point-of-Care Imaging

**DOI:** 10.64898/2026.06.23.26356282

**Authors:** Jagannaath Shiva Letchumanan, Siddhesh Gandhi, Heyu Yin, Samuel G. Blackman, Jason D. Fabbri, Elisa E. Konofagou, David O. Kessler, Kenneth L. Shepard

## Abstract

Point-of-care ultrasound has transformed bedside diagnostics, yet current systems remain limited by rigid form factors, bulky external electronics and the need for skilled operators. Here we report a conformable ultrasound imaging patch that integrates a 1024-channel CMOS ultrasound application-specific integrated circuit (ASIC) directly beneath a conformable piezocomposite transducer array. The 10 mm × 8 mm, 1024-element ASIC contains on-chip transmit and receive beamforming, reducing the effective off-chip channel count by 16× while preserving image fidelity. Fabricated on a flexible polyimide substrate and bonded using anisotropic conductive film, the patch operates untethered from conventional ultrasound consoles and requires only a laptop for control and data acquisition. The device supports focused, plane-wave and diverging-wave transmission with steering over ±30° in azimuth and ±15° in elevation, achieving peak-to-peak acoustic pressures up to 7 MPa at a 4.4-MHz center frequency (mechanical index of 1.7), within diagnostic safety limits. Phantom experiments demonstrate three-dimensional imaging with axial and lateral resolutions (in both XZ and YZ planes) of 0.5 mm and 2 mm, respectively, and accurate contrast reproduction in tissue-mimicking phantoms. Human studies further demonstrate three-dimensional (3D) visualization of the internal jugular vein and carotid artery, as well as rib-shadow-free imaging of pleural motion during respiration. This work establishes a scalable architecture for chronic, wearable ultrasound imaging and highlights the potential of CMOS-integrated, conformable ultrasound systems for continuous physiological monitoring and remote diagnostics.

## INTRODUCTION

Point-of-care ultrasound (POCUS)^1,2^ has been revolutionizing healthcare by enabling early diagnoses and greatly improving the time to care for patients in remote regions or those with poor healthcare infrastructure where X-ray and MRI scanners are not likely to be found. POCUS has reduced the median time to diagnosis in emergency departments^3^ and is indicated for a broad spectrum of conditions from hemodynamic assessments of the carotid and internal jugular vein^4,5^ to diagnosing pneumothorax^6,7^, pericardial effusion^8,9^, abnormal aortic aneurysm^8^ and deep vein thrombosis^9^ amongst others, with high sensitivity and specificity (above 85% for most applications)^9^. POCUS is also particularly useful in resource-constrained environments, such as combat casualty care. Consequently, its adoption has seen a growing trend, with over 70% of emergency departments in France and 90% of rural emergency practitioners in Canada having access to POCUS^9^.

These POCUS systems take the form of hand-held probes with active clinician involvement in their use. In contrast, after initial device placement, wearable ultrasound imagers promise continuous physiological tracking in the absence of trained clinicians, enabling remote monitoring of patients. With a mechanically conformable form factor, these imagers can maintain good acoustic coupling over body curvature, often without the need for contact gel. These potential advantages have resulted in significant interest in the development of wearable transducer arrays^10–16^. Unfortunately, all of these systems require wiring to bulky external electronics, such as the Verasonics Vantage 256 system. This tethered wiring defies characterizing these systems as true wearable devices. Additionally, the lack of front-end electronics close to the transducers leads to long interconnect to the transceiver electronics, introducing loss and lowering achievable signal- to-noise ratios (SNRs). Any systems that have incorporated electronics have done so at the scale of a single channel^17,18^ or few channels^19^ multiplexed to a larger number of transducers, significantly limiting the frame rate that can be achieved.

Because of the growing interest in ultrasound applications, there have been a number of efforts directed toward the development of application-specific integrated circuits (ASICs) integrating some of the electronics required for an ultrasound system^20–23^. These systems take one of two forms; an ASIC bonded to an array of capacitive machined ultrasound transducers (CMUTs) or a system in which bulk piezoelectric transducers are integrated onto the ASIC. The former has achieved the best imaging results since the acoustic environment of the CMUTs is unaffected by the presence of the ASIC^24^. In the latter case, these devices have been implemented on rigid CMOS substrates on rigid printed circuit boards with no consideration of the acoustic environment required to reach bandwidths sufficient for *in-vivo* imaging^20–23^. None of these systems have been implemented as conformable devices (see **Supplementary Table S1**).

In this work, we present FlexUS, a conformable two-dimensional ultrasound imaging device (**Fig. 1a**). This is made possible by combining several innovations, including a custom ASIC design that incorporates on-chip beamforming at the scale of 1024 channels; monolithic integration of mechanically flexible piezoelectric composites; and use of thinned CMOS, polyimide packaging, and appropriate backing and matching materials to provide both conformability and acoustic performance to enable transmit pressures of up to 145 kPa per channel (measured at 0.5 mm from the face of the device), receiver sensitivities of 245 nV/Pa, and receiver fractional bandwidths (FBW) in excess of 34.5% at the 5-MHz operating frequency. The system supports focused-beam peak-to-peak pressures of up to 7 MPa (measured for 20 mm focal distance setting). With these capabilities, we explore how FlexUS can be used for several applications that take advantage of the volumetric imaging capabilities of this two-dimensional (2D) array. We consider use in guiding and monitoring central venous catheters (CVCs), for which ultrasound-guided placement^25^ significantly reduces the risk of infection, air embolism, pneumothorax and bleeding^26^. In addition, with an aperture size of 5.74 mm × 5.74 mm, FlexUS can be positioned intercostally, i.e., between the ribs. With this positioning, FlexUS can be used for observing lung sliding, and artifact-free imaging of the pleural walls allows for diagnosis of conditions such as pneumothorax.

**Figure 1.**
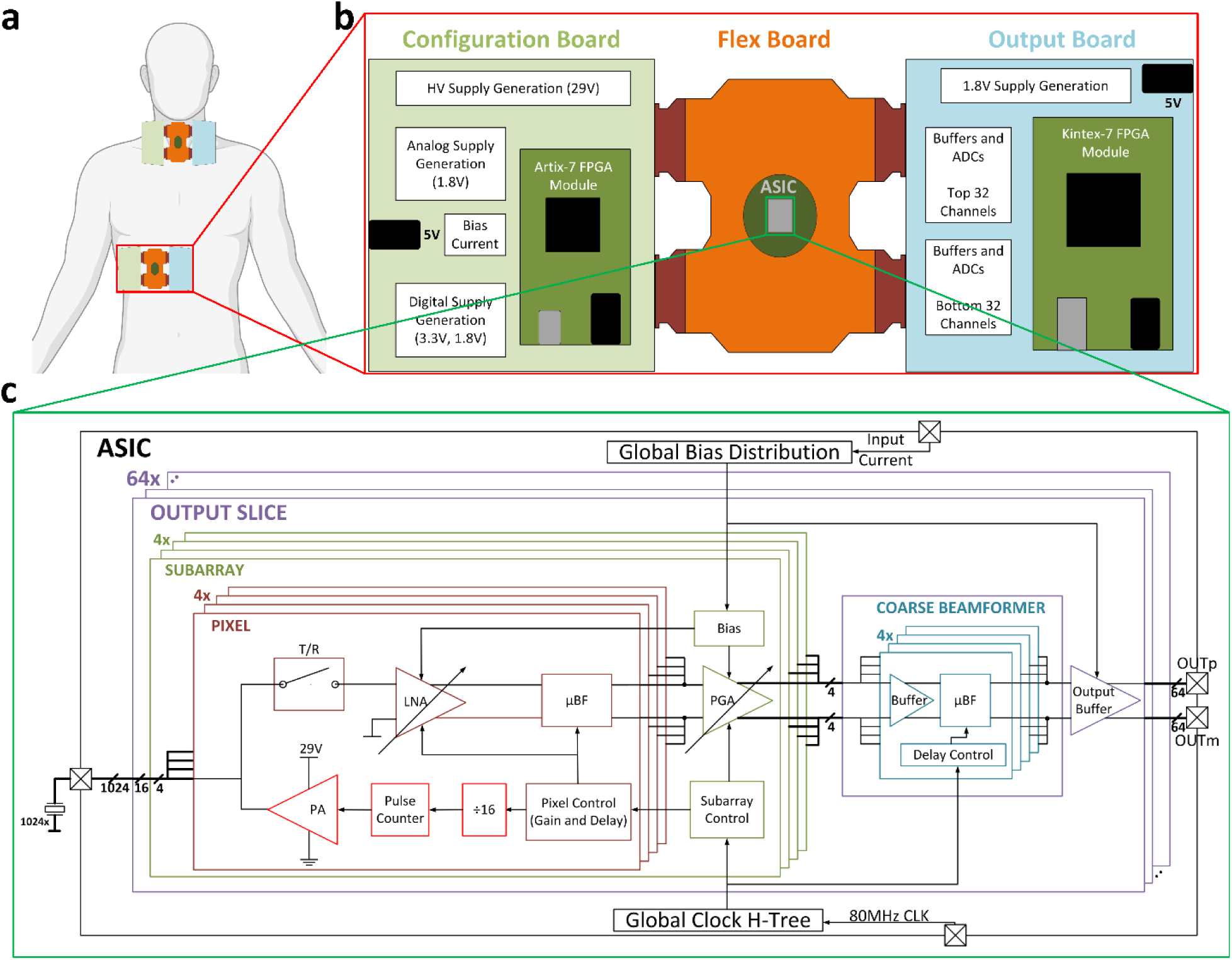
FlexUS System Overview. **a,** Target application of the FlexUS system with multiple patches placed on the subject for simultaneous and chronic observation **b,** System block diagram including the configuration board, flex-board with the ASIC, and the output board **c,** ASIC block-diagram detailing the architecture

## RESULTS

### CMOS ASIC Design

The 10 mm ×8 mm ASIC includes 1024 channels in a 32×32 array that is pitch-matched to 205 μm for transducer integration, two-thirds the wavelength of ultrasound in tissue at the 5-MHz operating frequency. Typically, pitch-matched systems aim for half-wavelength center-to-center spacing between elements for a full ±90° steering. However, for the target applications considered here, more than ±30° steering is not necessary, allowing for larger transducer pitch since grating lobes will be outside the field-of-view (FOV) with this restricted steering. The array is divided into sub-apertures, or sub-arrays (**Fig. 1c**), each consisting of a 2×2 cluster of pixels. Each pixel contains high-voltage (HV) transmit circuitry, a low-voltage receiver, and pixel controller. The ASIC operates from three supply voltages, 29 V, 1.8 V, and 3.3 V. The latter is used only for the off-chip digital interfaces. For each of the 1024 transducer elements, the bottom plate is connected to the transceiver while the top plate is biased to 0.9 V.

Micro-beamformers (μBFs) in the form of delay-and-sum blocks are used to compress the 1024 channels on-chip to 64 output channels without the loss of any channel information. Fine μBFs combine the output of the four pixels in each sub-array. Outputs of four such sub-arrays are subsequently combined in coarse μBFs to reduce the channel count from 1024 to 64, effectively generating a single output for every 4×4 cluster of pixels (see **Supplementary Discussion S1**). Delays between 12.5 ns and 387.5 ns can be set in steps of 12.5 ns, i.e., at the period of the 80-MHz system clock frequency.

For the transmitter (TX), digital counters are used to generate the 5-MHz carrier from the 80-MHz system clock with a programmable delay for the start of the TX pulse. These delays allow phased-array functionality for the entire array, including the ability to support plane waves or focused or diverging beams on transmit. The trailing edge of the TX pulse is generated by a programmable pulse-width counter. This TX signal is level-shifted to 5V before driving the input of the high-voltage (HV) 29-V TX power amplifier (PA), which in turn directly drives the pixel-level piezoelectric element. The programmability of the pulse-width allows transmitted power to be traded off against axial resolution with a smaller pulse-width providing the highest resolution. The PA output drives the piezoelectric element at the pixel directly. A transmit-receive (T/R) switch with 32-dB of isolation protects the low voltage (LV) receiver front-end circuitry from the HV transmitter. Feedthrough is the result of coupling between the electrodes and the low-noise amplifier (LNA) feedback capacitors in the receive circuitry that are vertically stacked, but time-multiplexed operation prevents this from affecting imaging performance while providing sufficient isolation and over-voltage protection to the LNAs.

During receive mode, the T/R switch connects the received signals to the LNA in each pixel. This LNA is fully differential with one input connected to the bottom plate voltage of the individual piezoelectric element for that pixel while the other input is connected to the top-plate, which is common for the entire element array (at 0.9 V). The LNA has a band-pass response with poles at 2 MHz and 10 MHz to limit the in-band noise power while maintaining a -3-dB signal bandwidth of 6 MHz, important for maintaining axial resolution. The LNA gain is programmable in steps of 6 dB from 0 dB to 24 dB with the output of the LNA connected to the in-pixel fine µBF, which delays the outputs by pre-programmed amounts (see **Supplementary Discussion S1**). The outputs of all four pixels in a sub-array are summed at the parasitic input capacitance of a programmable gain amplifier (PGA), which has a band-pass response between 1 MHz and 10 MHz, further reducing the output noise, with programmable gain settings of 0 dB, 6 dB and 12 dB. The net result is a signal chain providing an overall programmable gain between 0 dB and 36 dB. The PGA outputs are then driven along the columns of the array to the peripheral circuits, which contain the coarse µBFs. For specifics on circuit implementation and topologies used, see **Supplementary Discussion S2.**

The ASIC is programmed through a scan-chain distributed through the pixel array with each row of the array having its own scan-chain input. This configuration includes enables for each pixel, gain settings for the LNA, and the phase shifter delay settings. These 32 scan inputs allow for fast reconfigurability with the entire array reprogrammable in only 3.6µs. The scanned data is shifted in during the longer receive phase and then latched during the transmit phase, eliminating the dead-time between successive transmit and receive cycles (see **Supplementary Figure S1**).

The achievable frame rate is dependent on the required depth of the desired FOV and the number of transmit and receive acquisitions required per frame for spatial sampling. For the imaging results presented here, the maximum depth of interest is 55 mm giving a maximum pulse repetition frequency of 14 kHz, for which each focal zone or synthetic focus requires a reconfiguration of the array through the scan chains. The ASIC consumes an average power of 1.2W when imaging at 7 fps with 32 evenly-spaced raylines per ±30° sector, or approximately 1.16mW/channel, and has an SNR of 57 dB with an input-referred noise density of 11nV/√Hz and a dynamic range (DR) of 88dB for the measurement channel electronics. The overall front-end has a band-pass response between 2 MHz and 8 MHz. Across the entire ASIC, the phase variation is less than one least-significant-bit (LSB) or 12.5 ns, including the effect of varying gain on phase, and overall gain variation is less than 0.6 dB across channels, eliminating the need for complex calibration approaches (see **Supplementary Discussion S3**).

The 64 outputs of the ASIC are buffered and digitized off-chip on the supporting printed-circuit boards (**Fig. 1b**) with the outputs sampled at 20 MSamples/s. The data stream into a field-programmable gate array (FPGA), are buffered, and are then output to a host computer over USB3.0 after a full frame is received with direct-memory-access-(DMA-)like transfer from DRAM. Each rayline produces 1536 10-bit samples at the 20 MSamples/s sample rate for a 77-µs sampling interval, resulting in an effective aggregate transfer rate from the ASIC of 12.8 Gbps across 64 channels. At 32 raylines per frame (125.9 Mb/frame), this could produce a frame rate of more than 400 frames/sec. In the current implementation, however, data transfer rates are limited by what is achievable over the USB3.0 interface to approximately 881 Mbps, reducing the achievable frame rate to 7 frames/sec, which is the highest employed here. The supporting boards also provide the voltage supplies, current bias and scan-chain inputs to the ASIC to program it for a specific transmit beam pattern. The functions and implementations of these supporting electronics are detailed in **Methods** and **Supplementary Discussion S4**.

### Packaging and system design

Monolithic integration of the piezoelectric transducers and the packaging of the system are important to achieving both mechanical conformability and acoustic performance. In this packaging flow, the aluminum pads of the ASIC (**Fig. 2b**) are first electrolessly plated with nickel-palladium-gold (ENEPIG) to build-up these electrodes from 1.6 µm below the level of passivation to 6 µm above (**see Supplementary Discussion S5**). The ASIC, which is thinned to a total thickness of 50 µm (see **Methods**), is wire-bonded to a flexible polyimide PCB, 200-μm-thick, that is further backed by 700-µm thick FR4 directly below the ASIC pad-frame.

**Figure 2.**
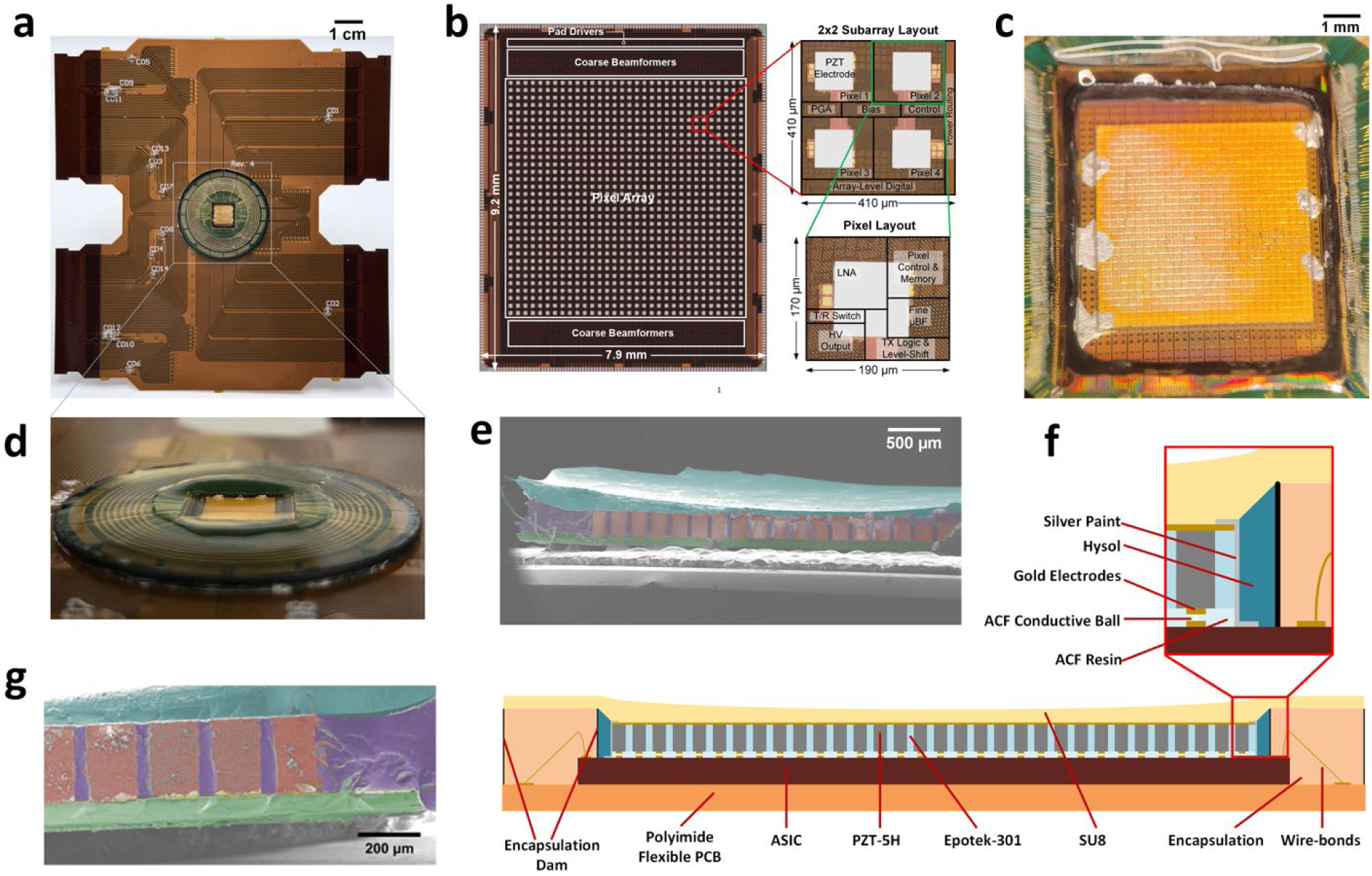
FlexUS System Assembly and Packaging. **a**, Fully-assembled FlexUS device with ASIC, flexible polyimide substrate, piezocomposite, and matching layer **b**, ASIC die-photo with annotations for sub-arrays, pixels, coarse beamformers and pad-drivers **c**, ACF-bonded piezocomposite with silver paint connections to the top-contact **d,** Concave profile of the SU8 matching layer **e,** Low-magnification SEM image of the fully assembled FlexUS device **f,** Full device stack-up including all materials in the acoustic signal path **g,** SEM image of the edge of the composite array with the ASIC (green), the Epotek-301 in the kerfs between the pixels (purple), the ACF (yellow) under the piezoelectric pillars (red), the hysol on the edge of the array (also purple on the right), and the matching layer (light blue).

Beyond flexion, the thinning of the ASIC further serves to improve the acoustic backing of the stack-up^20–23^. In the absence of thinning, the high acoustic impedance of the silicon ASIC would effectively shift the resonance from half-wavelength mode towards the quarter-wavelength mode^27^, while having a significant effect on the system’s bandwidth. Thinning the silicon to thicknesses below those of the polyimide substrate, however, minimizes these backing effects, while enabling the lower-impedance backing presented by the polyimide and the lossy backing presented by the FR4 to reduce the quality factor of the resulting acoustic stack, improving FBW and axial resolution. The stack’s flexural rigidity is dominated by this 700-µm FR4 backing layer, whose per-unit-width plate rigidity *D = Et³/[12(1−ν²)]* ≈ 0.64 N·m (here *D* is the plate flexural rigidity per unit width, *E* ≈ 22 GPa is the in-plane Young’s modulus of the FR4, *t* is the layer thickness, and ν ≈ 0.14 is Poisson’s ratio) exceeds that of the PZT composite by ∼23× and that of the polyimide PCB and thinned silicon ASIC by ∼330×. This FR4 is stiff enough to provide a defined acoustic backing, keeping the ASIC die from flexing too much at the wirebonds while keeping it compliant enough to wrap around a ∼3–5 cm radius; the wirebond connections between the ASIC and the flexible PCB limit the achievable bend radius of curvature to approximately 3.05 cm in concave mode and 4.83 cm in convex mode (see **Supplementary Figure S2**). The combined polyimide-FR4 also includes additional thermal vias and metal to conduct heat away from the face of the ASIC. These metal features, however, are explicitly excluded from the region directly under the array itself to prevent the formation of acoustic resonant cavities. In contact with the body, steady-state heating is limited to 40°C in continuous operation.

The transducers are a 190-μm-thick 3,1-piezocomposite of PZT-5H with Epotek-301 used as the kerf-filler material (see **Methods**). The addition of a lower density and lower acoustic impedance epoxy reduces the overall acoustic impedance of the composite material, bringing it closer to that of the tissue^28^. In addition, composites have lower Q than raw piezoelectrics, increasing both mechanical flexibility and transmit efficiency^29^. The thickness, chosen based on Krimholtz, Leedom, and Matthaei (KLM)^30^ model simulations (see **Supplementary Figure S3**) of the acoustic stack-up, is less than the half-wavelength thickness (∼290 μm) of the composite PZT-5H for vertical vibration modes (bar mode). The element widths and lengths are 155 µm with a kerf width of 50 µm, allowing FlexUS to be pliable while maintaining as large an element size as possible for maximum pressure^31^.

The composite, fabricated through a dice-and-fill process, is bonded to the ASIC using an anisotropic conductive film (ACF) (see **Methods**). The pressure required during the bonding process to achieve the minimum resistance for this connection is determined with repeated plane-wave echo tests (see **Methods** and **Supplementary Figure S4**). The top electrode contact is then made using conductive silver paint to pads on the ASIC. The doughnut epoxy encapsulation of the wirebonds is sealed internally with a low-acoustic-impedance sealant to prevent lateral mechanical reflections (**Fig. 2c**) and the entire well is spin-coated with a 150-µm-thick layer of SU-8 (see **Methods**), which acts as a matching layer^32,33^. The thickness of the SU-8 increases from the center to the periphery (**Fig. 2d**), the effect of which is less than a single period of the 20-MHz sampling clock across the entire array for any steering direction and does not warrant lens corrections (see **Supplementary Discussion S6**). The tight clearance between the wirebond encapsulation and the composite provided insufficient space for the connections to the top electrode contact. The device employed here for imaging, therefore, bonded only a 30×30 composite array, for which only the inner 28×28 elements were electrically connected. A cross section of a fully-assembled FlexUS device (**Fig. 2e, g**), imaged using a scanning electron microscope (SEM, see **Methods**), confirms the concave profile of the SU8 matching layer and expected thicknesses in the stack-up (**Fig. 2f**). The zoomed-in SEM image of the device edge (**Fig. 2g**) shows all the constituent elements of this stack-up.

### Acoustic characterization

The basic transmit and receive characteristics of the array are first characterized. The patch is capable of the three most common transmitter modalities – focused wave, plane wave, and diverging wave, including the full range of steering angles from -30° to +30° in the azimuth (**Fig. 3a**) and from -15° to +15° in the elevation (orthogonal to azimuth planes). The focal depth for transmit, i.e. the Rayleigh or far-field distance, is limited by the aperture (5.74 mm × 5.74 mm) and the excitation frequency (5 MHz) to approximately 26 mm. The transmitted pressure profile for focused waves at 20-mm focal depth (**Fig. 3b**), for plane waves (**Fig. 3c**), and for diverging waves with a 50-mm radius of divergence at the aperture (**Fig. 3d**) are measured in both the XZ and XY planes. As determined by the full-width-at-half-maximum (FWHM) relative to the peak pressure in the XY plane, the lateral resolution for these is estimated to be 1.5 mm, 2 mm and 2.4 mm, respectively.

**Figure 3.**
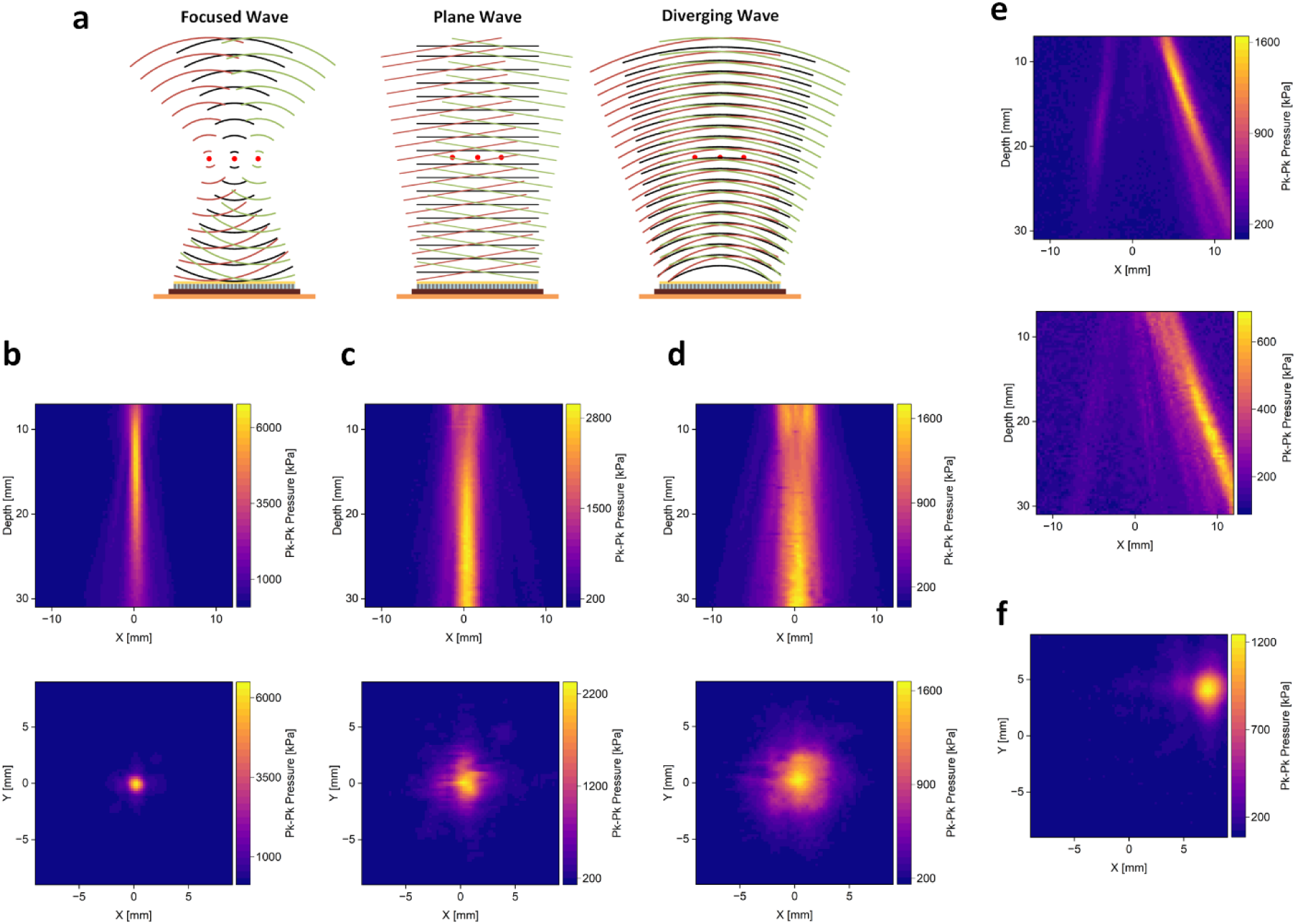
FlexUS beamforming modalities. **a,** Different beamforming modalities that can be generated by the FlexUS device. **b,** Device pressure profile in the XZ and XY planes for a focused beam with 20 mm focus and 0° steering. **c,** Pressure profile in the XZ and XY planes for a plane wave with 0° steering. **d,** Pressure profile in the XZ and XY planes for a divergent beam with –50 mm focus and 0° steering. **e,** Pressure profile in the XZ planes for a focused beam with 20 mm focus (top) and a plane wave (bottom), both with 22.5° steering. **f,** Pressure profile in the XY planes for a plane wave and 15° steering in azimuth and 11.25° steering in elevation.

Steered pressure profiles for 22.5° azimuth are also measured for 20-mm-depth focused waves and plane waves (**Fig. 3e**) to characterize the ability of the aperture to steer energy away from the normal direction. The peak pressure for the 20-mm-depth focused wave is almost 7 MPa (peak-to-peak), while that for the steered focused wave, plane wave and steered plane wave are 1.65 MPa, 3 MPa and 0.7 MPa, respectively. The drop in pressure for steered beams results from the reduction in contribution from elements in the array that have a larger distance to the focal point due to the inverse-square relationship between pressure and distance. Moreover, the directivity of the individual elements is not spherical and will also affect the pressure in non-perpendicular directions. Beams can also be steered in both azimuth and elevation (**Fig. 3f**), with all profiles being symmetric in azimuth and elevation. A peak negative pressure of 3.5 MPa corresponds to a mechanical index (MI) of 1.7, which is below the limit of 2.0 for medical ultrasound. A grating-lobe is visible in the profile for the steered focused wave (**Fig. 3e**) with an amplitude one-quarter of the peak pressure.

Pulse waveforms were also analyzed (see **Supplementary Figure S6**) to determine the level of control the ASIC has on pulse duration, with the device capable of generating pulses of the commanded width with a resolution of 500 µm — a prerequisite for fine axial resolution, since axial resolution scales with pulse-width. The transmit efficiency per channel based on the peak-to-peak pressure of the 20-mm-focused wave at the focal point is approximately 300 Pa/V (obtained by dividing the focal point pressure by the total number of elements and applied voltage at each element); the 6-dB one-way fractional bandwidth (see **Methods**) is 44%, and the 6-dB two-way fractional bandwidth is 34%. This fractional bandwidth is quite comparable to that reported for purely passive PZT-based conformable devices^14,16,17^. The minimum-detectable pressure (MDP) for the 44% fractional bandwidth at an SNR of 0 dB is measured at 62 Pa_rms_, and the noise-equivalent pressure (NEP) is measured to be 45 mPa_rms_/√Hz (see **Methods**), both on par with rigid catheter-form-factor ASIC-based ultrasound devices^34,35^. These receiver-noise-floor benchmarks are set primarily by the front-end electronics rather than the acoustic stack-up.

### Phantom imaging

A 3D-printed water tank is affixed onto the FlexUS imager with four 250-µm-diameter metal wires placed in an orientation such that when imaging from the XZ plane, all four wires are visible as point targets, while YZ plane imaging shows the wires along their lengths (**Fig. 4a**). Imaging is done in one of two orientations: XZ-centric and YZ-centric, which define the azimuth and elevational direction (see **Supplementary Figure S7**). The center of the array defines the origin (X=0, Y=0, Z=0) with all wire phantom images obtained using 32 evenly-spaced scan lines using a 3-MPa plane wave excitation at 5 MHz for each scan line and the minimum possible transmit pulse-width of 100 ns. Reconstruction is performed using delay-and-sum (DAS) beamforming^36^ (see **Methods** and **Supplementary Discussion S7**) on a 128 × 128 voxel grid.

**Figure 4.**
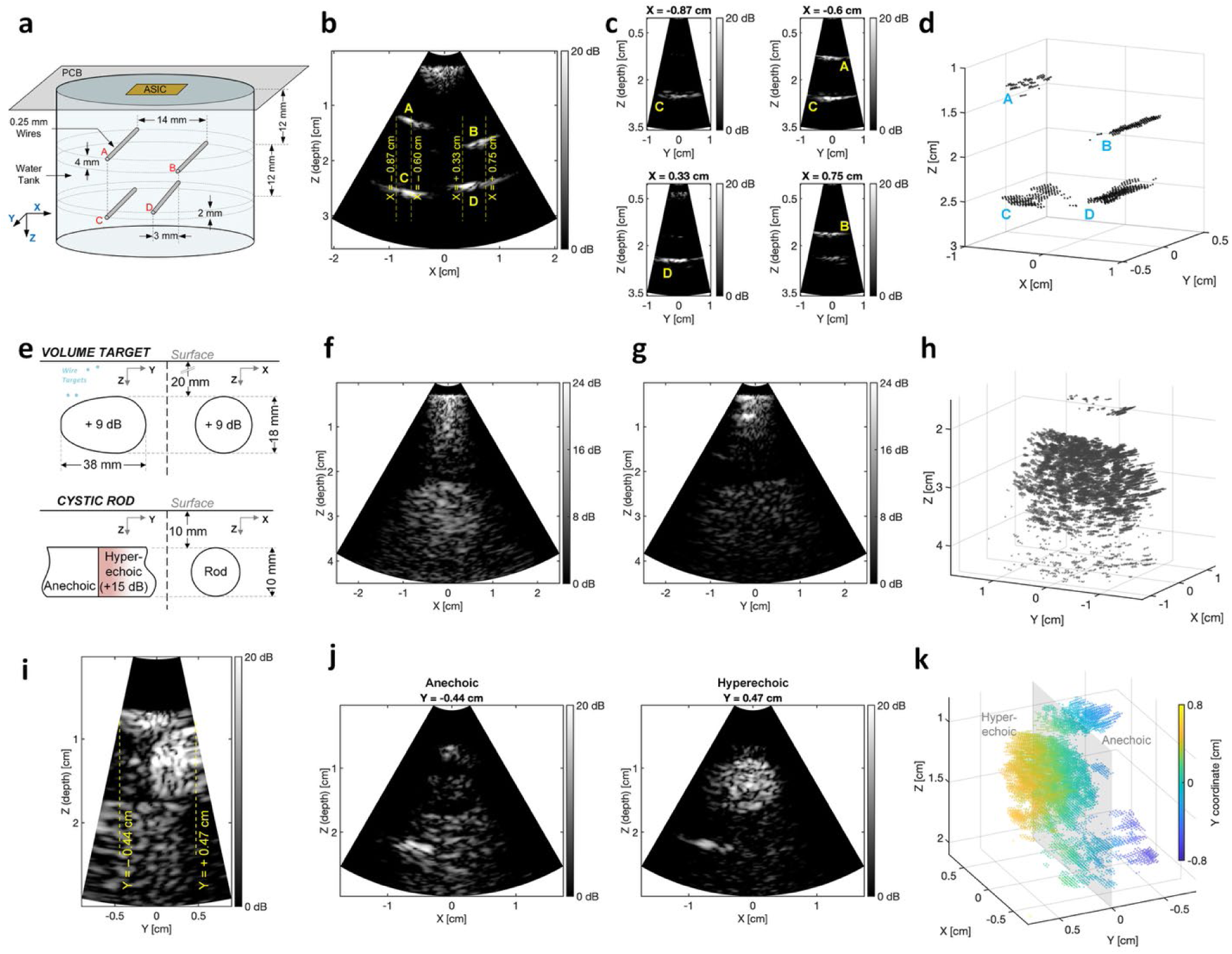
Phantom experiments with FlexUS. **a,** Wire locations relative to the patch in the custom wire-phantom **b,** XZ cross-section image of the wire-phantom with wires and YZ slice locations annotated **c,** YZ slice locations at X=−0.87 cm, X=−0.6 cm, X=0.33 cm and X=0.75 cm from top-left to bottom-right **d,** 3D rendering of the wire-phantom **e,** Phantom volume and rod object locations, dimensions and acoustic characteristics **f,** XZ (short-axis) cross-section of the volume object **g,** YZ (long-axis) cross-section of the volume object **h,** 3D rendering of the volume object **i,** Limited-sector YZ (long-axis) cross-section of the rod object between hyper-echoic and anechoic regions with XZ slice locations annotated **j,** XZ (short-axis) cross-sections of the anechoic (Y=-0.44 cm) and hyper-echoic (Y=0.47 cm) rod object **k,** 3D rendering of the rod object with both hyper-echoic and anechoic regions visible

For the XZ-centric images, sectors are defined by the ±30° azimuthal angle. **Fig. 4b** shows the XZ-centric image of the wire phantom along the Y=0 plane with axial and lateral resolutions (from FWHM) estimated to be 0.5 mm and 2 mm, respectively. The locations of the wires in the image are in agreement with their locations in **Fig. 4a**: wires A, B, C and D in the B-mode image are at depths of 12.8 mm, 16.6 mm, 26 mm and 24.5 mm respectively, and the horizontal separation between wires A and B is 13.2 mm while that between wires C and D is 10.5 mm.

YZ-centric images are then reconstructed along orthogonal slices (**Fig. 4c**) at the X coordinates indicated by vertical lines in **Fig. 4b**. Sectors in this case are defined by the ±15° elevantional angles. Going from negative to positive X, Wire C appears at X=−0.87 cm with Wire A coming into view next at X=−0.6 cm along with Wire C. Wire D then appears at X=0.33 cm with Wire B appearing at X=0.75 cm. The wires are at the same depths as in **Fig. 4b**, and are present across the full length of the sector because imaging is along their length in this case. A full 3D reconstruction of the wires was created with amplitude thresholding (see **Methods**) and is shown in **Fig. 4d**. Wires C and D are characterized by poorer resolution than Wire A and B as they are more distant.

We then used the FlexUS imager with a custom ultrasound phantom (see **Methods**) made of a material matched to the speed of sound (1540 m/s) and attenuation (0.5 dB/cm/MHz) of tissue. Embedded within this tissue phantom is a 10-mm-diameter rod across the full width of the phantom that has hyper-echoic and anechoic halves and an ovoid object with 18-mm diameter, 39-mm length, and 9 dB of acoustic contrast at a 30-mm depth (**Fig. 4e**). Images are generated using steered-plane-wave excitation with a pulse-width of 400 ns, and the reconstructions use a focal ratio (F-number)^36^ of one in both X and Y.

**Fig. 4f** shows the imaging of the ovoid object in the phantom along the XZ-plane with a rough diameter of 16mm and a contrast of approximately 10 dB. The same image is plotted along the YZ plane (**Fig. 4g**) with the longer axis of the volume being seen. The patch FOV at the 30-mm depth of this object is smaller than the 39-mm length of the object; as a result, the object appears roughly rectangular and occupies the full FOV. The YZ-centric slices are reconstructed in 3D in **Fig. 4h**.

For the rod object in the phantom, **Fig. 4e** shows both the hyperechoic and anechoic regions with the rod’s cross-section aligned to the patch’s XZ plane. In **Fig. 4i**, an XZ-centric 3D acquisition is run and the YZ plane is plot on an orthogonal grid. The YZ plane is along the length of the rod, with the left of the image being the anechoic region and the right being the hyperechoic region. Along the designated Y slices, one in the anechoic region and one in the hyperechoic region, the XZ-plane images are shown in **Fig. 4j**. The hyperechoic region appears as a clear 9-mm-diameter circle with 15-dB of contrast. However, only the top and bottom interfaces of the anechoic region are visible with higher speckle in the shadow below the rod. Very little pressure reaches this region with the gain control of the FlexUS imager amplifying the lower SNR received signals from below the rod. The sides of the rod are not visible since the transmitted pressure at none of the steering angles is directly incident on the sides without first passing through the top interface of the rod. The 3D reconstruction of the rod is shown in **Fig. 4k** with the hyperechoic region now at the left. The larger speckle below the anechoic region is, yet again, visible with the top and bottom interfaces of the anechoic region seen to the right of the hyper-echoic region.

### Human imaging

For human imaging, we began by placing FlexUS on the neck to image the internal jugular vein (IJV) and carotid artery (CA) (see **Supplementary Figure S8**). To do this, FlexUS is placed with its x-axis (XZ imaging plane) along the length of the vein and flexed towards the lateral side of the neck (**Fig. 5a**). A Philips medical ultrasound system with an S5-1 phased array at 5-MHz excitation frequency is used to image the same side of the neck and independently locate the IJV and carotid artery (**Fig. 5b**). The optimum location that allows the viewing of both vessels, despite the anatomic variants in the subject, is used in subsequent images. The commercial array is placed approximately 5-mm from the skin surface with a gel interface, and the top of the IJV starts at a depth of 2 cm, with the carotid visible directly beneath the IJV at 3 cm. When rotated 90°, the long-axis of the vessels is observed (**Fig. 5c**). Guided by these images, the FlexUS device is placed along the long-axis of the IJV. The device conforms to the neck without the need for coupling gel. Signals are acquired at a frame rate of 5 frames/sec.

**Figure 5.**
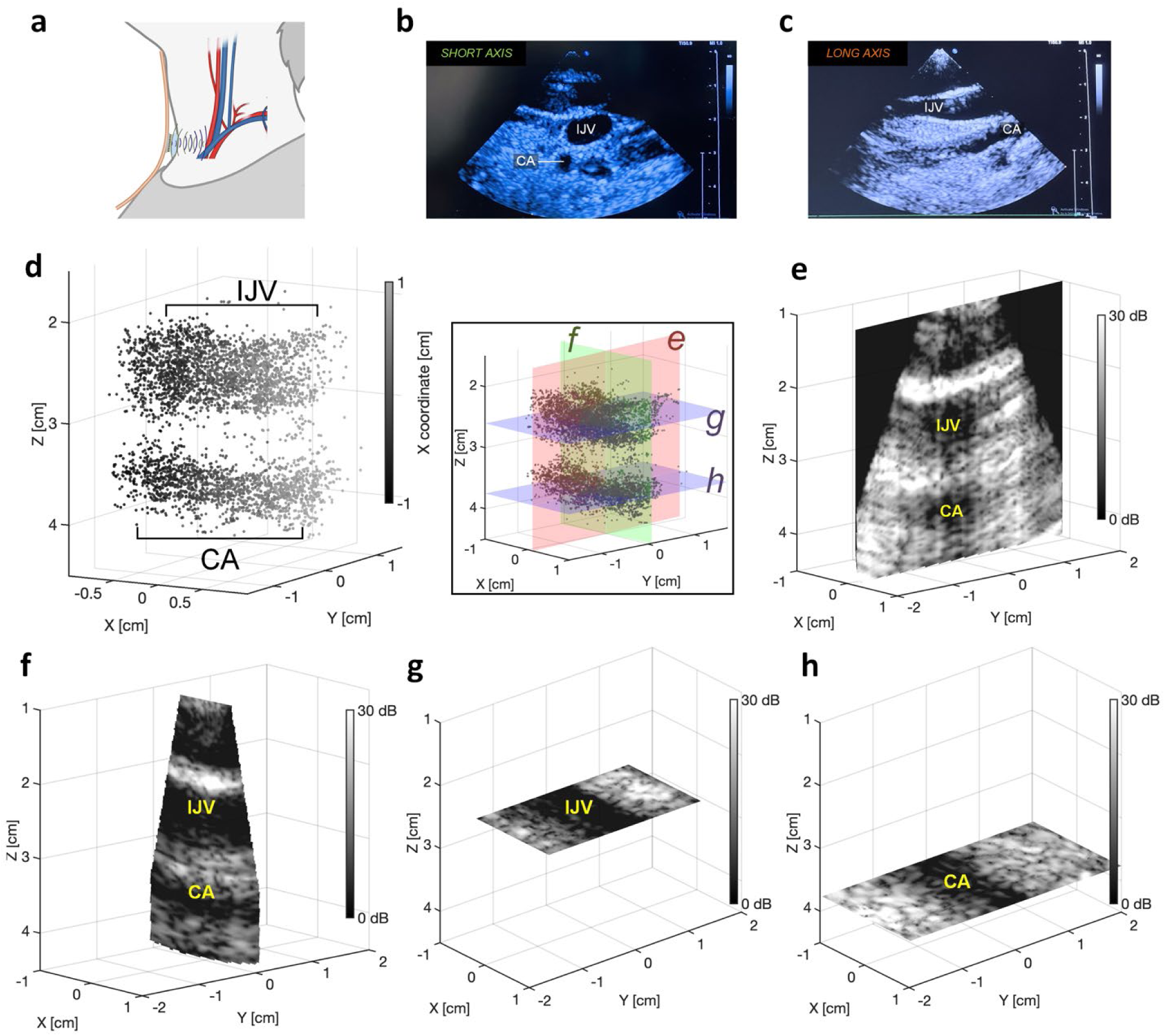
Imaging in the neck with FlexUS. **a,** Application of the patch for observing and guiding CVC placement **b,** Short-axis cross-section of the IJV and carotid artery from Philips S5-1 probe **c,** Long-axis cross-section of the IJV and CA from Philips S5-1 probe **d,** Inverted-brightness 3D rendering of the neck viewed from the long-axis (XZ) plane. The points in the plot represent least speckle in a standard B-mode image, with the inset showing the reconstruction planes for 2D slices in **e-h e,** 2D B-mode image reconstructed along the short-axis of the IJV and CA at -4.5° elevation **f,** 2D B-mode image reconstructed along the long-axis of the IJV and CA at -4.5° azimuth with narrower FOV due to orthogonal reconstruction **g,** 2D B-mode image reconstructed on the XY plane (parallel to skin) at Z = 2.6 cm showing only IJV **h,** 2D B-mode image reconstructed on the XY plane (parallel to skin) at Z = 3.75 cm showing only CA

For the device’s orientation, more negative X positions correspond to lower positions on the neck. A YZ-centric three-dimensional acquisition was performed and after reconstruction, the brightness magnitude of the regions corresponding to the blood vessels was found to be ∼25 dB lower than the surrounding tissue. A 3D image of the IJV and CA (**Fig. 5d**) is generated by only displaying those points in the reconstruction grid whose brightness corresponds to this lower brightness level (see **Methods**). This visualization method was chosen instead of the standard approach of displaying higher-brightness points to highlight the ability of the device to map vasculature in 3D and to aid the visibility of the lower-speckle vasculature against the bright speckle in the surrounding tissue. The IJV is visible at a depth of 2.6 cm and the CA at a depth of 3.75 cm, with the diameter of the latter (approximately 6 mm) being roughly half that of the former (approximately 10 mm). Moreover, the vessels are not parallel to each other, with the CA approximately parallel to the X-axis and the IJV in the direction +4.5° from the X-axis.

Four different planes were chosen to display 2D slices as labelled in the inset image in **Fig. 5d** and these slices are displayed in **Figs. 5e-h**. The 2D slices are physically oriented along the corresponding planes in 3D space to aid in visualization. The planes in **Fig 5e** and **Fig. 5f** are chosen so that they are perpendicular (short-axis) and parallel (long-axis) to the direction of the IJV, that is, their normals are each +4.5° from the Y- and X-axes, respectively. Accordingly, the cross sections of the IJV (larger diameter, at depth 2.6 cm) and CA (smaller diameter, at depth 3.75 cm) are visible in **Fig. 5e**. The longitudinal sections of the IJV and CA are visible in **Fig. 5f** at the same depths. There is considerably more speckle below the IJV because of the significant absorption of ultrasound energy by the blood of the IJV. The carotid also shows higher speckle due to the higher flow rate; pulsation in both vessels is similar to the frame rate in these experiments, leading to additional noise aliasing. The reconstruction planes in **Fig. 5g** and **Fig. 5h** are parallel to the XY plane and are located at Z = 2.6 cm and Z = 3.75 cm which show sections of the IJV and CA along the plane roughly parallel to the skin, further highlighting the 3D imaging capabilities of the FlexUS device.

We next positioned the FlexUS device in the right second intercostal space overlying the pectoralis muscle (where needle thoracostomy is emergently performed) to minimize the distance to the pleural surface (see **Supplementary Figure S9**). The reference images from along the YZ plane (parasagittal slice of the lung) and XZ plane (transverse slice of the lung, in between the ribs) are acquired with the Philips S5-1 phased array at 5 MHz (**Fig. 6b**). As a healthy lung expands and contracts during respiration, the visceral pleura moves against the parietal pleura causing shimmering, time-varying speckle patterns along the pleural line in a B-mode ultrasound image, known as lung sliding^7^. However, in cases of air present in between these layers, a condition known as pneumothorax, lung sliding is not observed indicating that the pleural layers do not move in opposition to each other, and making lung sliding a useful tool in evaluation of pulmonary health^37^ and respiration rate. The B-mode slice needs to be parallel to the sagittal axis of the human body in order to be along the direction of lung movement, which in the case of the positioned Philips S5-1 device is the YZ direction, making this the more important axis for detecting lung sliding. In the YZ reference image (**Fig. 6b**), the ribs can be seen on the edges with large shadows below them and the intercostal tissue directly between them. The two pleural walls can be seen at depths of around 2.2 cm as bright arcs between the edges of both ribs.

**Figure 6.**
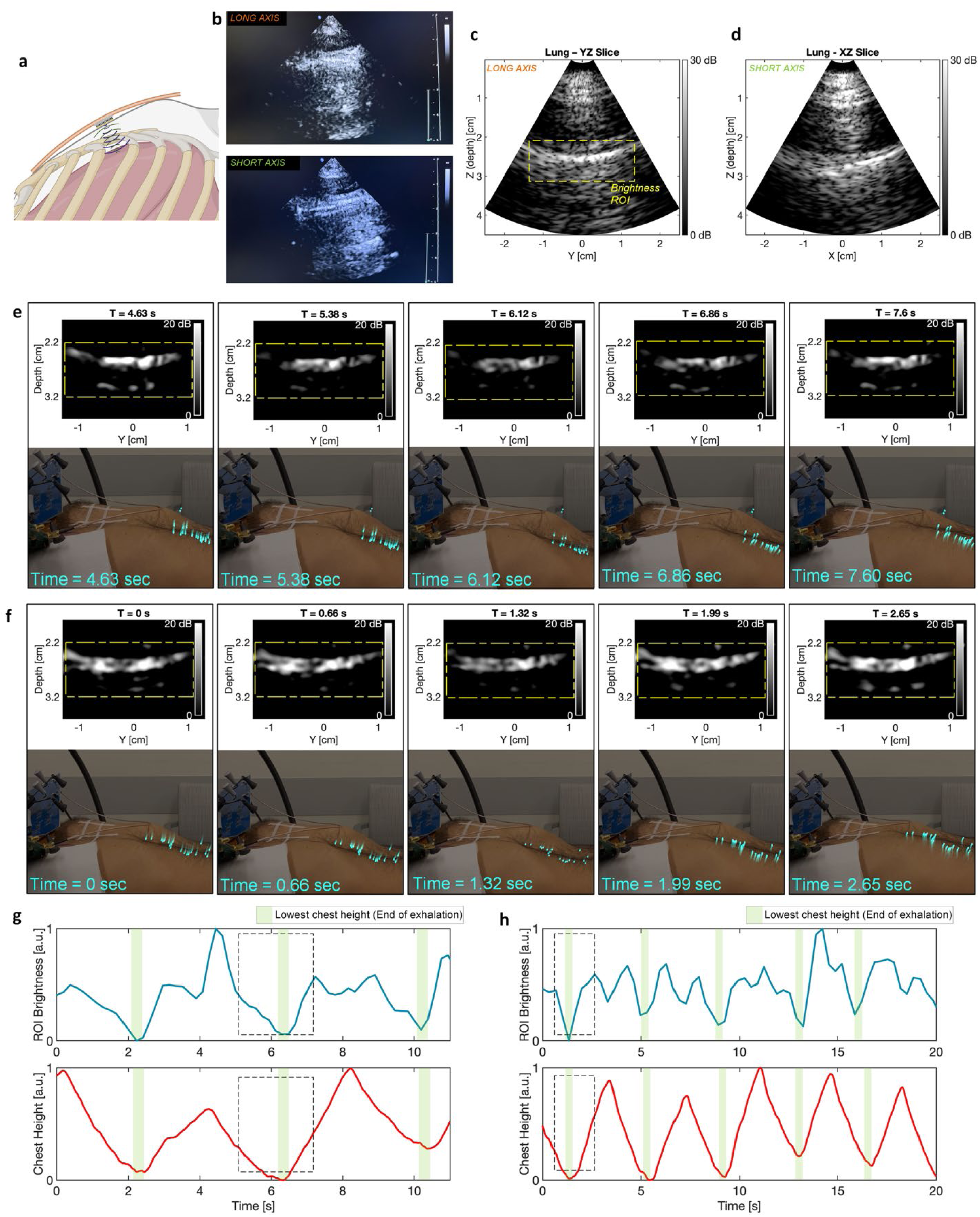
Lung imaging with FlexUS. **a,** Application of the patch for diagnosing pneumothorax **b,** Parasagittal (perpendicular to the ribs, top) and transverse (parallel to the ribs, bottom), slices of the right lung from Philips S5-1 probe **c,** Parasagittal slice reconstruction of the right lung using the FlexUS device **d,** Transverse slice reconstruction of the right lung using the FlexUS device **e,** Frame-by-frame snapshots of the Gaussian-blurred ROI overlaid on recording of the subject’s chest motion for one exhalation cycle (as indicated in Fig. 6g) for the higher frame rate acquisition. The blue dots represent the chest height tracking points and their tails represent the velocity of these points **f,** Frame-by-frame snapshots of the Gaussian-blurred ROI overlaid on recording of the subject’s chest motion for one exhalation cycle (as indicated in Fig. 6h) for the lower frame rate acquisition. The blue dots represent the chest height tracking points and their tails represent the velocity of these points **g,** ROI brightness-sum and chest displacement plot against time with the three highlighted regions showing end-of-exhalation for the higher frame rate acquisition **h,** ROI brightness-sum and chest displacement plot against time with the five highlighted regions showing end-of-exhalation for the lower frame rate acquisition

The FlexUS device was positioned with its X-axis aligned parallel to the ribs and its superior aspect directed caudally. In this orientation, the positive Y-direction corresponds to movement inferiorly along the subject, and the positive X-direction corresponds to movement medially toward the sternum. The XZ/transverse plane (**Fig. 6d**) is used primarily for localization, with the YZ/parasagittal plane (**Fig. 6c**) being used for characterizing lung-sliding. The transverse slice was imaged at 4 MHz using a steered plane wave with a pulse-width 400ns, while the parasagittal slice was imaged at 5 MHz using the same transmit pattern and pulse-width. The transverse slice shows the diagonal intercostal tissue seen in the gold-standard image, with the pleural wall being the lower horizontal interface at the correct depth of 2.2 cm. The parasagittal slice only shows the pleural wall, confirming that the patch imaging aperture fits between two ribs. The shadowing from the ribs is slightly visible at the bottom left and bottom right edges of the FOV.

Real-time 2D frames were then acquired at a frame rate of 5.4 fps, with the subject taking slow deep breaths over the course of recording. An M-mode equivalent is created by condensing the center region of the B-Mode frame into a single brightness line for each time stamp (see **Methods**), and is then plot against time (see **Supplementary Figure 10a**). This shows the presence of a sea-shore sign^37^, which consists of a bright horizontal line that corresponds to the pleural line (at 2.6 cm), several parallel lines (waves) above the pleural line that correspond to the static tissue layers (such as skin and muscle) and random speckle patterns below the pleural line (sand) that correspond to the time-varying speckle patterns that arise from lung sliding. The presence of the ‘sand’ artefact below the pleural line confirms lung sliding in the subject. In addition, artefacts called “A-lines”^37^ can be observed in the M-mode images at a depth of 4 cm. A-lines are bright lines that appear parallel to and under the pleura and are caused by repeated reverberations of sound waves between air in the lungs and the skin/transducer, and are roughly the same distance from the pleural wall as the pleural wall is from the transducer.

To confirm that the brightness variation observed in the B-mode and M-mode images correspond to actual lung motion and to extract the breathing rate from this variation, a brightness metric for the region of interest (ROI) surrounding the pleural region was devised and compared against video footage of the subject’s chest movements (see **Methods**). Moreover, the chest height of the subject was also tracked (see **Methods**), with the tracked points being displayed over the video of the subject’s respiration (see **Supplementary Movie S1**) and programmed to fade out after each frame, resulting in a composite video where the tail behind the tracked points in the video is representative of the velocity of the points. The ROI brightness-sum and chest height for the same acquisition as the sea-shore plot (see **Supplementary Figure 10a**) are plot side-by-side for comparison (**Fig. 6g**), and frame-by-frame snapshots of the B-mode image overlaid on the subject’s chest movement are shown for select frames in one exhalation cycle (**Fig. 6e**). It can be observed that the minima in the brightness signal correspond to the end of the exhalation phase (lowest chest height) in each breathing cycle. There are also minima corresponding to the end of the inhalation phase (highest chest height) but these are not as strongly correlated. The dip in brightness at the end of each phase occurs due to the momentary pause in pleural sliding as the direction of lung movement changes from inferior to superior (or otherwise).

The breathing rate based on the chest motion and ROI brightness-sum is 0.25 Hz or one deep breath every 4 seconds. The acquisition is repeated with a slower frame rate (3 fps) to include five full inhales and exhales instead of the three full breaths in the 5.4 fps recording, with the corresponding sea-shore plot (see **Supplementary Figure 10b**), brightness-sum and chest height across time plot (**Fig. 6h**), and the frame-by-frame snapshots of the B-mode frames overlaid on the chest movement (**Fig. 6f** and **Supplementary Movie S2**) confirming the above observations regarding lung sliding and the 0.25 Hz breathing rate and further demonstrating the use case of the FlexUS patch as a gauge of pulmonary health.

## CONCLUSIONS

This work presents FlexUS, a conformable patch-based point-of-care ultrasound imager for chronic monitoring and hands-free procedural guidance that contains integrated electronics in the form of a custom integrated circuit with pitch-matched transceiver electronics and monolithically integrated transducers. This architecture allows for future scalability to larger arrays if power and data transfer bottlenecks can be addressed. These, when developed, will ultimately lead to wearable 2D arrays have the potential to take POCUS one step further by eventually eliminating the need for a physician to be physically present for the evaluation of the patient, which could be invaluable in certain settings such as combat casualty care or at-home monitoring.

The ASIC that is central to the FlexUS design is capable of both transmit and received beamforming for a wide range of beam patterns (focused, plane, diverging) and steering directions (-30° to 30°). The use of active on-chip beamforming allows a 16× channel count reduction without information loss while supporting a wide range of gain settings. The conformable packaging provides sufficient heat sinking while providing appropriate acoustic matching and backing to achieve FBWs in excess of 34.5% at the measured 4.4 MHz center frequency. Packaging also includes the integration of piezocomposites to the ASIC, delivering both lower acoustic impedance through kerf filling while maintaining conformability. Employing SU8 as a matching layer improved signal transmission by ∼54% while introducing insignificant delay variation across the array. In phantom imaging, the axial, lateral and elevational resolutions achieved are 0.5 mm, 2 mm and 2 mm, respectively. The former is set by the achieved FBW while the latter is limited by the width of the array aperture. SNR and DR allow contrast as high as 88 dB to be observed. High imaging pressures are also possible with FlexUS, with peak-to-peak pressures as high as 7 MPa with focused waves and up to 3 MPa with plane waves and diverging waves, which correspond to mechanical indices of 1.7 and 0.67 respectively.

FlexUS was used to image the IJV and carotid artery and to observe lung sliding of the visceral pleura against the parietal pleura. Both of these have application in critical care environments. IJV/CA imaging can be used for observing and guiding the insertion of a central venous catheter, which could greatly help reduce the possibility of pneumothorax, arterial puncture and its complications like hematomas, losing a guidewire, catheter malposition, catheter kinking, and wall puncture. Such imaging can also be used for functional hemodynamic monitoring for hypovolemia. Because of the continuous nature of the technology being used, we can increase the sensitivity in identifying bleeding patients by looking at changes over time. Increasing respiratory variation in vein diameter over time can be used as an internal jugular index, identifying volume loss prior to changes in stroke volume and allowing for the quantitatively assessment of hemorrhagic shock^4^. For lung imaging, patients are evaluated for the presence, or more importantly, the absence of pleural sliding to diagnose pneumothorax^6^. As in the results presented here, we can automate the diagnosis of pneumothorax by detecting pleural sliding and lung point using motion tracking in B-mode US images. Movement of the pleural line region can be quantitatively tracked by a discretized grid of points. Using horizontal motion vectors of these points as classification features, a supervised machine learning method can be applied to determine whether the patient has pneumothorax.

Limitations of the current generation patch include the lack of on-chip time-gain control (TGC) to improve SNR at higher depths, a limit on the maximum elevation angles that can be achieved for maximum azimuth, variability in the matching layer profile which could cause non-zero lensing delay variation across the array (currently limited to less than a single cycle of the sampling clock), lack of space for a full 32×32 composite array, and an upper limit in the frame rate of 7 fps when full-volume spatial sampling is required. Further integration of data conversion and data transfer should eliminate the need for additional components currently on the support board. Frame rate improvements will require some level of beamforming or data compression on the FPGA in addition to more complex memory arbiters that allow simultaneous read and write operations to the DRAM without long wait times for the data to be valid. The use of more flexible acoustic backing materials (such as tungsten-loaded epoxies) and non-wirebond connections between the ASIC and the package (such as connections made with solder bumps or anisotropic conductive films) can further improve device conformability. In addition, moving to non-wirebond ASIC integration strategies would help make the matching layer more uniform, since the presence of the wirebonds led to the enclosed geometry of the composite that caused the concave matching layer profile.

Taken together, FlexUS demonstrates that monolithic integration of pitch-matched transceiver electronics with a conformable transducer array can bring beamformed, volumetric ultrasound into a wearable patch without sacrificing image quality. By pairing the channel-count reduction of on-chip beamforming with conformable packaging, FlexUS bridges the gap between rigid ASIC-based probes and passive flexible arrays. As the power and data-transfer bottlenecks identified above are addressed, this architecture points toward a new generation of self-contained, body-conformal imagers capable of continuous, operator-independent point-of-care diagnostics.

## METHODS

### Design and fabrication of the ASIC

The FlexUS ASIC was designed in a custom mixed-signal design flow. The generated GDSII design format was validated against design rules specified by the foundry using Calibre Design Rule Check (nmDRC, Siemens EDA) and Calibre Layout-vs-Schematic (nmLVS, Simens EDA). Further validation of the layout was performed using Calibre Resistance-Capacitance Extraction (xRC, Siemens EDA) to generate netlists that include parasitic RC components to more accurately account for the non-idealities in nanofabricated layouts. The system controller and other digital systems are written in the Verilog hardware description language (HDL) and simulated using Modelsim (Siemens EDA). The netlists are synthesized using Design Compiler (Synopsys), and undergo place and route using Innovus (Cadence). The digital logic is validated at the gate level and the place and route stage using the same test harnesses used for validation in the initial behavioral Verilog to ensure functionality. A top-level simulation is performed using Virtuoso ADE (Cadence) and the Spectre (Cadence) simulation engine for final checking and verification. The integrated circuit is fabricated in a 1.8V-5V-29V 0.18-µm bipolar-CMOS-DMOS (BCD) process by the Taiwan Semiconductor Manufacturing Company (TSMC). Dice are thinned using the X-Prep system (Allied Hi-Tech, USA) to a die thickness of 50 μm.

### Packaging and assembly

The ASIC is assembled on a custom three-level-interconnect polyimide printed-circuit board of thickness 200 µm (EPEC, USA). Chips are attached using Epotek H20E-D silver epoxy and wirebonded to the printed-circuit board.

The piezoelectric composite is then bonded to the ASIC. The PZT-5H piezoelectric composite (CTS Corporation, USA) consists of 155 µm×155 µm pillars of the piezoelectric with the 50 µm kerfs filled with Epotek-301 epoxy. The composites are fabricated as 5 cm×5 cm sheets and are coated with 0.15 µm NiCr and 1.25 µm Au on both sides. The electrode contacts on the ASIC side are defined by spin-coating SPR-220-4.5 photoresist on both sides, soft-baking at 115°C for 2 minutes, exposing using i-line (365 nm) with a 125 mJ/cm^2^ dose, and etching in Au etchant followed by Cr etchant for 7.5 minutes and 2 minutes respectively. The etched composite is cleaned with de-ionized water and the photoresist is stripped by spraying acetone. These substrates are diced in a DISCO DAD3220 dicing saw (DISCO Corporation, Japan) using a DISCO Z09 series blade of thickness 50 µm at 30000 rpm to a size of 6.15 mm×6.15 mm. A film of 8 µm-thick ACF (H&S Hightech, South Korea) with 5 µm gold-coated polymer balls is cut to match the area of an etched and diced composite array and placed with the tackier side down onto the composite. The composite and ACF are then placed onto the chuck of the Finetech Lambda pick-and-place tool (Finetech, Germany) with the stage vacuum on; the ACF is smoothed on the composite surface using the flat side of a carbon-tipped tweezer. The Finetech Lambda arm is brought down (with its vacuum disabled) until it hovers over the ACF. The ACF is then thermally tacked to the composite by heating to 80°C for 30 seconds, securing the ACF to the composite. A copper sheet, cut to the same size as the composite, is then prepared with thermal release tape (Semiconductor Equipment Corporation, USA) attached on its non-foaming side. The ACF-composite is then attached to the copper sheet by adhering the foaming side of the release tape to the side of the piezocomposite that does not contain the ACF. The copper-composite-ACF stack is placed on top of a Gel-Pack (Gel-Pak, USA), ACF side down, with vacuum applied to prevent the ACF from sticking to it.

Picked up by the Finetech Lambda tool, this composite is then positioned over the flex-board with the wire-bonded ASIC, which is held by vacuum onto the stage; the composite is aligned to the electrodes on the ASIC after bringing both the composite electrodes and ASIC electrodes into focus. The arm is lowered gradually and 160°C is applied for 30 seconds with 70N of force. Once the ACF is cured, 100°C is applied for 2 minutes to remove the copper and thermal release tape. The top-contact of the composite is then defined by applying conductive silver paint (Ted Pella, USA) onto the edges of the composite to connect the top-contact to the appropriate pads on the ASIC. This paint is cured at 60°C for 1 hour.

### Support electronics

The flex-board of the FlexUS device is connected to two small rigid support boards, a configuration board which programs the FlexUS device and provides power, and the output board, which performs data conversion and data transfer to a host computer. There is a wireless version of these support boards, as well, (see **Supplementary Discussion S8**) which were not used in these applications.

The configuration board of the FlexUS device generates all power supplies and scan-chain controls for the ASIC. Buck converters are used for the low voltage supplies to improve efficiency. A 5-V-to-3.8-V Buck converter is used in conjunction with an adjustable low drop-out linear voltage regulators (LDOs) to generate the 1.8-V and 3.3-V power supplies. Separate 1.8-V analog and digital supply voltages are created to provide power-supply-noise isolation. The HV supply is generated using a 5-V-to-32-V Boost converter, with the outputs then passed to HV LDOs to generate the 29-V supply. Positive-temperature-coefficient resettable fuses are added in series with all power supply outputs to protect from overdraw in case of device damage or incorrect connection. The flex-board is connected to the configuration board through two 80-pin connectors and is fastened to the connector using a cable sleeve and plug-jacket, which ensures connectivity regardless of the tension on the cable region of the patch. An FPGA module is used to program the scan-chains and check their outputs to confirm accurate ASIC programming (see **Supplementary Discussion S4**).

The output-board contains off-the-shelf analog-to-digital converters (ADCs), that are four-channel modules with 10-bit ADCs of sampling rates up to 65 MSamples/s (AD9219, Analog Devices). The ASIC outputs are first buffered by fully-differential unity-gain feedback opamps (THS4524, Texas Instruments) in four-channel packages using 1 kΩ feedback resistors that can be compactly placed next to the ADC. The ADC outputs are low-voltage differential signaling (LVDS), which are driven onto 100-Ω-differential impedance-controlled traces with 100-Ω-differential loads at the receiving FPGA pins to reduce reflections. Differential coplanar traces are used since they introduce ground shielding nets on either side of the differential pair, minimizing crosstalk between output channels. The FPGA on the output board (XEM7360, OpalKelly) interfaces to dynamic random-access (DRAM) memory (IS43TR85120BL, Integrated Silicon Solution, Inc.) to synchronize all outputs of the 64 ADCs, to store the data from all beams in a single frame, and then to transfer this frame data to the host computer at the end of each frame (see **Supplementary Discussion S4**). The output-board uses the same 80-pin connectors as the configuration board for connection to the flex-board.

### ASIC electrical characterization

For electrically testing the ASIC prior to packaging with the piezoelectric composite (data presented in **Supplementary Discussion S3**), a modified output-board with outputs connected to header sockets is used with the outputs directly connected to a Keysight InfiniiVision oscilloscope (DSOX4034A, Keysight, USA) controlled with PyVISA, an open-source tool based on the NI-VISA (National Instruments, USA) backend. For the gain tests, a single-pixel is stimulated in a probe-station using a DC probe (DCP- 100, FormFactor, USA) over the frequency range from 100 kHz to 30 MHz. The programmable gains are varied from 0 dB to 36 dB with the input amplitude set such that the output voltage does not exceed 640 mV peak-to-peak.

We next test the beamforming phase shifts on the chip. To test the fine μBF phase control within a sub-array, we enable two pixels in that sub-array with fixed gain while disabling the other two pixels. The corresponding coarse µBF unit is bypassed to record the output of the selected sub-array alone. One pixel has its delay setting kept constant while the other has its delay swept across the full range of fine µBF delays over the same range of frequencies used for the gain measurements. To measure course μBF phase control, two pixels that correspond to the same coarse µBF (in the same 4×4 cluster) are selected with one held constant and the other having its delay varied. This test is repeated for pairs of pixels that do not correspond to the same output channel. In particular, pixels in one 4×4 cluster are compared to pixels in different corners of the array to determine the worst-case phase error between different pairs of pixels.

To test the transmitters, the rise time, on-voltage, transmitter beamforming delay, and pulse-width are measured for the transmitted waveforms differentially between two pixels, the delay between which is varied. A HV active differential probe with a 1.5 pF input capacitance (DP0010A, Keysight, USA), which closely matches the load presented by the transducer is used for these measurements.

To measure the output-referred noise of the ASIC for a single output channel, the selected output channel goes to the output-board which then drives the 50 Ω load of the spectrum analyzer (E4448A, Keysight, USA) through a DC block. A resolution bandwidth of 10 kHz was found to be a sufficient compromise between measurement time and resolution. To subtract the noise power of the output board from the measurement, the noise added by the output board is measured by shorting its input and measuring the “baseline” power spectral density (PSD) at the output. Shorting is the appropriate approach here because the ASIC output is low-impedance (10Ω), making voltage-mode noise dominant. Then, the ASIC is connected with all transducer pads shorted together and connected to the mid-rail common voltage. For the maximum LNA and PGA gain settings, the output noise PSD is measured with the baseline PSD subtracted from this.

### SEM cross-sectional imaging

Samples were cross-sectioned either through manual cleaving with a sharp steel blade or by using a precision saw (IsoMet, Buehler, USA) at low speed with a SiC blade. To improve the smoothness of the interface, a grinder-polisher (SBT910, South Bay Technology, USA) with progressively finer SiC grit pads is used followed by an alumina slurry polish. Secondary electron SEM images are acquired using a scanning electron microscope (Sigma VP, ZEISS, Germany) operating at 5 kV. A 5-nm-thick layer of AuPd is sputtered onto the samples to prevent charging. False coloring is done using the fuzzy select and colorize tools in GIMP v2.10.

### Plane wave test

The resistance of the connection through the ACF of the piezocomposite to the ASIC is quantified by performing a plane wave test with a short column of water (1 cm) above the ASIC. A 3D-printed, clear-resin tank of inner diameter 48 mm, thickness 4 mm and height 40 mm, is attached to the FlexUS device by using polydimethylsiloxane (PDMS) and cured at 60°C over 12 hours in an oven. This tank is then filled with a constant water column (25 ml) of distilled and de-gassed water. The ASIC is then tested row-by-row and column-by-column with only one pixel per coarse µBF stimulated at a time (see **Supplementary Figure S4**) and the reflected wave measured at the same pixel. The received signals are band-pass filtered between 1 MHz and 8 MHz, and the measured peak-to-peak amplitude is used to characterize transducer conductivity to the ASIC.

### Transmitter acoustic characterization

To characterize the transmitters, hydrophone measurements of the pressure profile are performed. As done for the plane wave test, a 3D printed water tank is attached to the flex-board using PDMS. A bullet hydrophone (HGL-0200, Onda Corp., USA) is connected to a preamplifier (AG-2010, Onda Corp., USA) with measurements taken on an oscilloscope (DSOX4034A, Keysight, USA). The hydrophone and preamplifier are mounted on a custom 3D-printed attachment to a 3D motorized stage (MTS25-Z8, ThorLabs, USA) driven by stepper motors (KST101, ThorLabs, USA). The hydrophone has a 200-µm-diameter aperture and is rated for frequencies between 0.25 MHz and 40 MHz with a calibrated sensitivity of 43 nV/Pa. De-gassed distilled water is used in these measurements. The oscilloscope triggers on a transmit enable signal routed from the configuration board to the scope. The hydrophone is scanned in a grid pattern with 0.5 seconds of settle time at each location. The peak-to-peak voltage is recorded and then divided by the 43 nV/Pa sensitivity of the hydrophone and the 10× gain of the pre-amplifier to determine the pressure at the measured point. To determine the efficacy of the matching layer, the measurements are conducted before (see **Supplementary Figure S11**) and after (**Fig. 3**) SU8 deposition.

### Bandwidth characterization

The transmit bandwidth of the transducer was calculated from the fast Fourier transform (FFT) of the transmitted pulse. The same 3D printed water tank that was used for acoustic characterization was affixed to the device and the hydrophone was moved to a position where it would capture the peak pressure for a plane wave transmission. The device was configured to apply a single cycle of a 6 MHz pulse (50% duty cycle), which is the narrowest pulse to which the transmitter circuit can be configured with reliable operation due to limitations in delivery the higher-frequency clocks needed to support shorter pulse lengths.

The transmitted waveform was captured using an Infiniivision oscilloscope (Keysight, USA) set to sample at 100 MS/s. This waveform was transformed to the frequency domain (MATLAB fft) and the peak frequency (natural frequency) and 6 dB bandwidth of the transducer were measured. Since the bandwidth is a property of the transducer’s impulse response, the rectangular shape of the pulse needs to be accounted for, with its own FFT removed from the transducer’s response to the 6 MHz pulse. The applied pulse (∼83 ns wide) is a zero-order hold (ZOH) filter’s output of a true impulse and would produce a sinc response (with first null at 12 MHz) in the frequency domain. The measured bandwidth of the transmitted pulse and the impulse response of the transducer obtained by correcting for this sinc response are plot (see **Supplementary Figure S5a)**. The 6 dB one-way bandwidth estimated in this manner is 1.93 MHz, corresponding to a FBW of 43.9% after correction and 45.7% before correction, using the measured center frequency of 4.4 MHz.

For the two-way bandwidth, i.e., the impulse response of the system for both transmit and receive, a reflector is needed. The wire phantom is used since the wire targets are quite small and have sufficient impedance mismatch to reflect the transmitted signals. The two-way bandwidth measured in this way includes the transducer bandwidth twice (both transmit and receive) along with the receiver front-end electronics bandwidth. The drawback of the chosen approach is that it assumes perfect reflection at the wire target and no frequency dependence on the reflection. The device again transmits a single pulse at 5 MHz, which propagates through the water column, get reflected at the wire target and propagates back to the transducers where it is received. The received FFT is then plot as a function of frequency before and after the ZOH correction (see **Supplementary Figure S5b**). The 6-dB two-way bandwidth was found to be 1.47 MHz or 33.9% after correction, with the decrease in bandwidth expected due to the transducer response being present twice in the two-way system response.

### MDP and NEP estimation

The first step in determining the minimum-detectable pressure and noise-equivalent pressure is obtaining the receive transfer function of the array. While a commercial probe could be used as the source of the transmitted energy, it would need to be characterized first to determine the incident pressure. Instead, a water column above the FlexUS device of 15 mm height was constructed above the array to characterize the transfer function based on received reflections of its transmit. This height was chosen as half the distance at which maximum plane wave pressure was achieved (**Fig. 3c**), and should be within the nearfield based on the Rayleigh distance for our device. This distance was also validated by the fact that the transmitted pressure for a plane wave stimulus did not decay significantly within our measured pressure field (**Fig. 3c**). The device transmits a 3-cycle pulse**-**width 5 MHz plane wave with the array positioned so that ultrasound propagation was normal to the face of the water column, and the received pressure from the reflected wave is estimated to be 3 MPa (peak-to-peak) for the round-trip distance of 30 mm (from **Fig. 3c**). The peak-to-peak amplitude of the received pulse for the center channels is 757 bits, which translates to 0.734 V. For the 3 MPa incident pressure, assuming no loss of pressure due to attenuation in water and to beam divergence, this translates to a receiver efficiency of 245 nV/Pa. Using the noise floor estimate of 11 nV/√Hz for a maximum receiver gain of 36 dB, which integrates to 15.3 µV_RMS_ across the device’s measured 43.9% fractional bandwidth, this gives an MDP of 62 Pa_rms_ and an NEP of 45 mPa_rms_/√Hz. This measurement is for a single channel alone, and the metrics are expected to be lower with receive beamforming.

### M-mode lung images

A 1.2-mm-wide strip in the center of a real-time YZ-centric B-mode image is chosen as the ROI. The Y points in the ROI are averaged to obtain a single Z-domain brightness line for each time stamp in the multi-frame acquisition and are plotted against acquisition time.

### Brightness tracking for breathing rate

A 1 cm deep and 2.6 cm wide ROI was defined around the pleural region and the image within this region was Gaussian-blurred (MATLAB imgaussfilt) with a standard deviation of roughly 0.4 mm applied to remove effects of randomly decorrelated speckle caused by the relatively low frame rate of the ultrasound acquisition. The standard deviation was chosen to trade-off noise reduction with spatial resolution since a wider Gaussian kernel blurs anatomical details while suppressing noise. To increase the contribution of points within the ROI that have a time-varying component and to prevent points with high brightness across frames from saturating the brightness signal, a filter based on the coefficient of variation (normalized standard deviation) of each point in the ROI across frames was generated. A sigmoid function was optionally applied to this filter but the effect on the brightness metric was minimal. Finally, the brightness value of all points within the ROI were summed for each frame and the brightness-sum was plotted against time to observe variation with chest height.

### Chest height tracking

The Shi-Tomasi algorithm^38^ (“goodFeaturesToTrack()” in OpenCV Python library) was used to detect boundary points on the subject’s torso and the Lukas-Kanade algorithm^39^ (“calcOpticalFlowPyrLK()” in OpenCV) was used to track the movement of these points across frames.

### Human experiments

All experiment were conducted under the Columbia University Institutional Review Board Protocol AAS6542, entitled “Ultrasound imaging using a flexible patch transducer array.” For the neck imaging, the subject is placed on a table with support under the shoulder blades to raise the chest and allow extension of the neck, with their head tilted to the right to provide sufficient space for FlexUS device placement. For the lung imaging, the subject is once again placed on a table with the arms to the side. All human experiments were conducted with 2-cycle pulse-width plane waves at 5 MHz excitation frequency and a maximum pressure of 3 MPa peak-to-peak, which corresponds to a mechanical index of 0.67.

### 3D Reconstruction Methodology

3D reconstructions are created by amplitude-thresholding pertinent reconstruction-grid points in the 3D acquisition and plotting them together on a 3D grid (“scatter3”, MATLAB). The thresholds are determined based on the regions of interest (IJV/lung/phantom) that need to be visualized in the 3D rendering. Threshold values that are too low will lead to low amplitude speckle points overwhelming the 3D rendering and obscuring features of interest, while threshold values that are too high prevent the visualization of features that are made of volumetric regions of medium-amplitude points (such as the volume targets in the phantom). For the 3D reconstructions of the wire phantom (**Fig. 4d**) the upper threshold was set to 20 dB and the lower threshold was set to 15 dB. For the CIRS phantom (**Fig. 4h** and **Fig. 4k)** the upper and lower thresholds were set at 5 dB above and below the gains of the corresponding targets respectively (9 dB for the volume object and 15 dB for the rod target). For the 3D reconstructions of the IJV and CA (**Fig. 5d**), the thresholds were set much lower to 3.5 dB and 2.5 dB respectively in order to plot the regions of low speckle/brightness which correspond to the blood vessels.

## Data Availability

Most data produced in this study are available online at https://github.com/klshepard/flexus. Additional data are available upon reasonable request to the authors.

## ACKNOWLEDGEMENT

This work was supported in part by the Defense Advanced Research Project Agency under Cooperative Agreement D20AC00004 (KLS) and the National Institutes of Health under Grant R18EB037607 (EEK).

## AUTHOR CONTRIBUTIONS

JSL, SG, and KLS conceived the project. JSL and SG designed the ASIC, support electronics, and software. HY, JDF, SG and JSL performed the packaging and assembly of the FlexUS device. DOK assisted with the human experiments. SGB helped with image analysis. JSL, SG, and KLS wrote the paper with contributions from all the authors. DOK, EEK, and KLS supervised the work. KLS and EEK provided funding support.

## DATA AVAILABILITY

All imaging data relevant to the figures presented in this paper are available at https://github.com/klshepard/flexus. All other relevant data are available from the corresponding authors upon reasonable request.

## CODE AVAILABILITY

All scripts used for data analysis are available at https://github.com/klshepard/flexus. All other relevant codes are available from the corresponding authors upon reasonable request.

## SUPPORTING INFORMATION

Supporting information includes Discussions S1 through S8, Figures S1 through S31, Tables S1 through S3, Supplementary Movies S1 and S2.

