## Supplementary information for "A Conformable CMOS Ultrasound System for Point-of-Care Imaging"

### Supplementary Discussion S1. Effects of delay quantization on beam-forming

We performed Field-II[1] simulations to determine resolution limits and understand the effects of delay quantization and averaging techniques on beam-forming accuracy.

**Delay resolution of receiver beam forming.** The 32×32 array of transducers, that are 150-µm wide and spaced 55-µm apart, is simulated for a 5-MHz center frequency and a 3D FOV with a point source in its focal region. This point source is kept at 15-mm focal depth and moved along different azimuth (−30° to +30°) and elevation (−15° to +15°) angles with the lateral resolution estimated from the full-width-at-half-maximum (FWHM) of the corresponding spot formed in the image. We expect that the maximum resolution will be achieved at 15-mm depth, since resolution generally degrades with depth and the device is not expected to be able to focus at shallower depths. To determine axial and lateral resolution, the sectors are plotted along the azimuth (**Fig. S12a**), and to determine elevational resolution, the sectors are plotted across elevation parallel to the YZ plane (**Fig. S12b**). At the 15-mm focal depth, axial, lateral, and elevational resolutions are 450 µm, 660 µm and 737 µm respectively. Axial resolutions can be expected to be worse that these estimates because Field-II does not consider the effects of the acoustic stack-up on bandwidth. These simulations also assume infinite delay resolution, when these delays are, in fact, quantized.

**Delay quantization.** For *N* bits of delay quantization, the system clock is required to be $2^N$ times higher than the 5-MHz center frequency of the array, with the delay quantization (for both transmit and receive) determined by the period of this system clock. While some designs have achieved beamforming through purely analog techniques yielding very high delay resolutions[2], this is not scalable to 1000s of electrodes with in-pixel transceivers since it requires the routing of six clocks of different phases from a delay-locked loop (DLL). In addition to the circuit overhead to create these phase interpolators, there is also not much re-usability between the transmit and receive circuits which would lead to a larger pixel area. The low center frequency of the design when compared to radar arrays allows the use of true-time-delays as phase shifters rather than vector modulators, eliminating the possibility of beam-squint from variation of phase shifts with frequency, greatly simplifying the transmitter design.

**Effects of delay quantization in receiver beam forming on resolution.** To determine the effect of quantization on receiver beamforming, we first vary the number of bits used for the delay (*N*) and simulate the change in resolution for point sources (**Table S2**). Beyond $N=3$, there is little resolution improvement as the large number of elements in the array helps to compensate for any effects of delay quantization such as phasing inaccuracies. At $N=2$, the energy of the point scatterer spreads more, especially when steered, leading to poorer image quality and degraded resolution. $N=4$ is used in the FlexUS device. The axial resolution is largely unaffected by this delay quantization since it is only a function of the receiver bandwidth and acoustic material stack.

**Effects of delay quantization and channel averaging on receiver beam forming.** For $N=4$, all delays in the array are quantized to the period of an 80-MHz system clock, i.e., 12.5 ns, which can cause errors in the directivity of the beam. Some designs perform channel averaging in lieu of beam forming as a means of reducing on-chip circuit complexity[3], a design simplification not performed on FlexUS. This averaging also results in errors in beam directivity. **Fig. S13** considers these effects in a MATLAB simulation of the array factor for a 32-element linear array using different beam-forming techniques for three distinct angles ($0^{\circ}$, $+15^{\circ}$ and $-30^{\circ}$): ideal (non-quantized), quantized to multiples of 12.5 ns, and every-two-element averaging. Array factors in each case are normalized to the ideal beamformer array factor at the same angle. The quantized beamformer can steer exactly to $0^{o}$ and $-30^{o}$ but can only steer to $14.4^{o}$ when programmed to $15^{o}$. The main-lobe energy and side-lobes remain unchanged from the ideal plot. For the case of two-element averaging, the grating lobe worsens with increased steering angle as the main lobe energy also reduces. This is because the effective element size in this case has twice the pitch[4].

**Beamforming on ASIC.** The quantized beamforming delays must be programmed into three sets of circuits on the ASIC: the transmit (TX) pulse generators, the fine µBFs, and the coarse µBFs. Across the full 32×32 array the worst-case delay reaches 3.4 µs, which would require eight bits of storage at each of the 1024 pixels, prohibitively area-intensive given the in-pixel transceiver. Decomposing each delay into a sub-array component plus a pixel residual removes this cost. The largest delay span within any single 2×2 sub-array is bounded (<187.5 ns by Field-II[1] simulation, for all transmit beam patterns and target focal depths), such that only four bits per pixel are needed to cover the residual, while the eight bits that span the full dynamic range are stored once per sub-array and shared by its four pixels.

**Fig. S14** illustrates the conversion. For each 2×2 cluster of pixels the minimum of the four ideal delays is taken as the sub-array delay, and the per-pixel delays are obtained by subtracting this sub-array delay from each pixel's ideal delay. The same four-bit pixel delay is used on both transmit and receive, halving the per-pixel flip-flop count; this is acceptable because most received energy arrives from the transmit direction, and it avoids the alternative of simply averaging pixel outputs. The coarse µBFs, which operate on receive only, follow the same min-subtraction rule when receive beamforming is aligned with the transmit direction (A-mode); for B-mode imaging the coarse µBF delays are instead chosen to maximize reconstructed SNR and contrast.

**Supplementary Discussion S2. FlexUS ASIC circuit design**

We consider the circuit implementation of the key components of the FlexUS ASIC design.

**Low-noise amplifier (LNA).** A fully-differential folded-cascode architecture (**Fig. S15**) is used for the LNA for power-supply-rejection-ratio (PSRR) in excess of 50dB. A resistor load is used instead of an active load to prevent the flicker noise that an active load would introduce[5]. The small area available for the LNA limits the maximum achievable size of the input device and sets the flicker noise corner in the 100s of kHz, i.e., just below the operating bandwidth of amplifier, between 2 MHz and 10 MHz. This LNA topology also uses cascoded devices to increase gain; these devices, however, only add noise at frequencies greater than their unity-gain current cutoff frequency ($f_T$), which is in the 10s of GHz. The output devices are sized to consume half the current of the differential pair to further reduce the noise contribution of the tail devices *M6a* and *M6b*, which are sized to have transconductances of less than 100μS.

A common-mode feedback (CMFB) circuit is required to bias the tail device *M6*. A typical CMFB circuit is a two-pole system. The first pole is at the output of the LNA, while the second is at the output of the CMFB amplifier, which in our case would be the gate of *M6*. To keep the CMFB loop stable with 60° phase margin, the loop-gain bandwidth must be twice the non-dominant pole, which is the output pole of the CMFB amplifier. Given the high gain of the *M5*- *M6* cascode, this would be quite difficult to achieve and either the loop gain would have to be reduced or the pole would have to be pushed to a higher frequency. The latter would involve changes to the LNA itself, such as downsizing *M6* (which is already quite small with a $g_m/I_D$ of 4). To reduce loop gain, the $g_m$ of the CMFB amplifier would have to reduced, making the $V_{GS}$ of the CMFB input device larger and limiting the range of LNA output voltages that can be supported.

For these reasons, CMFB for the FlexUS LNA is instead provided by a passive switched-capacitor (SC) circuit[6], which is an inherently stable single-pole system. For faster settling, $C_{FB1}$ is set larger than $C_{FB2}$; however, setting $C_{FB1}$ too large reduces the gain of the LNA if the $1/\ fC_{FB1}$ equivalent resistance of the CMFB circuit, where $f$ is the switching frequency of the SC circuit, becomes smaller than the output impedance of the LNA. We set $f$ = 20 MHz, meeting Nyquist rate requirements for the targeted maximum frequency of operation of 10MHz. This SC CMFB circuit settles within four cycles (200 ns) and is enabled before the receive phase begins. MOS capacitors are used, providing approximately 10 fF/μm$^2$ of capacitance. The CMFB circuit is disabled during the transmit phase causing the output devices of the LNA to float, thereby turning off the LNA and providing a power-gate mechanism.

The LNA has programmable gain in steps of 6 dB from 0 dB to 24 dB. This gain is realized through capacitive feedback ratios with the feedback capacitor held constant and the input capacitor varied. The unit capacitor is sized such that the input impedance of the LNA at maximum gain is still higher than the resonant impedance of the piezoelectric element to avoid signal divide-down at the LNA input. DC feedback is achieved through a large feedback resistor (250 kΩ). While area-inefficient compared to other active feedback techniques, the resistor sets a high-pass pole at 2 MHz, reducing low-frequency input-referred noise. The low-pass corner is set by the unity-gain bandwidth, which is designed to be 200 MHz, and varies with gain setting. At 24 dB gain, this corresponds to a 10 MHz pole. The closed-loop LNA has a phase margin of 50° and input-referred noise of 17.5μ$V_{RMS}$, while consuming 360μW of power. Assuming an ASIC output swing of 1V, a maximum system gain of 32, and 10-bit analog-to-digital converters (ADCs) at the output, this noise level correspond to less than two least significant bits (LSBs).

**HV transmitter.** The HV transmitter (**Fig. S16**) takes the 80-MHz system clock and scan-chain delay configuration as input and generates the required 5-MHz output pulses for driving the power amplifier (PA). The system clock is divided down in two steps, from 80 MHz to 20 MHz (to provide the clock required for the SC CMFB) and then from 20 MHz to 5 MHz with the output beginning with a positive edge to ensure a known initial state for the PA. The programmable pulse width of the 5-MHz signal, which can range from one to seven cycles, is determined by a three-bit counter. The pulse signal is level-shifted to 5-V using a latch-based level shifter and inverted (by *INV_N*) to drive the gate-to-source of the 29-V input device (*MN1*) of the PA through a transmission gate and a grounding device that turns off *MN1* when it is not transmitting. By starting driving pulses on the positive edge, the piezoelectric element is always grounded before being disconnected from the HV transmitter to prevent any charge from being left on it. *MP1* of the PA requires input voltages between 24V and 29V to provide a 5-V gate-to-source voltage. This DC shift is accomplished by using an AC coupling capacitor. To reduce the settling time for turning on the transmitter, a half-latch device is added between the output of inverter *INV_P* and its input.

The T/R switch also uses the 29V HV devices and requires its own level shifter. It uses a three-switch nFET-only architecture[3], that provides two redundant open switches, isolating the RX inputs and holding the middle node to a low voltage to mitigate the effects of capacitive coupling. The use of an nFET-only structure simplifies control since only 5-V level shifters are required without the need for DC level shifting. The T/R switch is shorted after a globally programmable time delay from the end of the transmit event to avoid saturation of the RX amplifiers from initial reflections off the transducer-surface-to-skin interface.

**Micro-beamformer (μBF)**. A fully-differential passive topology is used for the μBFs (**Fig. S17a**). The implementation includes 16 450-fF unit-capacitors, implemented as metal-insulator-metal (MIM) capacitors, allowing for the full-range of delays (up to 187.5ns) required to beam form in-pixel (see **Supplementary Discussion 1**). The LNA output is sampled onto these 16 unit-capacitors by closing the switches $S_1$ to $S_{16}$ one-at-a-time. The μBF outputs are read by closing switches $R_1$ to $R_{16}$ in the same order but after a programmed delay (**Fig. S17b**). The outputs of four pixels are then combined together at the parasitic input capacitance of the PGA. This parasitic capacitor produces a low-pass infinite impulse response (IIR) filter since each output depends on both the current charge integrated onto the capacitor as well as the past; this results in an additional effective pole at 10 MHz which further helps to set this as a sharp bandwidth limit. Monte Carlo simulations show means in the delay within 0.3 ns of the ideal delay with standard deviations under 3ps. The second and third harmonics are found to be at 65 dBc and 70 dBc, respectively.

**Programmable Gain Amplifier (PGA)**. The PGA also uses a fully-differential telescopic cascode architecture (**Fig. S18**). The CMFB structure used is identical to that in the LNA, and uses the same 20-MHz clock. The PGA is designed to have the same 2 MHz to 10 MHz band-pass response to minimize in-band noise power and consumes 144μW of power.

**Supplementary Discussion S3. ASIC electrical characterization**

Prior to integration with the piezoelectric transducers, extensive electrical characterization of the ASIC was performed.

**Gain measurements**. A wide range of pixels across multiple ASICs was tested, including major and minor diagonals, edge columns and rows, and central columns and rows arranged in a criss-cross pattern (**Fig. S19a**). Pixels were selected to capture the effects of bias distribution, power-supply distribution, and PCB output-trace length. For example, pixels in the edge columns experience the lowest bias currents, pixels near the center of the ASIC experience greater power-supply droop, and pixels on the left side of the ASIC have the longest output-trace routing on the PCB.

The outputs from all sweeps were collected and averaged to obtain the receiver front-end transfer function (**Fig. S19b**). The worst-case gain variation across all pixel combinations and gain settings for a single ASIC was found to be ±0.6 dB. This analysis accounts for differences in routing lengths between ASIC outputs on the flex-board and the output board, which can be as large as 13 cm between the shortest and longest signal paths, further demonstrating the robustness of the ASIC to varying loads on the pad drivers.

Most importantly, the small variation in frequency response with gain ensures that there will not be any significant phase change between channels during beamforming due to gain inaccuracies. The system bandwidth was measured to be 2–8 MHz at higher gain settings (12–36 dB) and 1–10 MHz at the 0 dB and 6 dB gain settings, meeting the target fractional bandwidth of 120%.

**Phase measurements**. The fine μBF test was performed over the same frequency range as the gain measurements and used a similar methodology, measuring the FFT magnitude at the stimulation frequency. The primary differences were that the second channel of the function generator was enabled and configured to track the first channel, and that both pixels were enabled without bypassing. For this sweep, an LNA gain of 12 dB and a PGA gain of 0 dB were used, with an input amplitude of 160 $mV_{pp}$. Delay settings were randomized to avoid the introduction of experimental bias or systematic error.

A wide range of sub-arrays across two ASICs was tested. Because all fine μBFs reside within sub-arrays with identical physical layouts and are digitally controlled, no significant performance variation was expected apart from differences in digital signal routing. Accordingly, sub-arrays were selected from both the center and the edges of the array (**Fig. S20a**). Within each sub-array, one pixel was randomly chosen as the reference (constant) pixel, and the second probe was sequentially placed on each of the remaining three pixels, fully characterizing the selected sub-arrays.

As expected, both constructive and destructive interference were observed (**Fig. S20c**). For clarity, only integer frequencies from 3 MHz to 7 MHz are plotted. The reference pixel delay was fixed at a setting of 4, and all frequencies exhibited peaks at the same delay setting. With the system clock operating at 80 MHz, only the 5 MHz and 4 MHz tones exhibit perfect nulls at delay differences of 100 ns and 125 ns, respectively. The destructive interference points for 6 MHz and 7 MHz cannot be precisely quantized by the 80 MHz clock and therefore show only partial nulls, while a full 180° phase shift cannot be achieved within 15 delay steps at 3 MHz.

The coarse μBF test was conducted using the exact same experimental methodology. Now, however, the corresponding coarse μBF units are set to delay-and-sum rather than bypass to combine the two inputs. The fine μBFs are set to bypass since only one pixel per sub-array is used. The delays are randomly programmed, but a wider range of coarse μBFs are tested across multiple ASICs (**Fig. S20b**), since more variability is expected due to the differences in PGA output routing between sub-arrays. Despite the differences in sub-array output routing lengths, the same points of constructive and destructive interference are seen as with the fine μBF test but over twice the range due to the larger delay range of the coarse μBF. A notch can now be seen for 6 MHz since a 540° phase shift can be sampled at 80 MHz.

The final tests were on global phase, i.e., pixels that do not correspond to the same output channel. These measurements were performed across two ASICs, with pixels selected to capture variations in on-chip and PCB routing lengths, bias conditions, and supply voltages (**Fig. S21a**). The same pixel was held constant in all tests, and the resulting amplitude-versus-delay waveforms at 5 MHz were plot to illustrate the best- and worst-case delay mismatches between tested pixel pairs (**Fig. S21b**). The worst-case delay difference between the best and worst pixel pairs on any given ASIC was found to be 7.95 ns, i.e., less than one unit delay step (12.5 ns). This indicates that no additional delay padding is required to align the received waveforms, significantly simplifying data acquisition and validating the low-mismatch design techniques employed, albeit at the cost of increased power and area.

The global measurements above, however, did not account for the impact of varying PGA gain. Higher PGA gain settings are expected to introduce additional phase variation due to reduced bandwidth and increased sensitivity to differences in sub-array output routing length. To evaluate this effect, the experiment was repeated with both pixels configured to the same PGA gain, which was varied between 1×, 2×, and 4×. Identical gains were used for both pixels, reflecting standard ultrasound operation in which gain is adjusted uniformly across the array over time rather than independently between pixels. Under these conditions, the delay difference between the best- and worst-case pixel pairs increased to 13.55 ns (**Fig. S21c**), marginally exceeding the unit delay step of 12.5 ns. This discrepancy is sufficiently small and can be neglected, as the output data are sampled at 20 MHz and the resulting phase error is not resolvable. Moreover, the large number of array elements further averages out the impact of this residual phase mismatch.

**Transmitter characterization.** The same pixels used for coarse μBF verification were evaluated, and the measured waveform differences were plotted as a function of delay setting for all tests (**Fig. S22a**). No deviations from ideal behavior were observed, with the measured offset equal to the pulse width (6.5 cycles at 5 MHz, corresponding to 104 delay settings). During these tests, all transmitters were enabled—not only those under test—to evaluate worst-case IR droop across the array. Under these conditions, the maximum observed droop was 28.4 V.

The transmit waveforms exhibited rise times within 12.5 ns, corresponding to one-eighth of the positive pulse duration at 5 MHz (**Fig. S22b**). This performance was sufficient to drive the transducers and supports programmable delays ranging from a minimum of 12.5 ns to a maximum of 3.4 μs, thereby achieving the designed delay resolution of 12.5 ns.

**Noise characterization.** The measured output-referred noise power density, in units of Watts/Hz, is multiplied by the measurement impedance of 50Ω to get $V^2$/Hz, and then the square-root is taken to get V/√Hz. The input-referred noise is then calculated by dividing by the system gain. The flicker noise corner was approximately 100 kHz, so it would not be a major contributor to the in-band noise, and the input-referred noise at the thermal-noise limit at 5MHz was found to be 11 nV/√Hz. At the output amplitude corresponding to the 1dB compression point (-3.6dBm), the signal-to-noise ratio (SNR) of the receiver for the full 120% bandwidth (2 MHz-8 MHz) was determined to be 57dB. The normalized output power spectral density (PSD) at this 1dB compression point is shown in **Fig. S23**. Based on the 1dB compression point and the noise floor, the dynamic range (DR) of the system was estimated at 88dB.

### Supplementary Discussion S4. Firmware and software development

Firmware was developed for the FPGAs of the configuration board and the output board. Software on the host computer transfers data and performs image reconstruction.

**Configuration board firmware.** The FPGA Verilog implementation (**Fig. S24b**) on the configuration board (**Fig. S24a**) uses on-module dynamic random-access memory (DRAM) to allow the scan-chains for all 128 beams in a slice to be programmed simultaneously. A finite state machine (FSM) forms the core of the implementation and programs the ASIC using four states: **Idle**, **Shift**, **TX**, and **RX**. The FSM remains in the RX state until a reset, such that each FPGA trigger initiates exactly one transmit–receive cycle. The FSM states operate as follows:

- **Idle**: Waits for a trigger to begin the scan-chain operation.
- **Shift**: Shifts in the scan-chain data, then transitions to TX.
- **TX**: Asserts the transmit enable one clock cycle after the transition from Shift. The FSM remains in TX for a preprogrammed duration to allow all pixels to transmit, then transitions to RX.
- **RX**: Waits for a preprogrammed delay after TX to prevent amplifier saturation from early reflections, enables the receivers, and remains in this state until the FPGA is reset.

To efficiently transfer data, the design leverages OpalKelly's block-throttled pipe (BTPipe) construct, which is optimized for block-based communication and eliminates frequent, time-consuming transactions between the configuration board and the host PC that would otherwise limit the achievable frame rate. With the BTPipe, the FPGA controls when data are read via a READY signal; once asserted and invoked from Python, data are transferred until a specified block size is reached.

The BTPipe is used for scan-chain input, with a dedicated controller responsible for asserting the READY signal. The controller output feeds an input first-in, first-out buffer (FIFO) that stores the pixel, sub-array, and coarse μBF scan-chains for a single beam. These three scan-chains, together forming a $576 \times 32$ binary vector, are combined into a single DRAM packet. Once a packet is transferred into the input FIFO, the BTPipe controller asserts the input FIFO read enable and the Sync FIFO write enable. The Sync FIFO stores the data in a 256-bit-wide format suitable for DRAM access.

A DRAM arbiter interfaces with the DRAM and writes data as soon as entries are available in the Sync FIFO, continuing until a complete packet is stored. Once the input FIFO is emptied, the BTPipe controller reasserts the READY signal to load the next beam's scan-chain. This process repeats until the DRAM contains 128 packets, corresponding to all beams in a single slice. At that point, a single packet is read into the Data FIFO.

The Data FIFO controller waits for the initial trigger from Python, then separates the data into pixel and sub-array/coarse μBF scan-chains. These are shifted row-by-row into their respective FSMs, which operate at 80 MHz but with different clock phases (0° for pixel scan-chains and −45° for sub-array/coarse μBF scan-chains). This phase offset reduces simultaneous switching activity at the FPGA outputs and helps relax power-supply requirements. As the data are shifted into the FSMs, they are also copied into write-back FIFOs. After a full packet is transmitted to the ASICs, the data are retransmitted from the write-back FIFOs to read out the ASIC scan-chain outputs from the initial scan. The ASIC outputs are then compared with the inputs, and the comparison results are stored in a FIFO. This approach allows all 128 beams to be processed sequentially, with comparison results accumulated and transferred together, significantly reducing overhead.

Following the initial Python trigger, the configuration board is subsequently triggered internally. During the RX state, the main FSM includes a counter that, after a preprogrammed interval corresponding to the maximum target depth of 56 mm, triggers the Data FIFO controller to initiate the next beam. This process continues until the final beam is completed, at which point the FSM remains in the RX state until a system reset.

**Output board**. Data arrive at the output-board FPGA as 1 bit per cycle from each of the 64 output channels. While this data could be written directly to DRAM and transferred to the host computer, the conversion overhead is substantial because each row of the output vector contains only one bit per channel. This conversion can take up to 5s per slice since it involves operating on binary arrays in Python. To reduce this overhead, the incoming data are first passed through a de-serializer (**Fig. S25**).

Each de-serializer uses a 16-bit-long, 8-bit-wide two-dimensional Verilog vector to shift incoming bits sequentially along the vector width. The width is chosen to be 16 bits to enable efficient decimal conversion using NumPy's **frombuffer** function, which operates on byte-aligned data. The output board is divided into eight regions, each clocked by a different phase of the 200 MHz data clock to synchronize all channels. Consequently, each de-serializer operates on eight channels, requiring a total of eight data de-serializers (DESERs). The upper six bits of each row in the 2D vector are initialized to zero. After 10 clock cycles—once all bits corresponding to a single sample across the eight inputs have been shifted in—the output is generated in a single cycle using a Verilog **for** loop that traverses the vector row-by-row from MSB to LSB. This introduces a single 20-MHz sample-clock cycle of latency, which is compensated by delaying the FIFO write enables by 10 cycles of the 200 MHz data clock. Through this process, the data are converted from a 10-bit serial representation per channel into a 16-bit parallel representation.

The de-serialized data are then written into eight Data FIFOs, with eight corresponding Data FSMs tracking the number of samples prior to de-serialization. Once the programmed sample count threshold ($N_{samp}$) is reached, the FSMs de-assert the de-serializer enables. After FIFO writes are complete, the data are transferred to a synchronization FIFO (Sync FIFO). This stage is required because the DRAM write width is 256 bits, whereas each Data FIFO output is 128 bits wide.

The Sync FIFO controller is triggered once all Data FIFOs reach their programmable full threshold ($N_{samp}$). It asserts the Data FIFO read enables and, one cycle later, the Sync FIFO write enables, while counting up to $4N_{samp}$ to account for de-serialization. Two Data FIFOs are read per cycle, requiring four cycles to capture data from all 64 channels, and hence the count extends to $4N_{samp}$. The selection of Data FIFO pairs is managed by a Data Multiplexer (Data MUX), which is placed between the Data FIFO outputs and the Sync FIFO and introduces a single-cycle latency to maintain functional correctness.

Reads from the Data FIFOs occur only when all FIFO outputs are valid, preventing glitches or dropped words between the Data FIFOs and the Sync FIFO. For a 73-μs imaging time corresponding to an imaging depth of approximately 56 mm, Data FIFO readout requires 29 μs after the FIFOs are filled. The Sync FIFO output is connected to the DRAM arbiter. When the Sync FIFO is non-empty, the arbiter reads a single 256-bit entry and asserts the DRAM write enable, waiting until the DRAM is ready to accept the data before checking the Sync FIFO again. DRAM writes require three cycles due to this handshake, whereas Sync FIFO writes take a single cycle. As a result, the Sync FIFO need only be large enough to buffer the overflow caused by the slower DRAM write speed.

Once the Sync FIFO is emptied, the full packet of $4N_{samp}$ entries has been written to DRAM, with the packet write taking 87 μs. This process repeats for subsequent beams, incrementing the DRAM packet count after each beam. In total, the dead time between beams is 116 μs for a 73 μs imaging time, enabling beam repetition approximately every 200 μs, corresponding to a beam repetition rate of 5 kHz.

When the DRAM packet count reaches the programmed value for a single slice (128 packets), the DRAM transitions to the read state. The arbiter asserts the DRAM read enable and waits until valid data appear at the DRAM output before asserting the output FIFO write enable and incrementing the DRAM read address and counter. Although simultaneous DRAM read and write operations are supported, doing so would significantly increase beam dead time, as reads require up to 20 cycles for valid data and block writes until completion. Therefore, read and write operations are performed sequentially.

After a packet is read, the DRAM asserts a transfer-ready signal to the Python interface, enabling readout from the output FIFO. The DRAM continues reading until the output FIFO contains one full packet, then pauses until the FIFO is drained. The output FIFO is accessed via an OpalKelly BTPipe construct. A custom BTPipe controller tracks the amount of data transferred and de-asserts the READY signal once the specified transfer size is reached, allowing the DRAM to refill the output FIFO. This mechanism enables a single BTPipe call from the PC to extract all data from DRAM without risking overflow or underflow.

DRAM readout takes approximately 300 μs per packet, well within the 10-s BTPipe timeout. As long as the READY signal is reasserted within this interval, the BTPipe operates reliably. The transferred data are written to disk as MATLAB-compatible .mat files rather than .csv files, as .mat files are approximately one-third the size and are read significantly faster by MATLAB. Additional FPGA outputs monitor DRAM and output FIFO status; if the transfer-ready signal is not asserted, the BTPipe is not invoked, and the program terminates.

**Host computer software**. A Python program running on the host PC orchestrates each acquisition. For each frame (or slice in 3D imaging) it loads the pre-computed scan-chains for the pixel, sub-array, and coarse μBF settings into the configuration board, triggers acquisition of all 128 beams, reads the corresponding data packet back over the output-board BTPipe, and writes the result to disk as a MATLAB-compatible .mat file. A scan-chain readback after each load flags any programming errors.

Both FPGAs are reset between frames to keep operating temperature down and to prevent any incorrect programming state from accumulating over a long run, an issue observed during testing; the small frame-rate cost is absorbed in exchange for reliable long-term operation.

The combined Verilog and Python flow achieves a frame rate of approximately 7 frames per second (fps). The largest contributor to frame time is data transfer from the output board, which requires approximately 100 ms per frame. Although this could be reduced by eliminating de-serialization, doing so would significantly increase data conversion overhead. The next largest contributor is imaging time, which takes up to 50 ms for the maximum target imaging depth of 56 mm across 128 beams. The remaining time is spent on configuration board scan-chain consistency checks and other communication overhead between the host computer and the FPGAs.

Of these contributors, only the 50 ms imaging time is intrinsic to ultrasound at the 56 mm depth used here; the remainder is supporting-electronics overhead. The ASIC itself imposes no additional limit on frame rate: the 12.8 Gbps aggregate ASIC output and the 3.6 μs scan-chain reprogramming time together support a theoretical ceiling of over 400 fps at 32 raylines per frame. The dominant bottleneck is the USB3.0 link from the FPGA to the host PC, which caps the achievable data rate at ~881 Mbps and sets the ~7 fps ceiling at 125.9 Mb per frame. Two further constraints, transferring the raw 10-bit samples without compression and a Verilog/Python control flow that reprograms the array far more slowly than the 3.6 μs scan-chain itself allows, would surface once this link is widened. Closing the gap to the ASIC's capability is, therefore, primarily a matter of replacing the host interface with a higher-throughput link, with on-board compression and faster array reprogramming as the next steps.

**Supplementary Discussion S5. Additional Packaging Information**

In this section, we provide some additional details on the packaging techniques employed in FlexUS, supplementing that presented in **Methods.**

**ACF bonding of the piezocomposites**. In this work, direct integration of the transducers onto the ASIC was selected to preserve the overall flexibility of the wearable patch, as the use of interposers would significantly constrain mechanical compliance. Anisotropic conductive film (ACF) was chosen to establish the electrical interconnects. ACF is a directionally conductive material that provides electrical conduction primarily in the vertical direction while remaining insulating laterally. It consists of metal-coated polymer spheres, typically a few micrometers in radius, dispersed within an adhesive resin.

Electrical connection is achieved by placing the ACF between protruding electrodes on the two components to be bonded (**Fig. S26a**), followed by the application of heat and pressure. During thermo-compression bonding (**Fig. S26b**), the polymer spheres trapped between opposing electrodes are compressed and flattened, rupturing the polymer core and leaving the conductive metal coating to form an electrical pathway. Simultaneously, the resin flows outward from the electrode regions into the spaces between adjacent electrodes, maintaining mechanical adhesion between the bonded parts. To prevent unintended shorts, the combined thickness of the opposing electrode features must exceed the diameter of the ACF particles.

The choice of ACF particle density involves a trade-off. Higher particle densities increase the likelihood that a sufficient number of particles will contact each electrode, thereby reducing contact resistance—a critical consideration for ultrasound applications, where minimizing loss between the transceiver and transducer is essential. However, higher densities also raise the probability of electrical shorts between neighboring electrodes. For this work, 3T-TFA22020-08 (H&S Hightech, South Korea) was used. This ACF consists of 5 μm gold-coated polymer particles embedded in an 8 μm thick resin, with a particle density of approximately 5,000 particles per $mm^2$.

The effectiveness of the bonding profile used – 165°C for 30 seconds – was characterized by cross-sectioning the device (see **Methods**) and imaging using scanning electron microscopy (SEM). False-color SEM images of regions outside (**Fig. S26c**) and inside (**Fig. S26d**) the electrode intrusions show intact and flattened conductive spheres respectively.

**ENEPIG plating**. The ASIC pads consist of 1.6 μm–deep openings in the passivation layer that expose the top metal. Consequently, the electrodes above the front-end array must be built up to enable reliable electrical contact using ACF. Achieving sufficient metal thickness to rupture the 5 μm ACF particles is not feasible with standard electron-beam evaporation or sputtering and therefore requires a plating-based approach. Although electroplating could provide the necessary thickness, it requires electrical biasing of the pads, which is incompatible with the single-ended electrode structure of the FlexUS ASIC. To simplify processing, electroless plating was used, specifically electroless nickel, electroless palladium, and immersion gold (ENEPIG), with target thicknesses of 7 μm for nickel, 0.2 μm for palladium, and 50 nm for gold.

Electroless plating coats all exposed metal surfaces, including the wire-bond pads. ENEPIG was therefore selected to ensure compatibility with both ACF bonding and preceding wire bonding. The palladium layer serves as a diffusion barrier and protects the nickel from corrosion and degradation at elevated temperatures, which are encountered during both wire bonding and ACF thermo-compression. Performing this plating at the wafer level improves yield and throughput, and profilometer measurements (P-17 Stylus Profiler, KLA Corp., USA) of the ASIC surface in the electrode array region after plating (**Fig. S27**) confirm the height of the build-up (5.5 μm) to be larger than the diameter of the ACF conductive particles (5 μm).

Following bonding of the piezocomposite to the ASIC, a top-side electrical contact is required. To simplify fabrication and biasing, a shared common electrode was implemented across all pixels. This approach provides a low-resistance conductive layer over the piezocomposite while avoiding the complexity of routing individual top contacts for each pixel. Sputtering gold over the piezo edges was not practical due to the high aspect ratio at the array boundaries. In particular, the wire-bond encapsulation is located approximately 500 μm from the left and right edge columns, while the piezocomposite thickness is approximately 200 μm, resulting in a slope too steep for a thin sputtered metal layer to maintain continuity. An alternative approach—wire bonding to the top electrode at discrete points around the piezo perimeter, similar to methods used for attaching passive components to ASICs[7] - was also evaluated. However, the high surface roughness of the piezocomposite and the thin gold layer deposited on it resulted in zero yield.

The selected approach for defining the top contact was to electrically short the first and last columns of pixel electrodes to the mid-rail voltage using the ASIC top metal layer and to open the passivation above these regions. This allowed the ENEPIG process to build up sufficient metal thickness in these areas, enabling reliable top-side contact using silver paint. Although this approach sacrifices two columns of pixels, the narrow region within the inner encapsulation dam does not provide sufficient space to make a top contact to a full 32 × 32 array using silver paint. As a result, the loss of these two columns does not materially impact overall system functionality.

**Supplementary Discussion S6. Matching Layer Delay Characterization**

SU8 is typically spin-coated on flat surfaces as a photoresist, such that the thickness is quite deterministic based on the spin rate. However, the geometry of center of the FlexUS device is not a level plane. Rather, it is a well surrounded by the wirebond encapsulation that will be taller than the total thickness of the composite and matching layer combined. Hence, spinning the SU8 will not create a flat matching layer, but a concave surface with higher thickness toward the edges, where the SU8 meets the encapsulation dam. Moreover, due to the high viscosity of the SU8 and its requirement to be refrigerated, a degassing step is necessary to remove the gas bubbles caused by bringing the SU8 to room temperature. This is performed in a vacuum oven after spin-coating and an initial soft-bake, and can cause the thin layer of SU8 that is 150-200 µm thick, to "pop". The SU8 is then allowed to settle and reflow which can lead to some additional variation of the shape of the final matching layer. Two such devices are measured using a profilometer (P-17 Stylus Profiler, KLA Corp., USA) with their profiles (**Fig. S28a** and **c**) showing the aforementioned variation caused by the degassing. The profilometer has a vertical range of approximately 150 µm per scan, less than the full thickness variation across the device. To stay within this range, the composite-array region was scanned as a 4×4 grid of sub-regions, each scanned independently. The reported thicknesses are, therefore, relative within each sub-region rather than absolute across the device.

To assess whether the SU8 matching layer acts as an unintended acoustic lens, the relative thickness map is translated into a relative per-element delay map. A large delay spread across the array would indicate lensing. The thickness map for each device is interpolated to obtain the thickness at the center of each element, $t(x,y)$, and the per-element delay difference is calculated from the speeds of sound in water ($c_{water}$) and SU8 ($c_{SU8}$) as:

$$\tau(x,y) = t(x,y) * \left(\frac{1}{c_{SU8}} - \frac{1}{c_{water}}\right)$$

where x and y are the row and column of a given pixel. The delay map for each device is normalized to the delay at the central element and plotted using a conservative $c_{SU8}$ of 2000 m/s (**Fig. S28b** and **d**). For both devices the worst-case delay variation across the array stays below 50 ns, one period of the 20 MHz sampling clock, even though Device 2 has roughly twice the thickness variation of Device 1. The matching layer, therefore, behaves as a flat layer rather than as a lens. The RMS wavefront delays of 11.59 ns (Device 1) and 16.51 ns (Device 2) are also close to the Maréchal criterion[8], which holds a wavefront to be diffraction-limited when the RMS phase perturbation is below λ/14 (15.3 ns in water at the 5 MHz operating frequency). Both devices sit close to this threshold, such that the lateral-resolution penalty is small (at most 25% for Device 1 and 47% for Device 2) and no lens correction is required in the reconstructions.

**Supplementary Discussion S7. Delay-and-sum beam-forming**

Image reconstruction from the received data is accomplished using the delay-and-sum (DAS) algorithm[9,10]. The fundamental principle of DAS is to model the insonified medium as a collection of point scatterers, each of which reflects the transmitted wavefront uniformly in all directions. To estimate the reflected intensity at a given point in the imaging volume, the total propagation time must be determined. This consists of the transmit time from the array to the point, and the receive time from the point back to each array element.

From the receive perspective, the propagation distance to array element $i$, denoted $d_{RX,i}$, is simply the Euclidean distance between the point ($x_p$,$y_p$,$z_p$) and the element location ($x_i$ ,$y_i$ ,$z_i$):

$$d_{RX,i} = \sqrt{\left\{(x_p - x_i)^2 + (y_p - y_i)^2 + (z_p - z_i)^2\right\}}$$

Because each array element occupies a different spatial location $d_{RX}$ is a vector across the aperture. The transmit distance $d_{TX}$ differs from the receive distance because transmission is performed by the phased array as a whole, generating a coherent wavefront rather than independent emissions from point scatterers. Common transmit beamforming strategies include focused waves, plane waves, and diverging waves, with plane and diverging waves often preferred for rapid volumetric imaging due to their larger insonified regions.

For a focused transmit wave (**Fig. S29a**), the wavefront propagates spherically from the array toward a focal point. The transmit distance to a given point scatterer is defined as the minimum distance from the array aperture to the point, accounting for the programmed transmit delays across the elements. Specifically,

$$d_{TX} = \min_{i=1 \dots N_{elements}} \left( \frac{\Delta\tau_{TX,i}}{f_{clk}} c + d_{RX,i} \right)$$

where $\Delta\tau_{TX,i}$ is the transmit delay for element $i$, $f_{clk}$ is the clock frequency (80 MHz), and $c$ is the speed of sound in the medium. The minimum is taken because the element with the shortest geometric distance to the point is not necessarily the first to transmit. Owing to the nature of coherent transmission, the resulting $d_{TX}$ is identical for all receive elements. Equivalent formulations apply for plane and diverging wave transmissions.

Once both transmit and receive distances are known, the total round-trip propagation time for each element is computed as $(d_{TX} + d_{RX,i})/c$. This time is converted to a sample index by multiplying by the sampling frequency $f_S$. Because the resulting index is generally non-integer, linear interpolation is used between the two nearest samples. For example, a return time of $11.2T_s$, where $T_S = 1/f_S$, corresponds to samples 11 and 12, weighted by 0.8 and 0.2, respectively. Although higher-order interpolation schemes are possible, linear interpolation was selected due to its minimal computational complexity.

The interpolated samples from all array elements are then summed to obtain the reflected intensity at the selected point. This process is repeated for all points in the imaging grid and for all transmitted beams (scan lines). The resulting intensities are normalized to the maximum value and log-compressed to a target dynamic range prior to visualization. Logarithmic compression enables simultaneous visualization of highly and weakly reflective structures without saturation.

Only points within the effective aperture-defined imaging region—an hourglass-shaped volume centered about the focal point (**Fig. S29b**)—contribute coherently to the reconstruction. Points outside this region introduce incoherent noise due to the finite directivity of both individual elements and the aperture as a whole. This effect is captured through the receive F-number ($f_{\#}$), implemented by excluding array elements located outside $z/(2f_{\#})$ for a given imaging point. Typical F-numbers between 1 and 2 were used, depending on wavelength, element dimensions, and steering angle.

Applying DAS reconstruction requires access to data from each individual array element. However, due to the 16× channel-count reduction in the system, each 4×4 sub-array produces a single combined data stream. Replicating this stream across all 16 elements would effectively increase the element size and spacing, resulting in grating lobes due to an effective pitch approaching 3λ. Acquiring individual element data by bypassing both fine and coarse μBFs would require 16× more acquisitions to collect data from all elements in 64-channel segments, significantly reducing frame rate. Moreover, due to the ASIC architecture, bypassing receivers necessitates disabling transmitters in the same sub-array, creating periodic inactive elements that generate grating lobes in the transmit field.

To avoid these artifacts, all pixels and sub-arrays were enabled during transmission, with only the coarse μBFs bypassed, resulting in four bypass configurations rather than sixteen. The fine μBFs remained active and performed beamforming using the transmit delay profile rather than simple averaging. This approach preserved full-aperture transmission while enabling sub-array-level reception, significantly improving image quality. Validation using data from a P4-1 probe on the Zerdine phantom's volume targets confirmed that the 4× channel-count reduction did not measurably degrade image quality. The reconstructed images with conventional DAS (**Fig. S30a**), i.e., using all pixels to beamform, and using the sub-array beamforming methodology presented in this work (**Fig. S30b**) are quite similar, with performance degrading only at larger angles past ±30°, which will not be used with the FlexUS device. Since the P4-1 is a linear probe, only a one-dimensional equivalent of the sub-array beamformer was applied, with every two elements forming a sub-array. However, to a first-order, this confirms the validity of the beamforming approach used since the element pitch of the P4-1 probe is comparable to that of the FlexUS device, at slightly over half a wavelength. Accordingly, all experiments were conducted using four acquisitions per transmitted beam—one for each coarse μBF bypass configuration. To adequately sample the imaging volume, 32 beams spanning a ±30° sector were used per frame or slice, resulting in 128 acquisitions per slice.

The DAS algorithm was implemented in MATLAB using nested for-loops. The outer loop iterates over beams, followed by a loop over spatial grid points. For each point, return times, sample indices, and interpolation weights are computed as vectors across all array elements. The innermost loop performs the summation of interpolated samples to compute the reflected intensity. Although matrix-based implementations such as those used in the MATLAB Ultrasound Toolbox (MUST)[10] are often assumed to be faster, the explicit for-loop implementation consistently achieved better performance due to MATLAB's internal optimization behavior.

**Supplementary Discussion S8. Wireless configuration board**

The wireless version of the configuration board (**Fig. S31**) is aimed to work with a future iteration of the ASIC that includes on-chip data conversion for all 64 channels, which would eliminate the need for the output board. Consequently, the functionality of both support boards would be performed by the configuration board alone, with its XEM7310 FPGA module (OpalKelly, USA) having to be replaced by the FPGA module on the output board, XEM7360 (OpalKelly, USA) due to its larger DRAM storage. This configuration board makes the system wireless as it is powered by a 4.2V 6000mAH Lithium Polymer (LiPo) battery, LP906090JH (Jauch Quartz, Germany). The battery allows up to 6A discharge current, which is necessary to drive the FPGA module, FlexUS ASIC and all relevant power regulators. With the change of the input voltage from 5V on the previous configuration board to 4.2V, all buck and boost converters need to be swapped for versions that can handle the lower input voltage. XT60 connectors were soldered to the configuration board and battery to allow easy swapping of a drained battery for a fully-charged one. With these changes, the configuration board can be powered wirelessly for up to an hour depending on the number of acquisitions run and whether they were 2D or 3D. The battery life based on typical operation is around 30 minutes.

The main limitation of the current wireless system is that a system-on-module (SoM) component like a Raspberry Pi (Raspberry Pi Foundation, UK) is needed to utilize the OpalKelly constructs like BTPipe since the FPGA module requires a USB3.0 connection. In addition, the large amount of data coming from the ASIC cannot be transmitted through WiFi without being reconstructed due to the bandwidth limits on WiFi. A Raspberry Pi would be able to perform the beamforming operation while utilizing the OpalKelly constructs and allowing for wireless communication, and was hence placed on the wireless configuration board. However, the additional power requirements of a full SoM, which is typically in the 2A-3A range would not be compatible with most LiPo batteries, and a lower power alternative is required to continue development of the wireless configuration board.

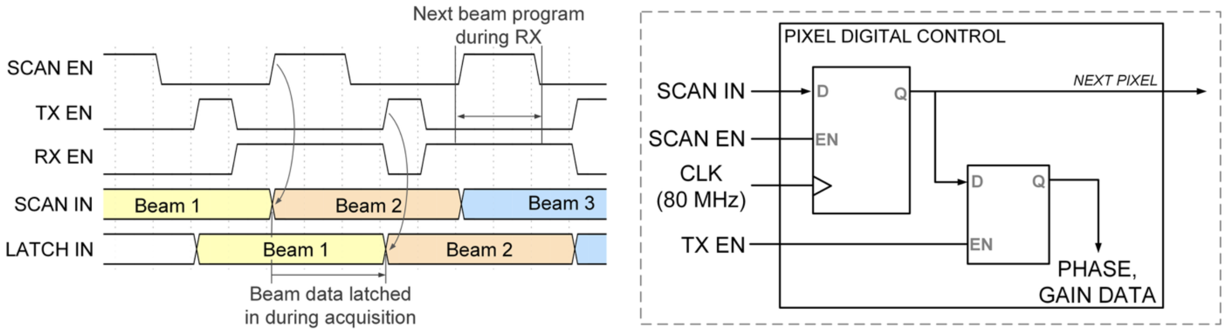


**Fig. S1. Real-time ASIC scan-chain programming a)** Timing diagram showing scan-chain programming during receive phase that is latched during the successive transmit phase for no dead time between acquisitions **b)** Scan-chain flip-flop and latch connectivity to eliminate dead time between successive scan lines

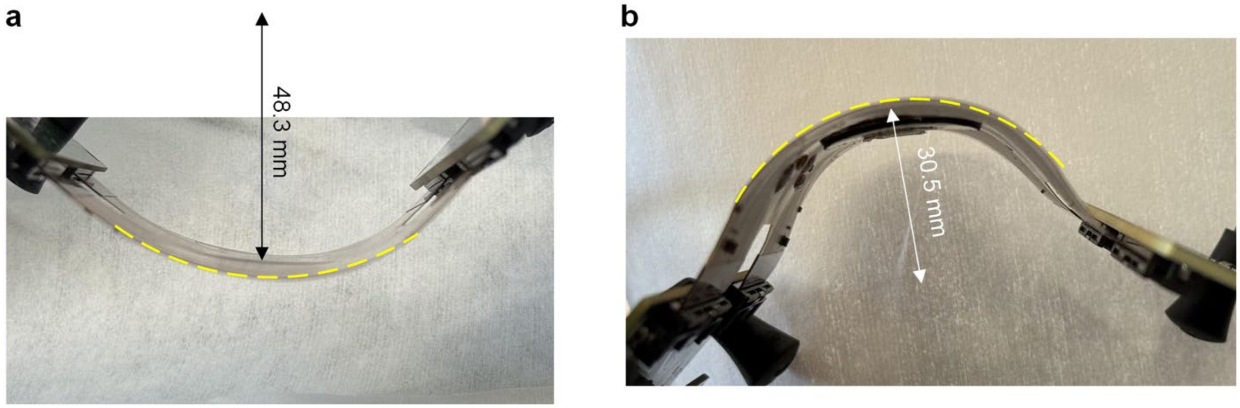


**Fig. S2. FlexUS radius of curvature estimation a)** Maximum flexion in convex mode before the connection to the ASIC is lost due to wirebond stress on the connection of the ASIC to the flexible printed-circuit board, with annotated radius of curvature at the array. **b)** Maximum flexion in concave mode before the connection to the ASIC is lost due to wirebond stress, with annotated radius of curvature at the array.

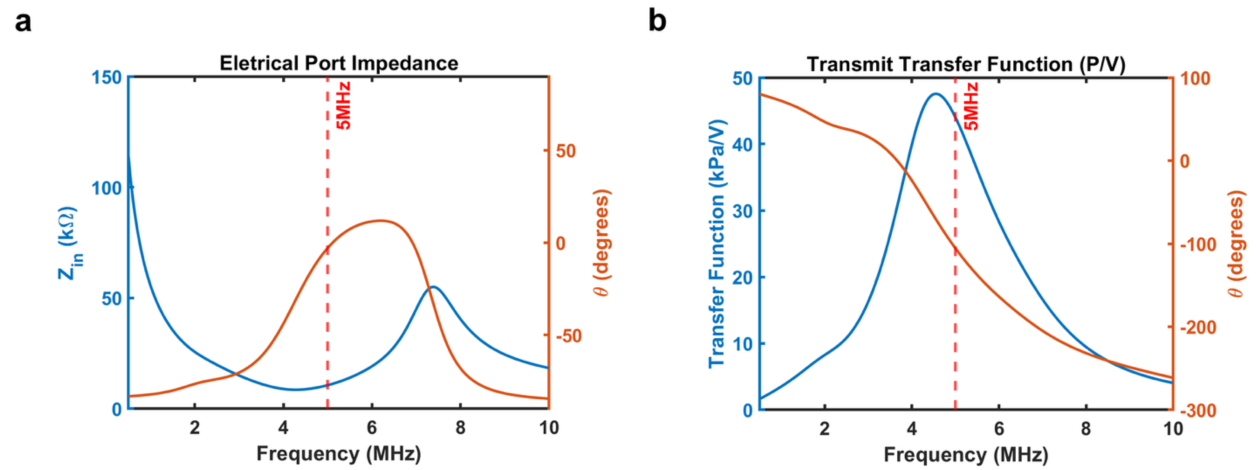


**Fig. S3. KLM simulation results of the acoustic material stack in Table S3 a)** Simulated electrical port impedance vs frequency with electrical resonance frequency (5 MHz) marked as the frequency with zero reactance **b)** Simulated transmit transfer function vs frequency with electrical resonance frequency marked to show difference between electrical resonance and mechanical resonance (peak transmit transfer function)

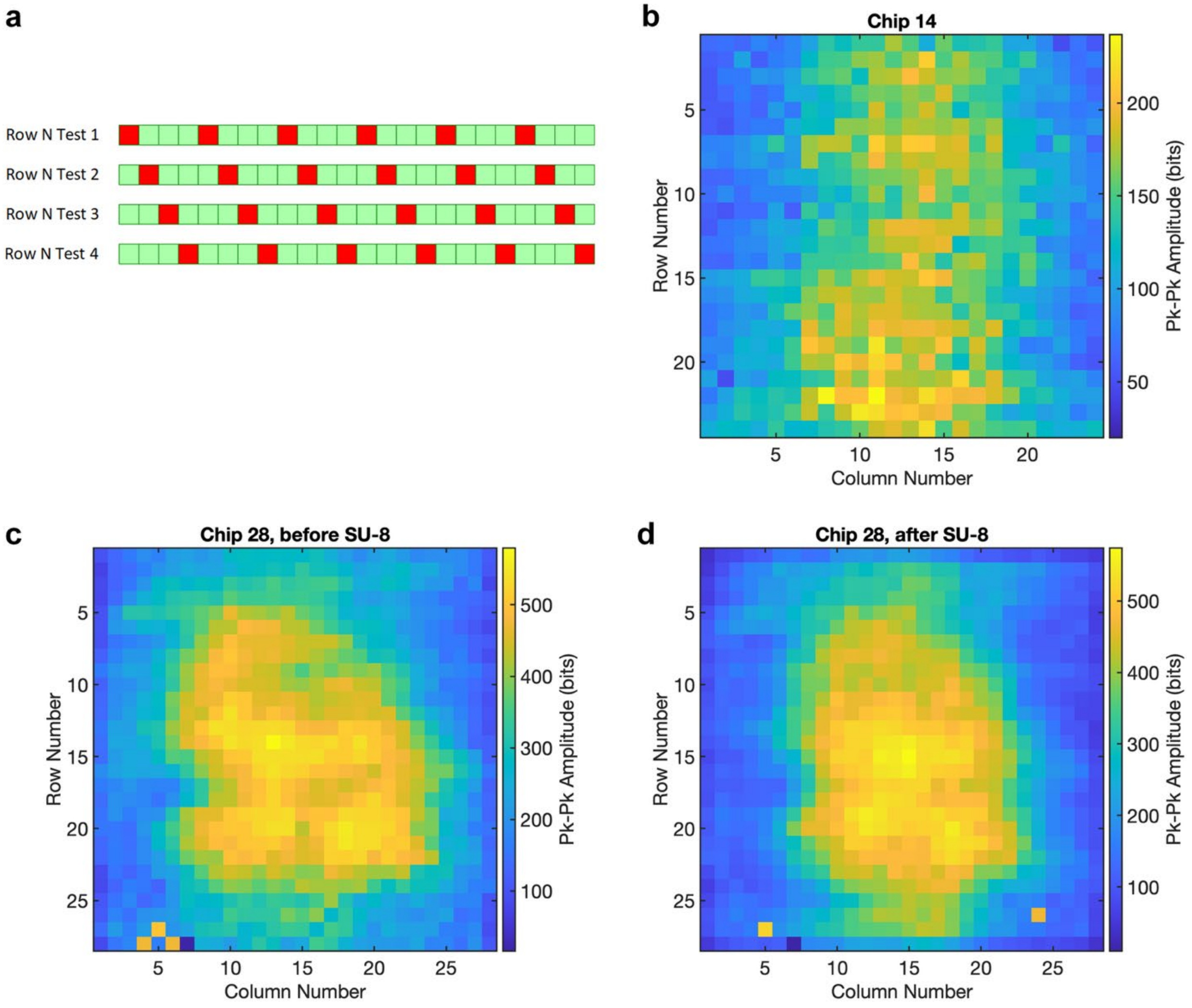


**Fig. S4. Plane-wave test for ACF bonding characterization a)** Pixel stimulation pattern for a single row in the plane-wave test showing the 4 different stimuli configurations to encompass all pixels **b)** Plane-wave test results for row-by-row stimulation before optimization of the fabrication parameters **c)** Plane-wave test results for row-by-row stimulation after optimization of the fabrication parameters with higher overall amplitude indicating better ACF connectivity **d)** Plane-wave test results for row-by-row stimulation of the same device after SU8 deposition and curing showing a more symmetric pattern with the matching layer

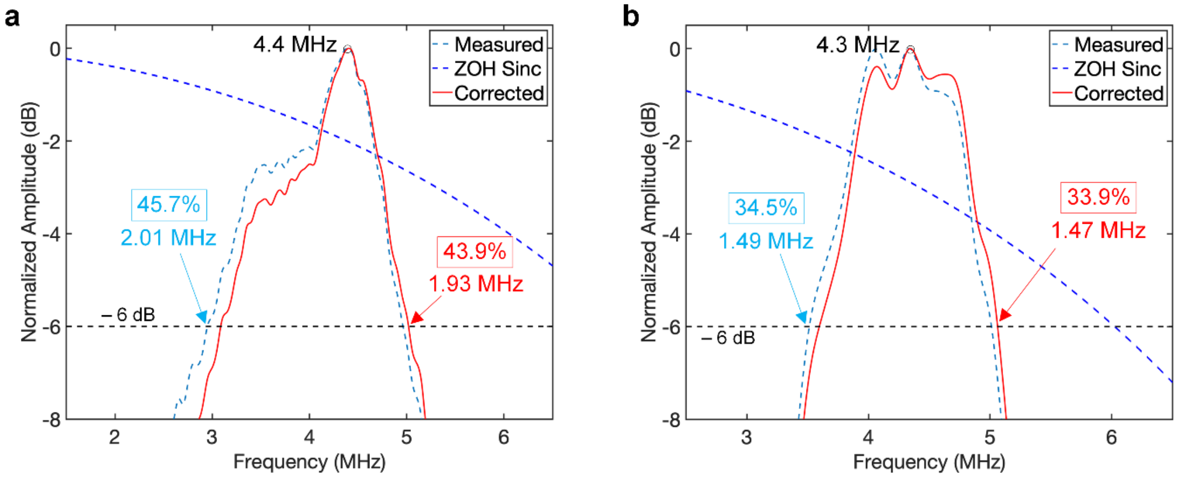


**Fig. S5. System bandwidth measurements a)** FFT of the transmitted single-pulse at 6 MHz measured with the hydrophone along with corrected spectrum accounting for the ZOH effect of the pulse duration **b)** FFT of the pulse-echo signal amplitude as measured at the output board for a wire phantom target along with the corrected spectrum

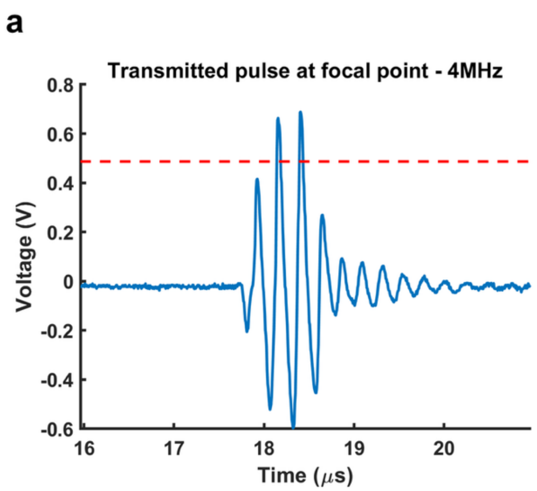


**Fig. S6. Acoustic pulses measured with hydrophone a)** 3-period pulse stimulus to transmitters at 4 MHz center frequency with half-power (3 dB) voltage annotated (in red) to determine transmitted pulse count **b)** 3-period pulse stimulus to transmitters at 5 MHz center frequency with half-power (3 dB) voltage annotated to determine transmitted pulse count

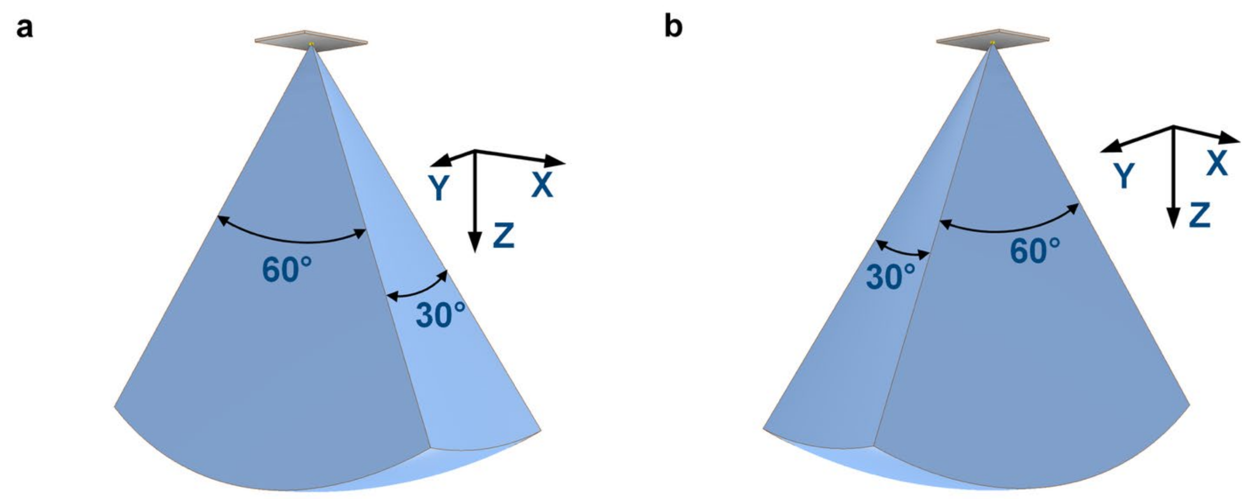


**Fig. S7. 3-D imaging modes of FlexUS system a)** XZ-centric 3D imaging with azimuth (±30º) in the XZ plane and elevation (±15º) in the YZ plane **b)** YZ-centric 3D imaging with azimuth (±30º) in the YZ plane and elevation (±15º) in the XZ plane

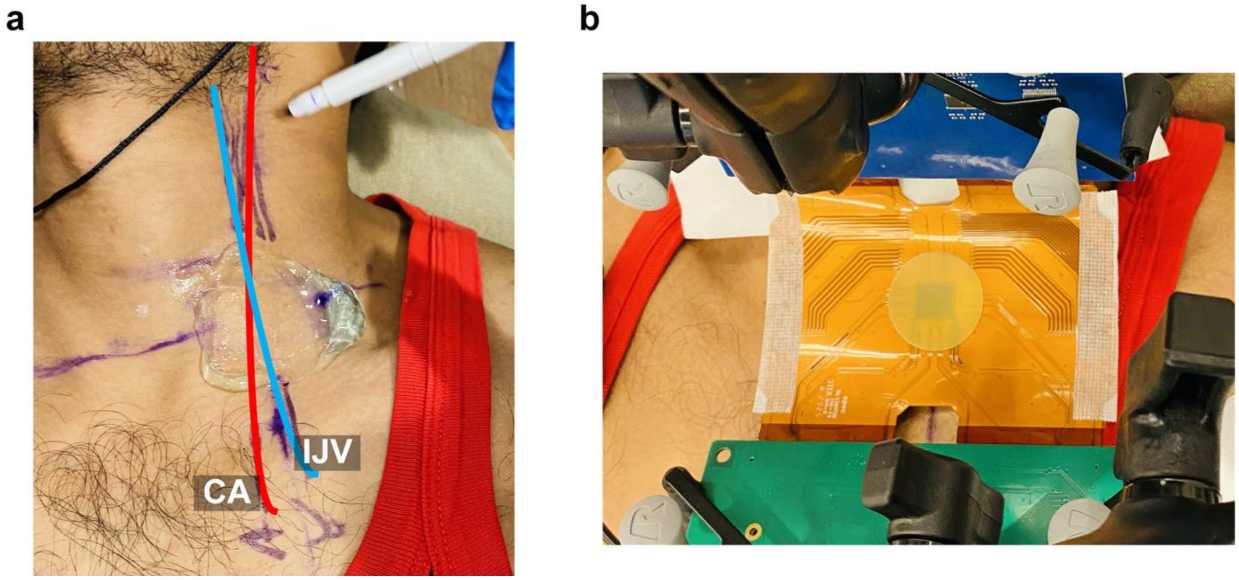


**Fig. S8. Positioning for imaging neck vasculature a)** Cross-hairs for length-axis positioning of the FlexUS device on the subject with their neck hyperextended, along length of carotid artery (in red) and IJV (in blue). Gel was used for conventional imaging with a commercial probe. **b)** FlexUS device placed on the IJV length-axis cross-hairs with its X-axis along the length-axis of the IJV

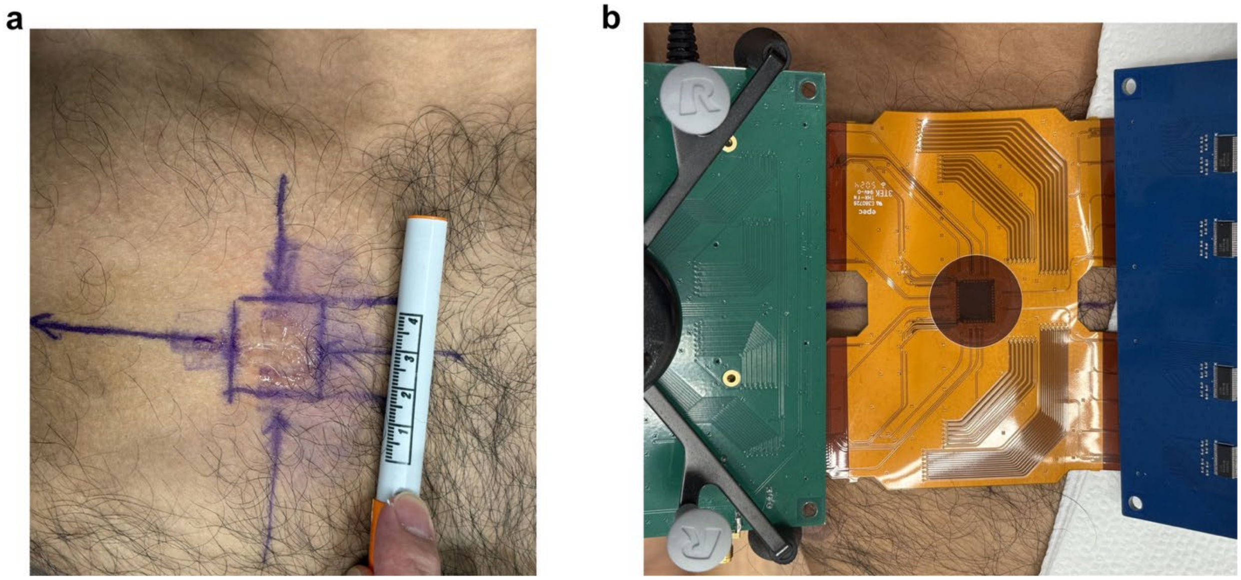


**Fig. S9. Positioning for lung imaging a)** Cross-hairs for positioning the FlexUS device on the right thorax of the subject based on recordings from a Philips S5-1 probe **b)** FlexUS device placed on the cross-hairs with its X-axis parallel to the subject's ribs

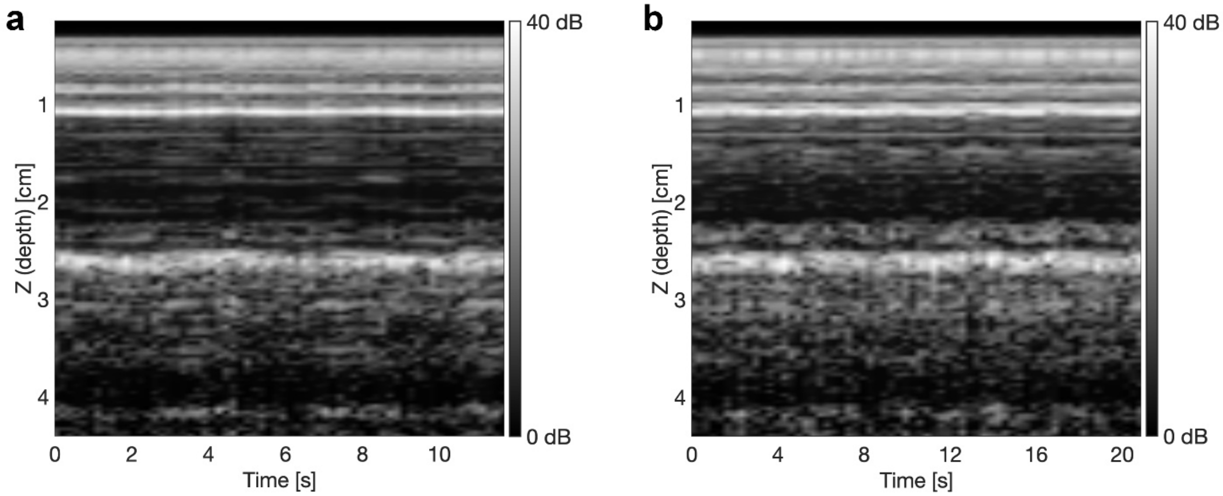


**Fig. S10. M-mode plot of the ROI in the real-time B-mode lung image displaying the sea-shore sign a)** Higher frame rate acquisition (5.4 fps) **b)** Lower frame rate acquisition (3 fps)

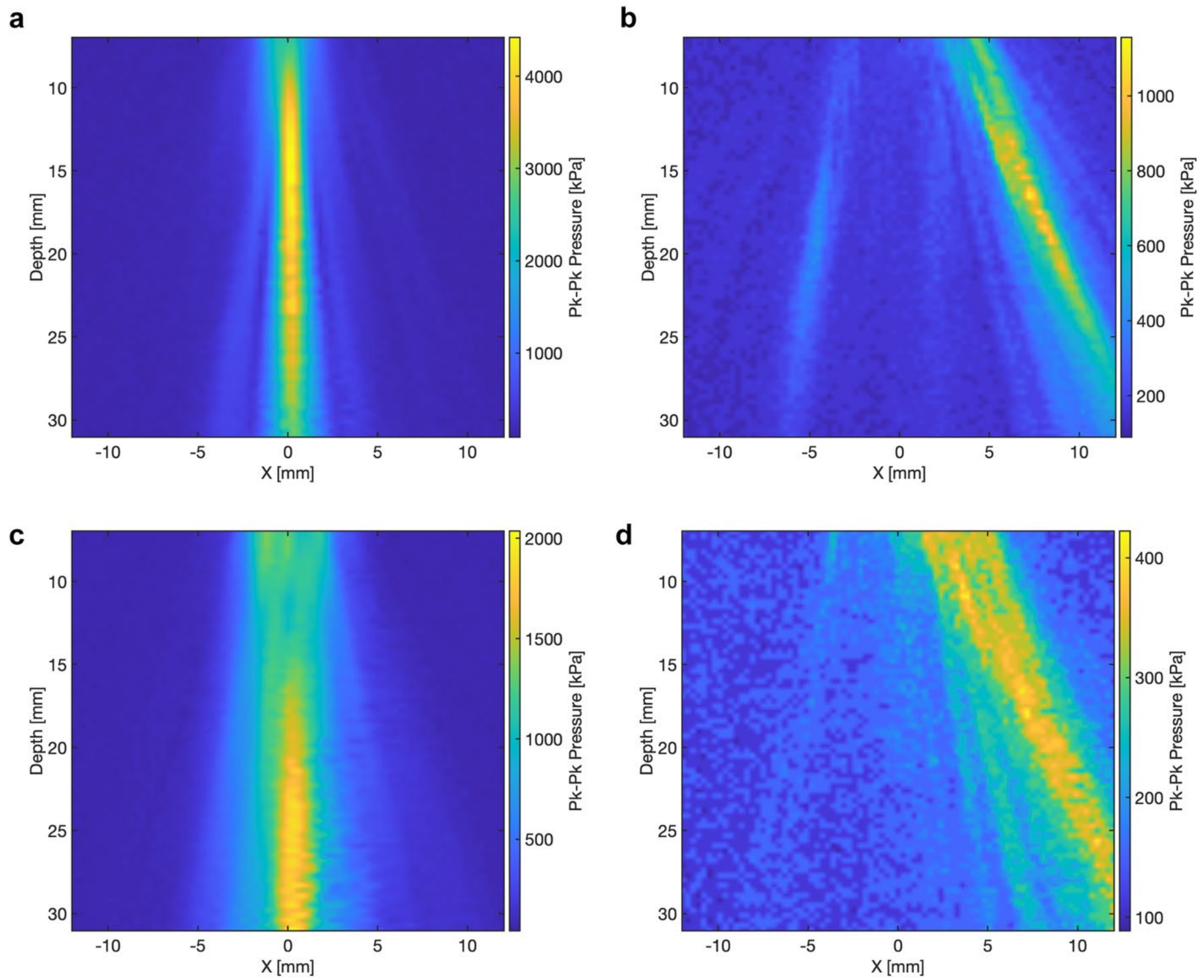


**Fig. S11. Pressure profile as measured by the hydrophone for the same device in Fig. 3 before SU8 deposition a)** Focused beam with 20mm focal depth and 0º steering along azimuth **b)** Focused beam with 20mm focal depth and 22.5º steering along azimuth **c)** Plane wave with 0º steering along azimuth **d)** Plane wave with 22.5º steering along azimuth

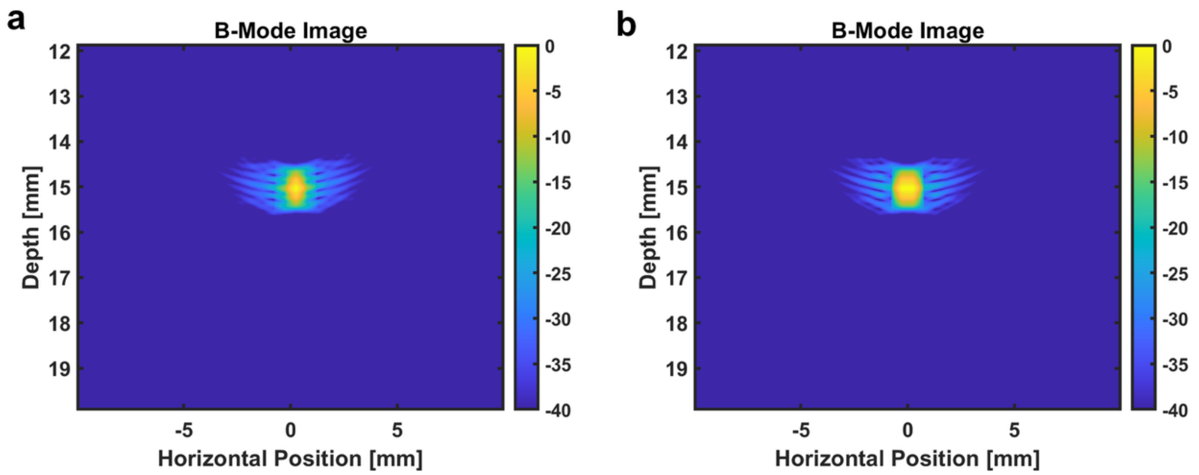


**Fig. S12. Field-II simulation for resolution a)** B-mode image for point scatterer at 15mm depth for an aperture with focal distance 15mm along XZ Plane with 0º elevation **b)** B-mode image for point scatterer at 15mm depth for an aperture with focal distance 15mm along YZ Plane with 0º azimuth

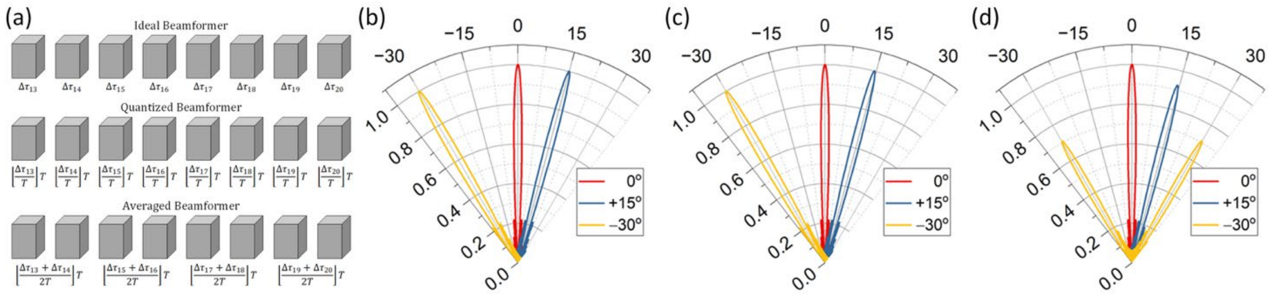


**Fig. S13. Effect of quantization and channel-averaging on beam pattern a)** Ideal, quantized at 16× the center frequency, and averaged beamformer architectures for the middle 8 elements in a 32-element linear array with corresponding pixel delays **b)** Beam profile of the 32-element ideal beamformer **c)** Beam profile of the 32-element beamformer quantized at 16x center frequency **d)** Beam profile of the 32-element beamformer with every-two-element averaging

**Delays for TX Beam-forming**

| | | | | | | | |
|---|---|---|---|---|---|---|---|
| 12 | 12 | 13 | 14 | 15 | 15 | 16 | 16 |
| 16 | 17 | 18 | 19 | 20 | 20 | 21 | 21 |
| 21 | 22 | 23 | 24 | 24 | 25 | 25 | 26 |
| 26 | 27 | 28 | 29 | 29 | 30 | 30 | 31 |
| 31 | 32 | 33 | 33 | 34 | 35 | 35 | 35 |
| 35 | 36 | 37 | 38 | 39 | 39 | 40 | 40 |
| 40 | 41 | 42 | 43 | 43 | 44 | 44 | 45 |
| 45 | 45 | 46 | 47 | 48 | 48 | 49 | 49 |

**Corresponding Pixel Delays**

| | | | | | | | |
|---|---|---|---|---|---|---|---|
| 1 | 1 | 1 | 2 | 1 | 1 | 1 | 1 |
| 5 | 6 | 6 | 7 | 6 | 6 | 6 | 6 |
| 1 | 2 | 1 | 2 | 1 | 2 | 1 | 2 |
| 6 | 7 | 6 | 7 | 6 | 7 | 6 | 7 |
| 1 | 2 | 1 | 1 | 1 | 2 | 1 | 1 |
| 5 | 6 | 5 | 6 | 6 | 6 | 6 | 6 |
| 1 | 2 | 1 | 2 | 1 | 2 | 1 | 2 |
| 6 | 6 | 5 | 6 | 6 | 6 | 6 | 6 |

**Corresponding Sub-array Delays**

| | | | |
|---|---|---|---|
| 11 | 12 | 14 | 15 |
| 20 | 22 | 23 | 24 |
| 30 | 32 | 33 | 34 |
| 39 | 41 | 42 | 43 |

**Fig. S14. Delay programming:** Example of beam-forming delays being divided into pixel-level and sub-array-level delays for an 8×8 section of the array focused to 20mm focal depth and steered to −30° in the azimuth and 0° in elevation

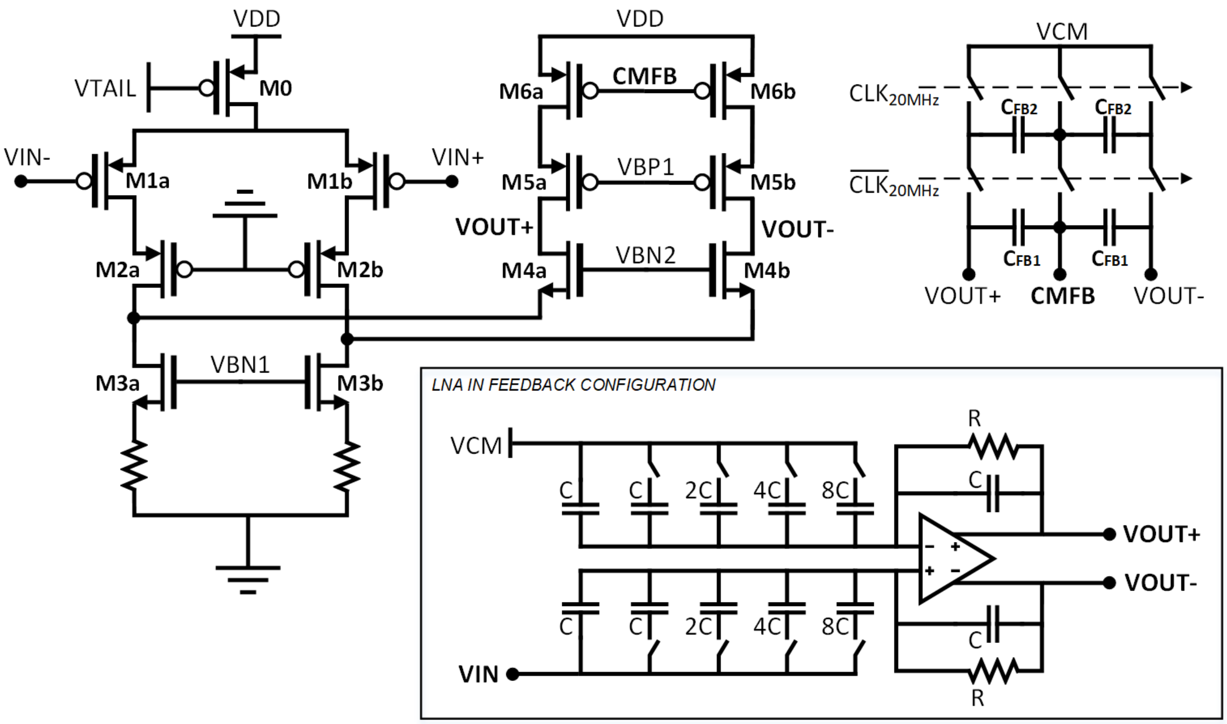


**Fig. S15. Low-Noise Amplifier:** Folded-cascode circuit schematic with capacitive-feedback closed-loop topology (bottom-right) for variable-gain, resistive-feedback for DC common-mode, and CMFB circuit (top-right)

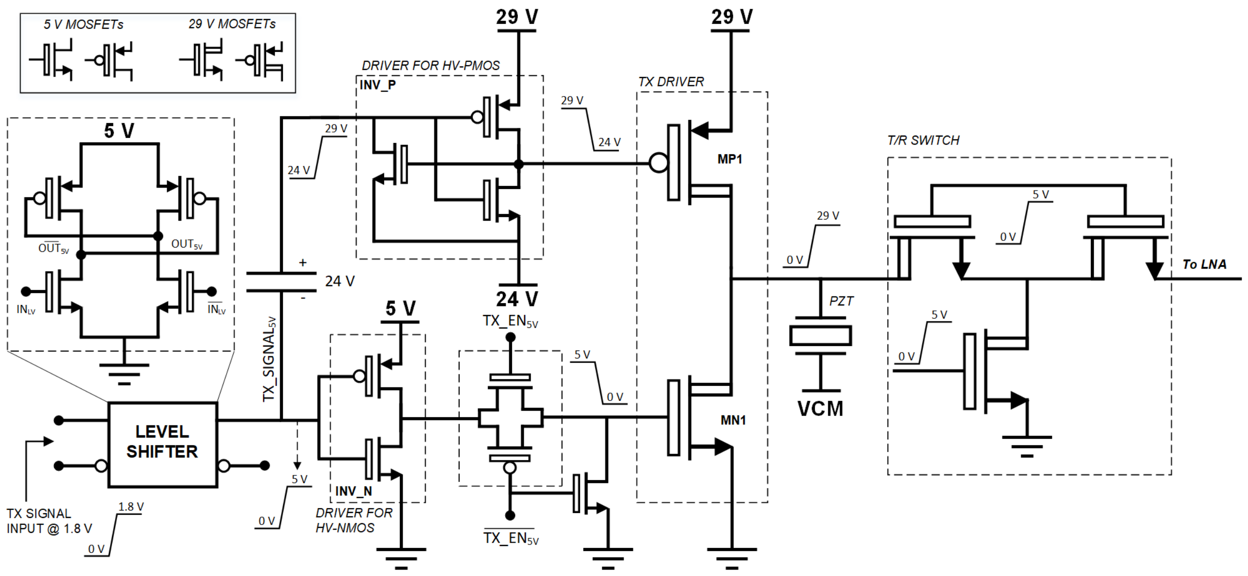


**Fig. S16. Transmitter circuit schematic including the T/R switch:** Transmitter signal path from 1.8V through necessary level-shifters to the 29V power amplifier, and double-isolation T/R switch

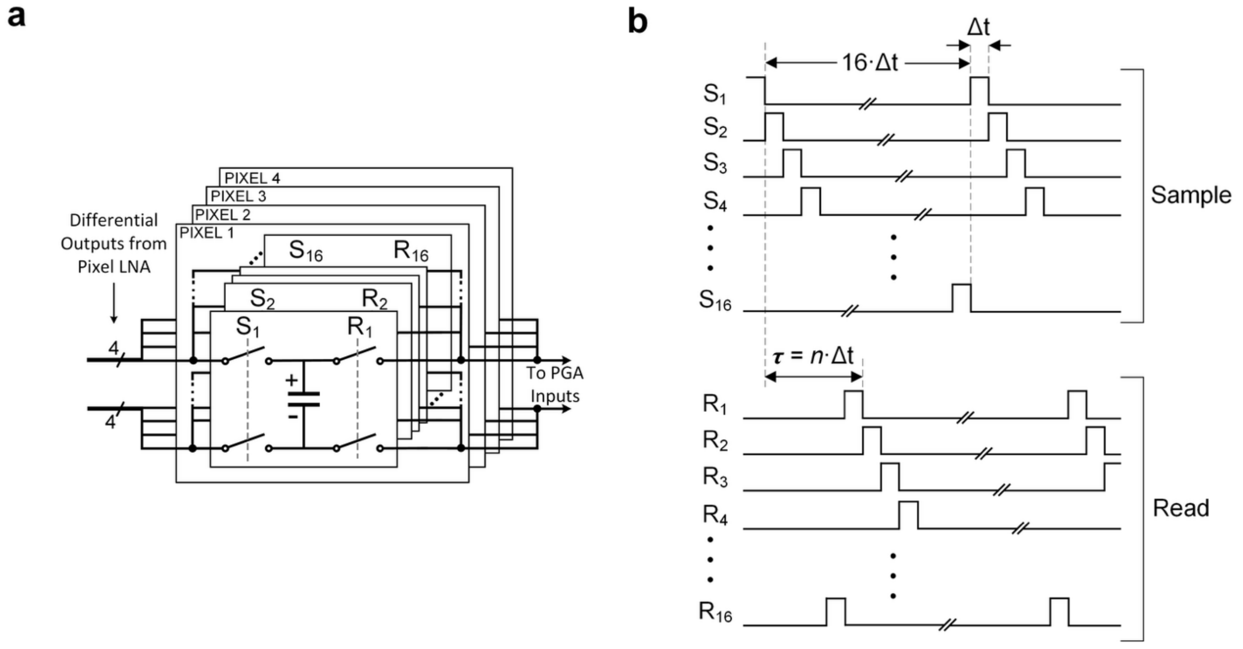


**Fig. S17. Micro-beamformer: a)** Circuit Topology for fine μBF (coarse μBF has twice as many delay units), **b)** Example waveforms for the Sample and Read Signals for the fine μBF for a pixel-level delay of $\tau = n\Delta t$ where *n* is the delay-value programmed into the pixel receiver and $\Delta t$ (= 12.5 ns) is the period of the system clock

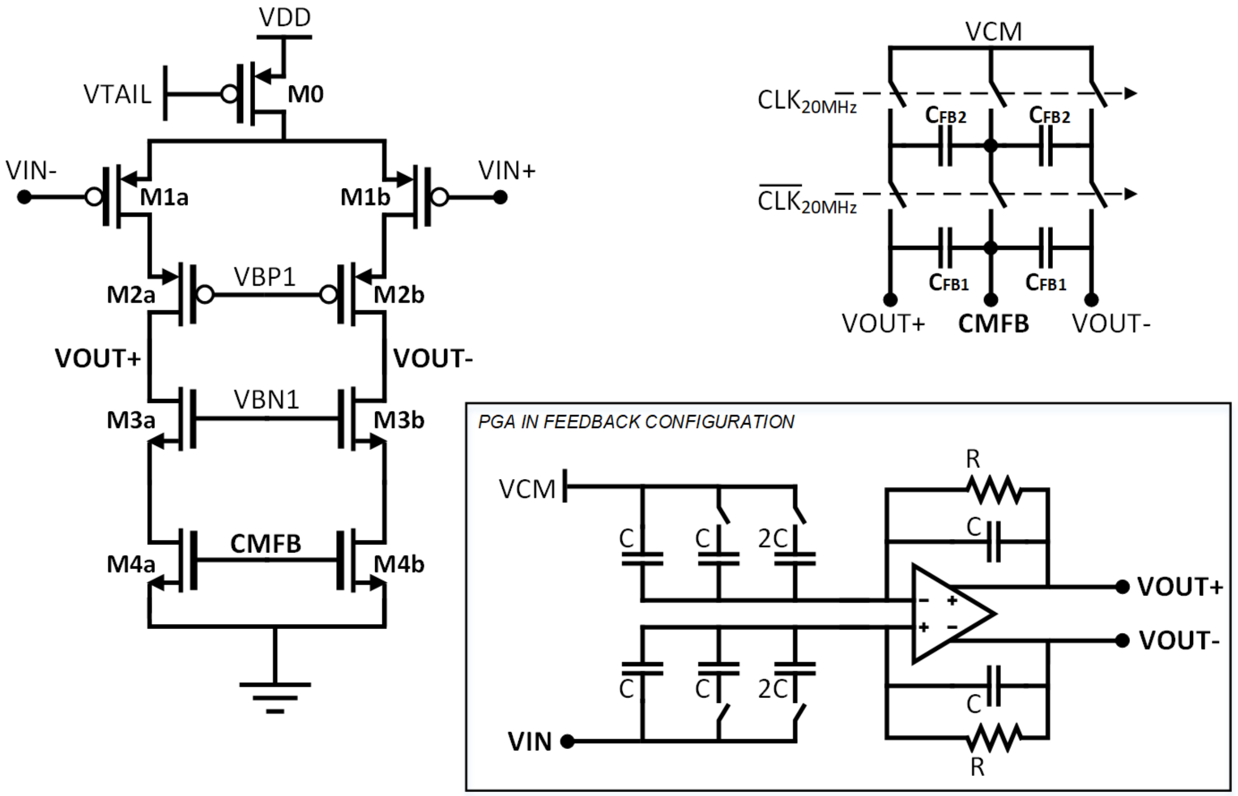


**Fig. S18. Programmable-Gain Amplifier:** Telescopic-cascode circuit schematic with capacitive-feedback closed-loop topology (bottom-right) for variable-gain, resistive-feedback for DC common-mode, and CMFB circuit (top-right)

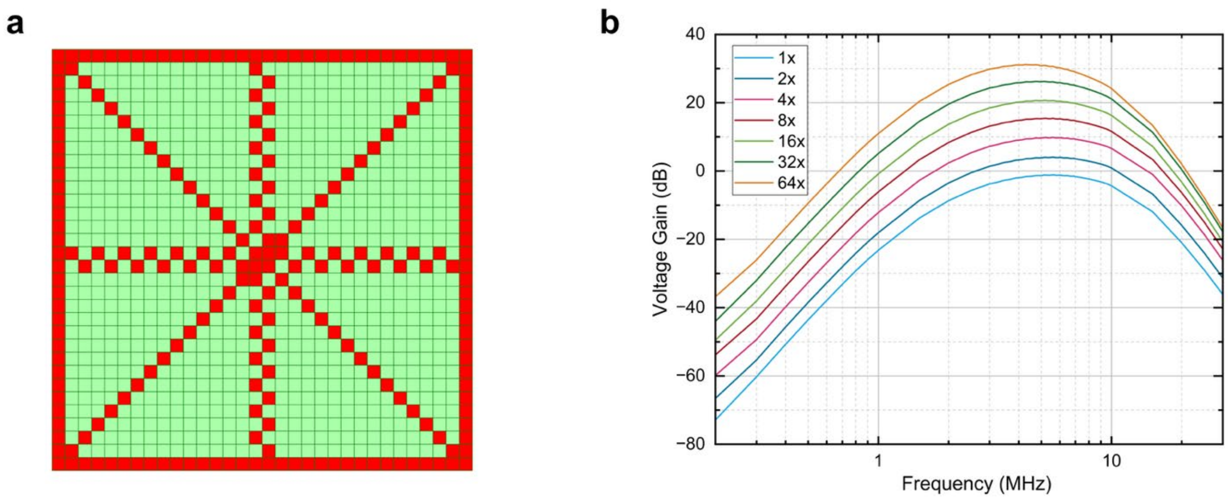


**Fig. S19. Front-end gain measurements. a)** Pixels selected (in red) out of the 32x32 array (in green) for covering all possible cases of bias and routing variation on ASIC front-end response **b)** Averaged front-end response of the pixels across an ASIC showing the designed band-pass response centered around 5 MHz

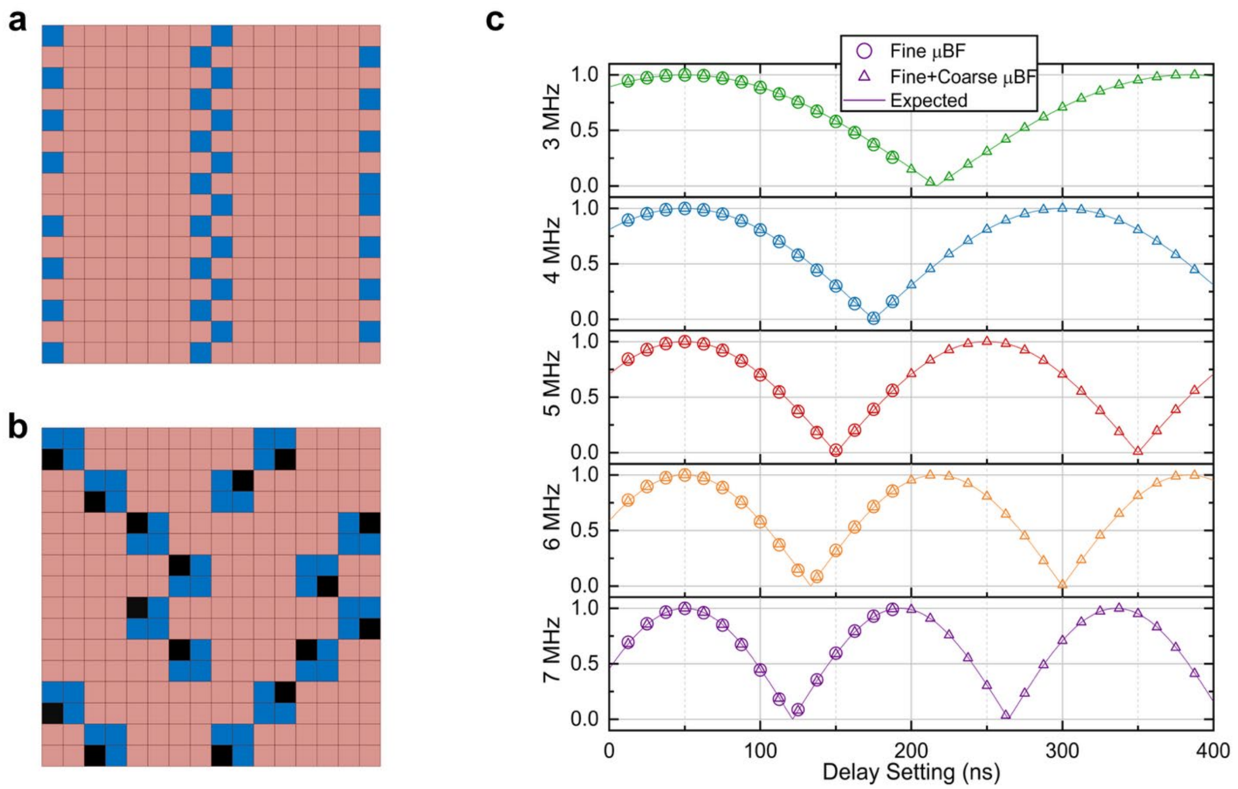


**Fig. S20. Receiver delay measurements. a)** Sub-arrays chosen (in blue) out of the 16×16 array of sub-arrays (in pink) for fine μBF phase tests **b)** 2×2 sub-array clusters chosen (in blue) out of the 16×16 array of sub-arrays (in pink) for coarse μBF phase tests with the sub-array whose delay setting is held constant highlighted for each test (in black) **c)** Averaged delay measurements for both tests compared to the ideal response obtained from MATLAB simulations for the chosen delays. The delay setting for the beamformer that is held constant for each of these tests was 50 ns (= four cycles of the system clock)

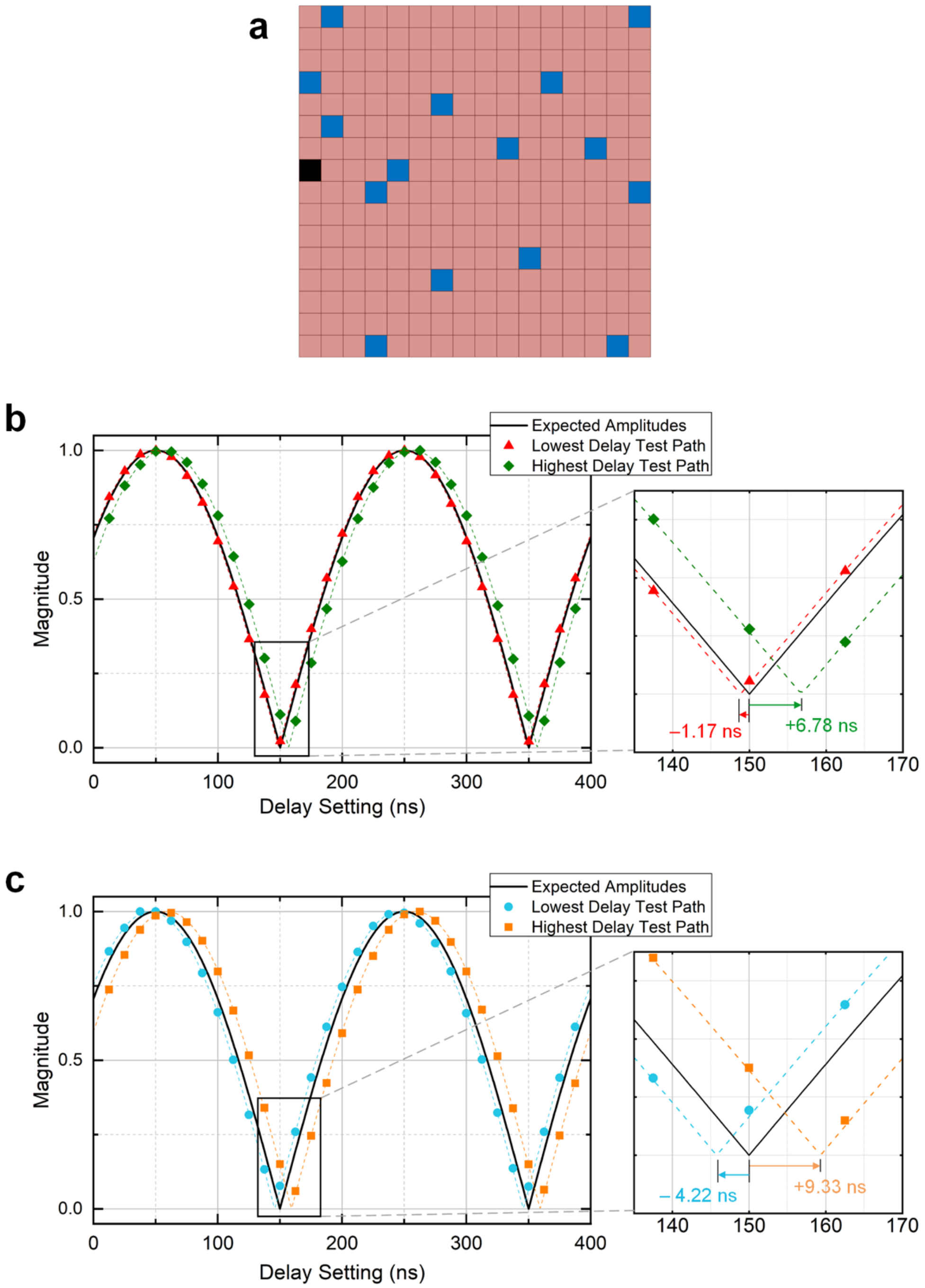


**Fig. S21. Chip-level receiver delay measurements: a)** Sub-arrays chosen for chip-level delay tests from amongst the 16×16 array of sub-arrays (pink squares) and marked as follows: black indicates the sub-array whose delay setting is held constant (at 50 ns); blue indicates the sub-arrays whose coarse delay settings are swept from 0 to maximum **b)** Magnitude of the sum of the outputs of the two channels-under-test plot against the delay setting of the second channel for the channels with minimum and maximum delays, along with the expected magnitude shown in solid black. PGA gain setting on both channels is 1x **c)** Results from the same measurement as (b) but with PGA gain varied from 1x to 4x. The delay variation in this case is larger (~13.55 ns)

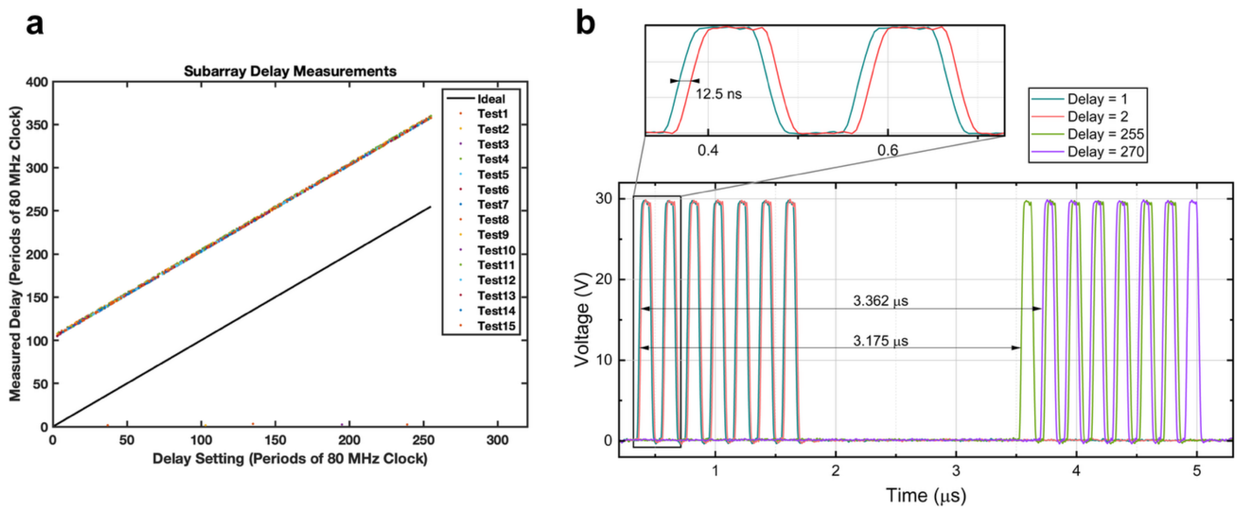


**Fig. S22. Transmitter measurements: a)** Sub-array transmitter delay test outputs for all settings against ideal performance, with the vertical offset being the pulse-width **b)** Transmit-waveforms for lowest non-zero delay between pixels (12.5ns or 1 delay setting), highest sub-array transmitter delay (3.175µs or 255 delay settings) and highest total transmitter delay (3.362µs or 270 delay settings) with an inset showing a zoomed-in waveform

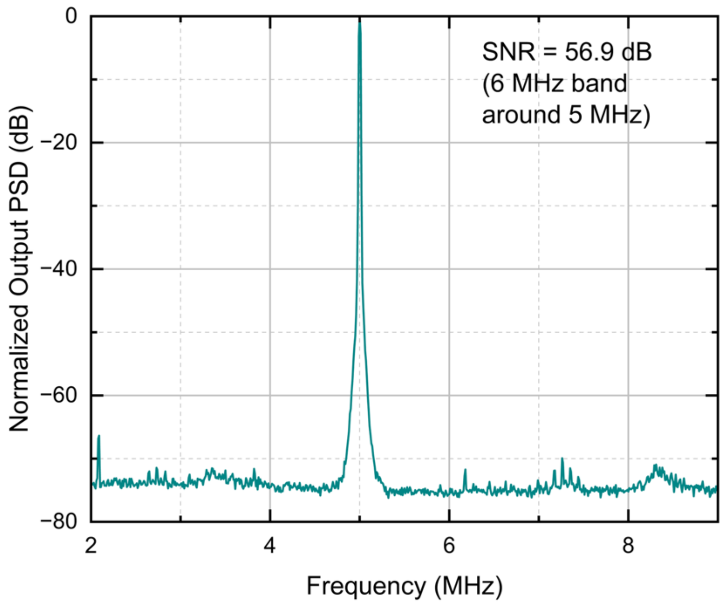


**Fig. S23. Noise measurements.** Normalized output power spectral density (PSD) for an input signal amplitude at the 1-dB compression point showing the noise floor of 11nV/√Hz and SNR of ~57 dB

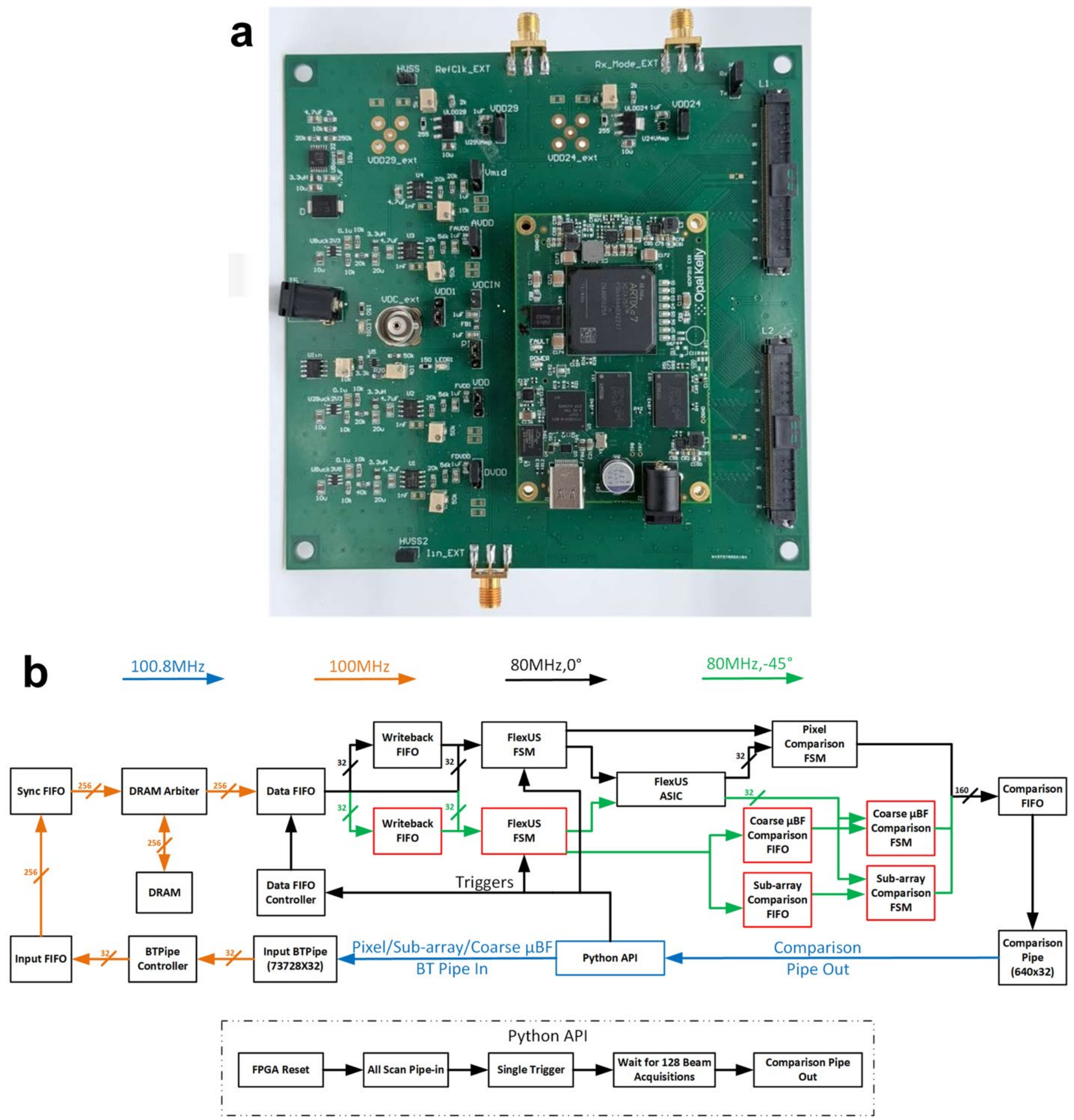


**Fig. S24. Configuration board. a)** Fully-assembled configuration board **b)** FPGA firmware implementation for scan-chain programming and software on the host computer for configuration the FlexUS ASIC showing the path of the scan-chain data from the host PC to the ASIC and back to the PC as comparison outputs

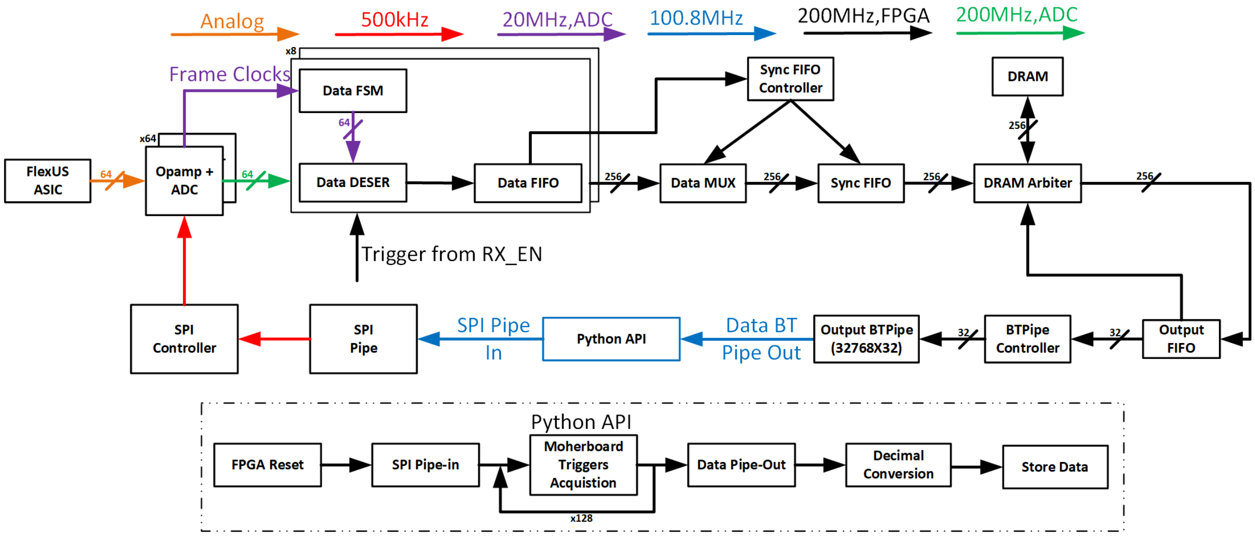


**Fig. S25. Output board FPGA firmware and host computer software** for real-time data acquisition from the FlexUS ASIC showing the modules through which the ASIC output data propagates after digitization. This includes de-serialization, buffering, storage and transfer to the PC.

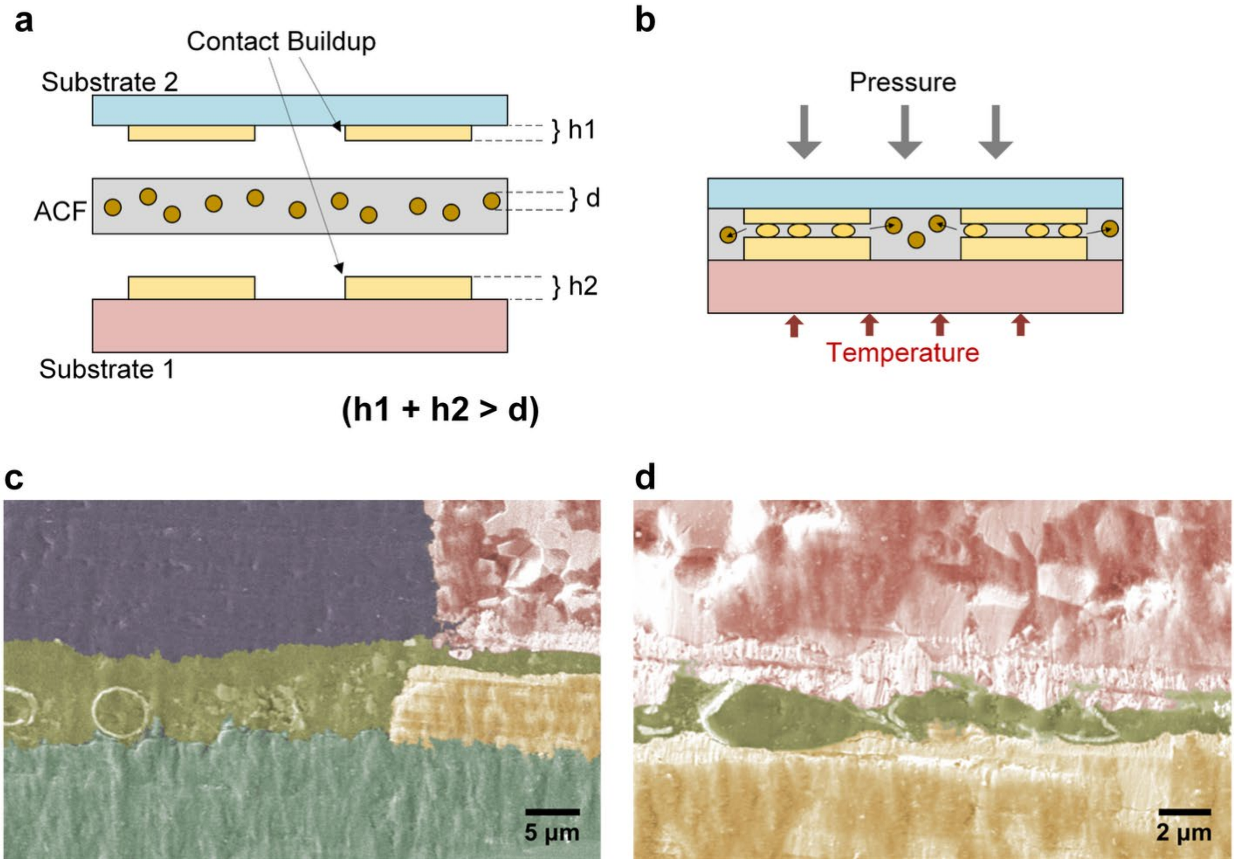


**Fig. S26. Anisotropic Conductive Film bonding a)** Material stack-up requirements for ACF bonding before application of heat and pressure **b)** Material stack-up for ACF bonding after thermo-compression **c)** False-color SEM image of ACF film showing intact conductive spheres in region outside electrode intrusion (gold) **d)** SEM image zoomed into region where conductive spheres have electrically connected gold ASIC electrodes with PZT contacts (pink)

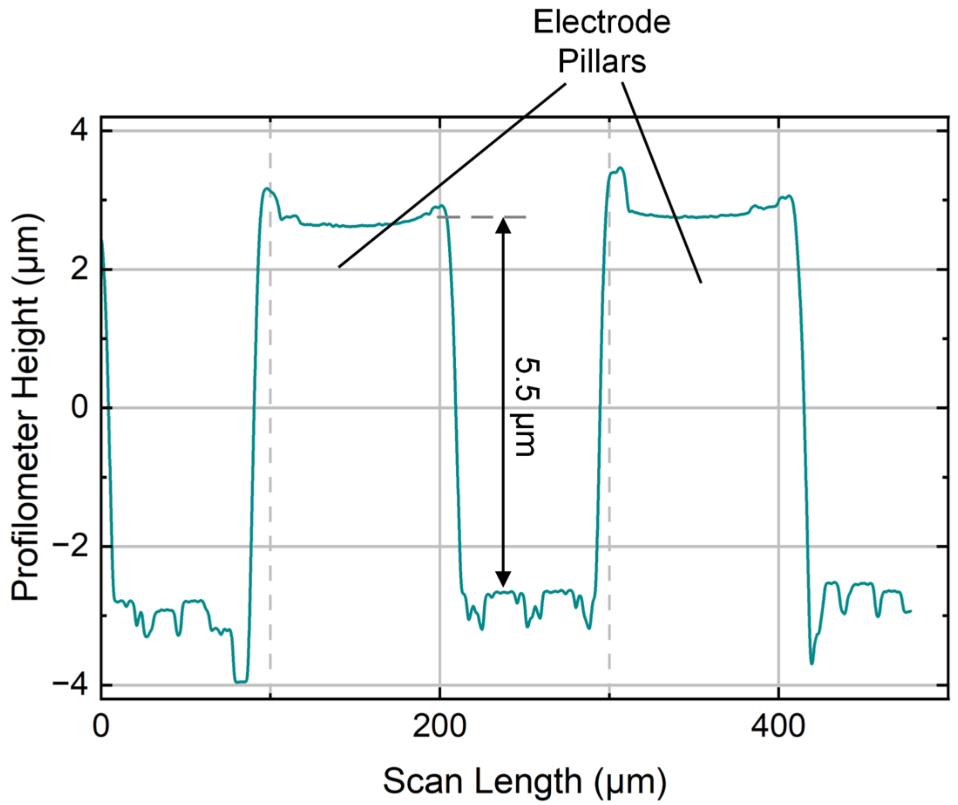


**Fig. S27. Profilometer measurements of ASIC electrodes after ENEPIG deposition** showing > 5 µm buildup over the ASIC surface. The electrodes on the ASIC as shipped from the foundry are ~1.5 µm under the surface of the passivation layer

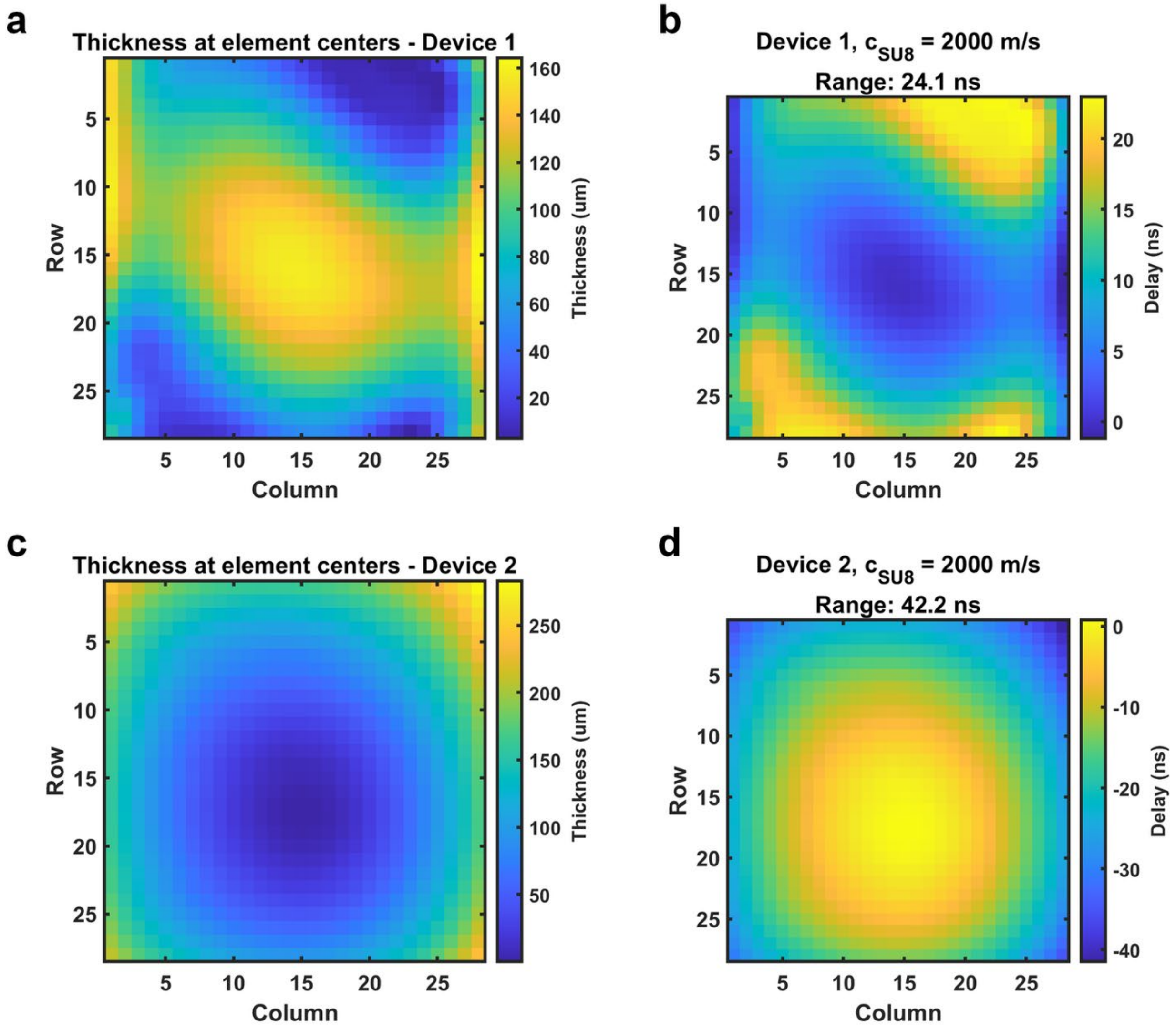


**Fig. S28. Matching Layer Characterization a)** SU8 thickness profile for the composite array region in device 1 measured with a stylus profilometer **b)** Wavefront delays caused by the shape of the matching layer for device 1 **c)** SU8 thickness profile for the composite array region in device 2 measured with a stylus profilometer **d)** Wavefront delays caused by the shape of the matching layer for device 2

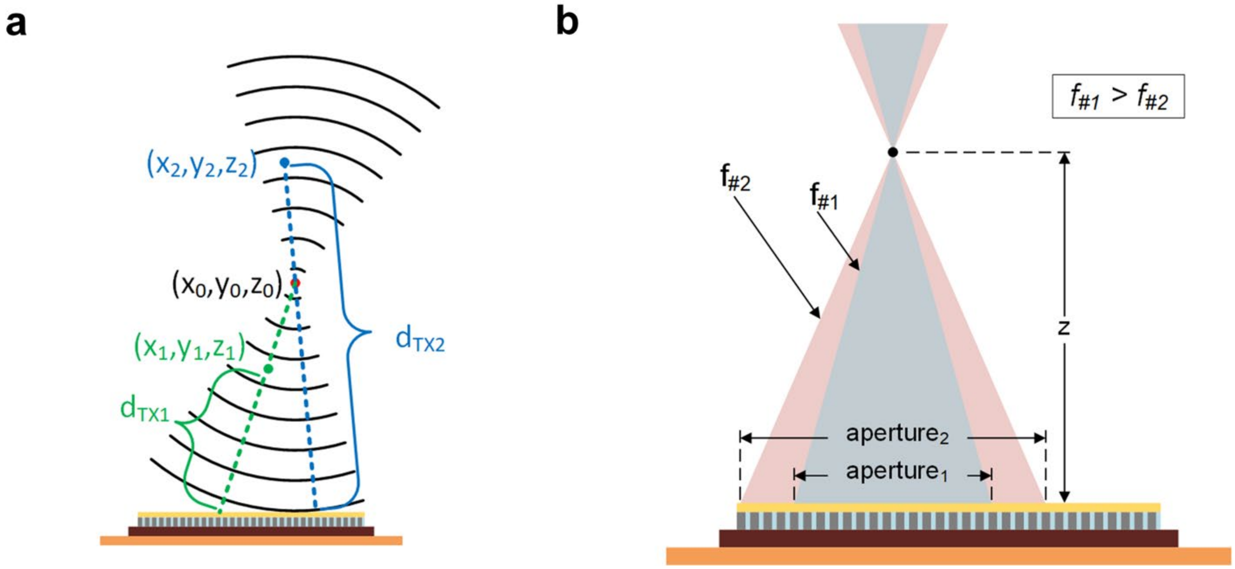


**Fig. S29. DAS reconstruction a)** Transmit distance calculation for 2 points in space insonified by a steered focused wave **b)** Example of receive aperture for a point in the image reconstruction space for two F-numbers

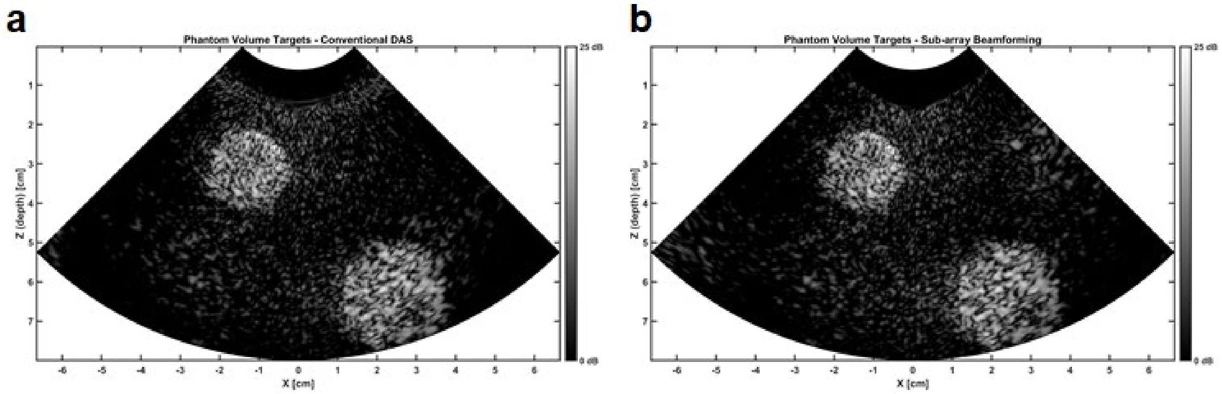


**Fig. S30. Sub-array beamforming methodology validation with P4-1 probe and Verasonics Vantage-256 system: a)** Reconstructed B-mode images using data from all 96 elements in conventional delay-and-sum **b)** Reconstructed B-mode images using the sub-array beamforming methodology presented in this work with data from 48 sub-arrays

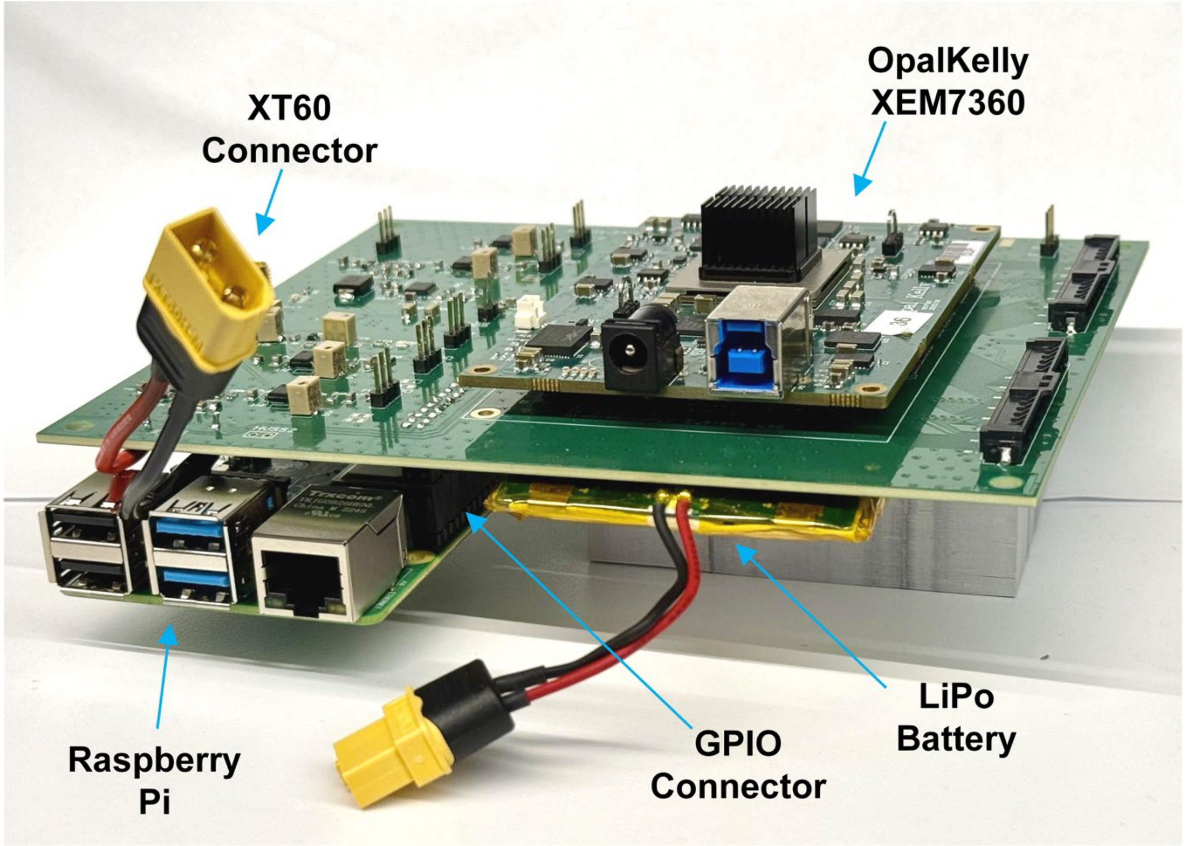


**Fig. S31. Wireless configuration board:** The wireless-capable version of the FlexUS configuration board showing the OpalKelly XEM7360 FPGA module on the top surface of the board and a Raspberry Pi 4 and lithium-ion polymer battery on the bottom surface of the board

| | FlexUS ASIC | Rosza, et al[11] | Hopf, et al[12] | Guo, et al[13] | Chen, et al[14] |
|---|---|---|---|---|---|
| **Technology** | 180 nm BCD | 180 nm BCD | 180 nm BCD | 180 nm BCD | 180 nm |
| **Transducer** | Composite PZT | CMUT | PZT | PZT | PZT |
| **Number of elements (TX/RX)** | 1024/1024 | 4096/4096 | 288/288 | 256/256 | 64/864 |
| **Center Frequency (MHz)** | 5 | 2.6 | 6 | 9 | 5 |
| **Element pitch (µm)** | 205 | 365 | 160 | 125 | 150 |
| **Sub-array Size** | 2×2 | 2×2 | 1×3 | 2×2 | 3×3 |
| **Integrated Transmitter** | Y | Y | Y | Y | N |
| **Transmit Voltage (V)** | 29 | 65 | 30 | 20 | 18 |
| **Channel Count Reduction** | 16× | 8×[a] | 18×[b] | 128×[c] | 9× |
| **Raylines/sec** | 14,000 | 7,700 | 7,000 | - | 5,000 |
| **Programming Time (µs)** | 3.6 | - | 15 | - | 0.54[d] |
| **Output Type** | Analog | Analog | Digital | Digital | Analog |
| **Delay Resolution (ns)** | 12.5 | 32 | 20.8 | 12.5 | 30 |
| **Active Area/Element ($mm^2$)** | 0.056 | 0.120 | 0.030 | 0.016 | 0.023 |
| **RX Power/Element (mW)** | 1.16 | 0.85 | 1.12 | 1.83 | 0.27 |
| **Dynamic Range (dB)** | 88 | 73 | 91 | 83 | - |
| **Peak SNR (dB)** | 57 | 52 | 52.2 | 54 | 51 |
| ***In-vivo* imaging characterization** | ✓ | ✘ | ✘ | ✘ | ✘ |

**Table S1. Comparison with state-of-the-art ultrasound ASICs.** The parameters used for comparison include various ASIC specifications such as number of elements, center frequency, element dimensions, whether a transmitter is present in each pixel, amongst others. The FlexUS ASIC, which is the only one with performance characteristics to demonstrate *in-vivo* imaging, takes the least amount of time to program, has the highest demonstrated raylines/s capability, and the highest reported SNR.

| Number of delay bits (N) | Lateral Resolution (µm) | Elevational Resolution (µm) |
|---|---|---|
| 6 | 660 | 737 |
| 5 | 660 | 737 |
| 4 | 660 | 737 |
| 3 | 660 | 737 |
| 2 | 698 | 752 |
| 1 | 746 | 774 |

**Table S2. Effect of delay quantization on lateral and elevational resolution.** The variation in lateral resolution and elevational resolution for a focused beam at focal depth 15 mm as calculated in Field-II is shown. N is the number of bits allocated to the programmable delays at the pixel level, which dictates the time-resolution of these delays.

| Material | Thickness (µm) | Longitudinal Velocity (m/s) | Density ($kg/m^3$) | Acoustic Impedance (MRayl) |
|---|---|---|---|---|
| SU-8[15-19] | 100 | 1590 | 1220 | 1.94 |
| Gold[20] | 1.2 | 3240 | 19320 | 62.60 |
| Composite PZT-5H[21,22] | 190 | 2900 | 4740 | 13.74 |
| Gold | 1.2 | 3240 | 19320 | 62.60 |
| ACF[23,24] | 8 | 1600 | 1200 | 1.92 |
| Gold | 7 | 3240 | 19320 | 62.60 |
| Silicon[20,25] | 50 | 8400 | 2329 | 19.56 |
| Polyimide[26,27] | 250 | 1646 | 1420 | 2.34 |
| FR-4 Laminate Backing[28] | 700 | 2740 | 1850 | 5.07 |

**Table S3. Material Stack:** The materials in the device cross-section are listed from the front of the device to the back along with acoustic properties.

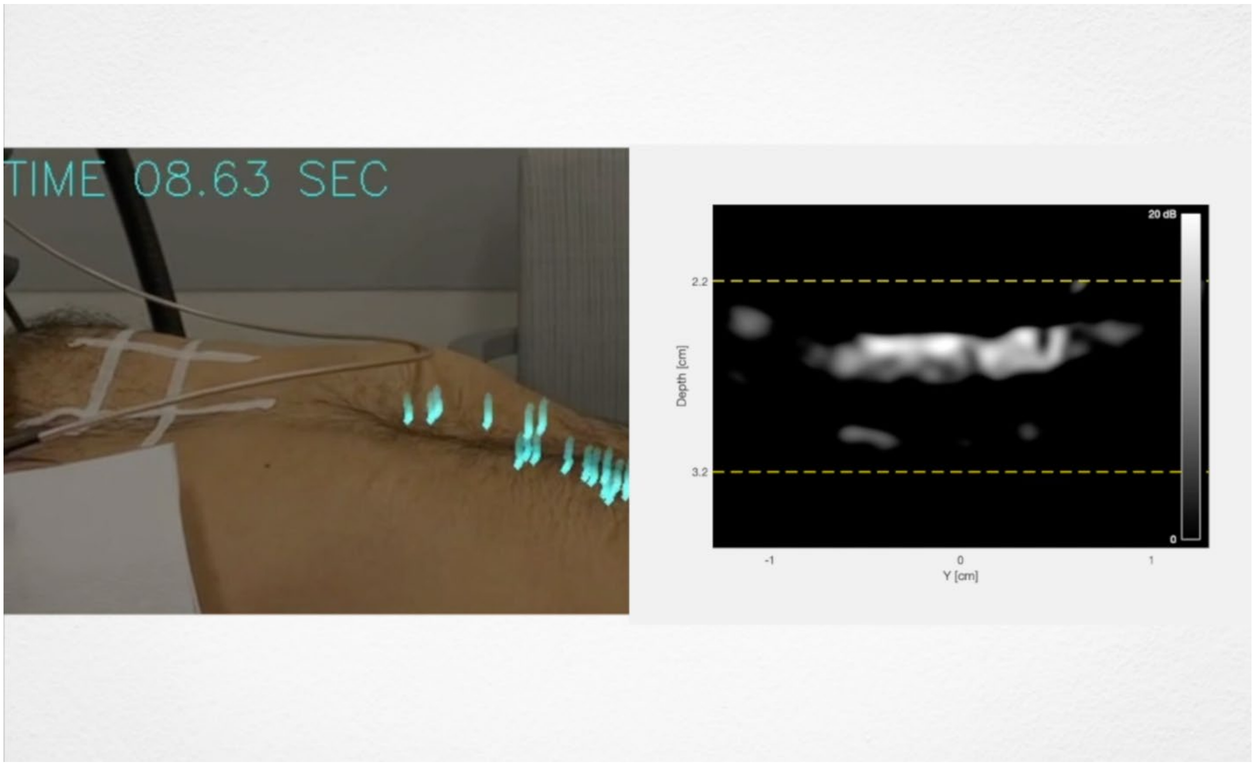


**Video S1. Chest motion alongside pleural brightness (ultrasound frame rate = 5.4 fps).** The subject's chest and abdomen are recorded (left panel) to track breathing cycles. The overlaid blue points are used for optical tracking to extract the motion of the chest against time. The real-time B-Mode image (right panel) displays the ROI (yellow box) and its brightness after Gaussian-blurring and filtering (see **Methods**)

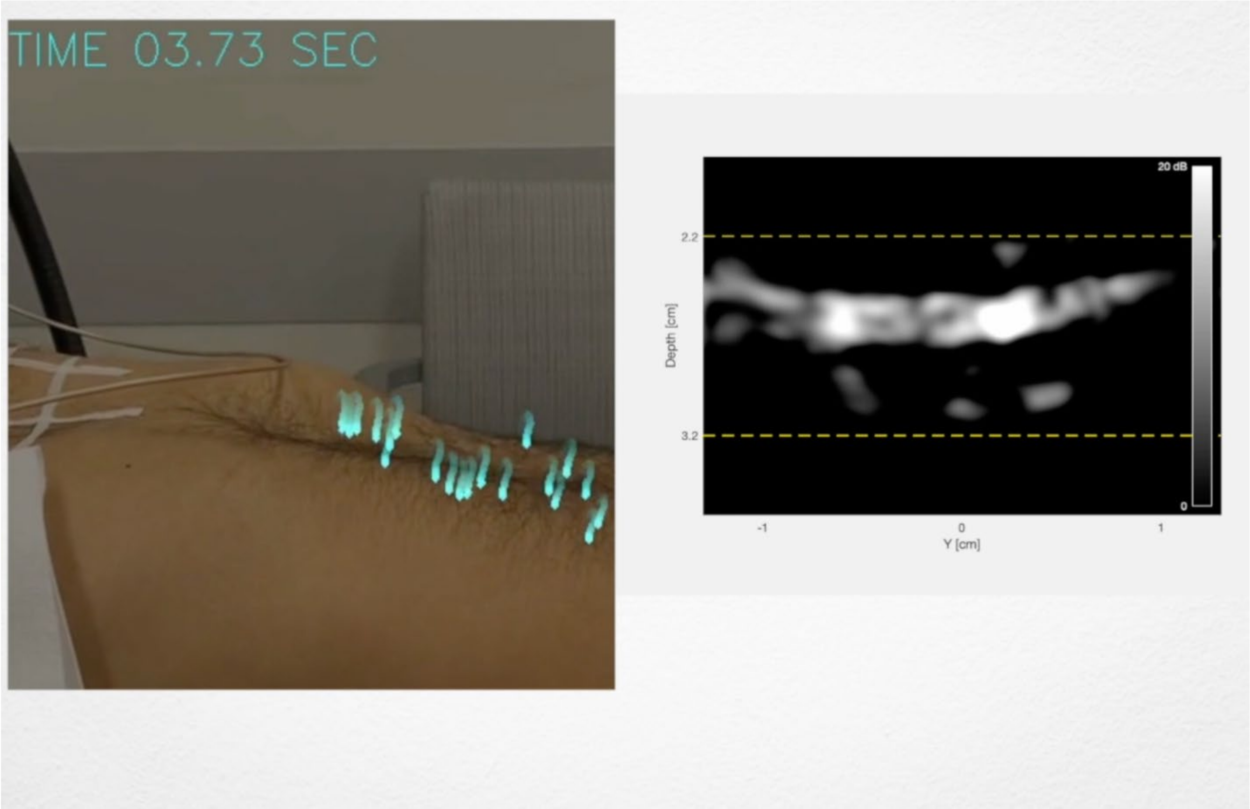


**Video S2. Chest motion alongside pleural brightness (ultrasound frame rate = 3 fps).** The subject's chest and abdomen is recorded (left panel) to track breathing cycles. The overlaid blue points are used for optical tracking to extract the motion of the chest against time. The real-time B-Mode image (right panel) displays the ROI (yellow box) and its brightness after Gaussian-blurring and filtering (see **Methods**)
